# NeuroDiscovery AI database: Comprehensive EHR dataset for Neurology

**DOI:** 10.1101/2025.06.01.25328302

**Authors:** S Selveshwari, Sonick Suri, Srinidhi Moodalagiri, Ansh Krishna, Nikita Chandra Kasivajjala

## Abstract

**Purpose:** The NeuroDiscovery AI database is a comprehensive real-world data (RWD) repository containing de-identified electronic health record (EHR) data from U.S.-based neurology outpatient clinics. The structured data encompasses sociodemographic details, clinical examinations, social, medical, and lifestyle histories, International Classification of Diseases (ICD-9/ICD-10) diagnoses, and prescribed medications. Additionally, the database integrates neuroimaging data and laboratory results, providing a robust resource for clinical research. This paper describes a subset of the NeuroDiscovery AI dataset and outlines the processes involved in its development.

**Participants:** As of October 15, 2024, the dataset includes EHR data from 355,791 patients, of whom 40.72% are male. Over 40.06% of the patients are aged 60 or older, spanning across 14,797 distinct diagnosis codes. The data represents more than 15 years of longitudinal patient information, with 26.87% of patients classified as active (defined as having had clinical encounters within the last 18 months). The median follow-up duration for active patients is 19.54 months.

**Limitation:** The large sample size, rigorous data processing, and robust data security of the NeuroDiscovery AI dataset are key strengths, enabling comprehensive studies on disease progression, treatment responses, and long-term outcomes in neurology. The dataset aligns closely with published demographic trends for various neurological conditions, including a female predominance in migraines, multiple sclerosis, and vertigo, with slight variations in age and gender distribution for conditions such as ALS. However, challenges remain, including missing data and data heterogeneity. Ongoing efforts to expand and diversify the dataset aim to improve its applicability and representativeness.

**Future plan:** The NeuroDiscovery AI dataset will expand by incorporating data from more providers and improving diversity, aiming to become one of the largest neurology-focused datasets. The platform will continue to evolve into a comprehensive analytical tool, integrating cohort building and data interrogation functionalities to streamline clinical workflows. These enhancements will enable faster, more accurate decision-making, and future efforts will focus on identifying key trends in neurological conditions and patient outcomes.

## Introduction

Neurodegenerative diseases are among the most complex and devastating conditions affecting the central nervous system. These diseases, marked by the gradual loss of multiple functions due to progressive neuronal damage, represent a significant and growing public health challenge due to their irreversible nature and rising prevalence. As the global population ages, the societal and economic burden of neurological conditions is expected to increase substantially. According to the Global Disease Burden Survey 2021 (GBD 2021 Forecasting Collaborators, 2024), the disability-adjusted life years (DALYs) due to neurological disorders was 113.5 million (95% CI: 69.18M - 178.9M) in 2022, with projections rising to 191.07 million (95% CI: 124.47M - 306.17M) by 2050. In the United States, DALYs due to neurological disorders were reported at 7.47 million (95% CI: 4.62M - 11.69M) in 2021(GBD 2021 Forecasting Collaborators, 2024).

For the U.S. population, the advancing age of the population further exacerbates the burden of neurodegenerative diseases. By 2060, an estimated 23% of the U.S. population will be aged 65 or older, representing 94 million individuals (Vespa et al., 2018). Dementia due to Alzheimer’s disease, which currently affects approximately 6.7 million people aged 65 and older in 2023, is projected to affect 13.8 million people by 2060 (Alzheimer’s Association, 2023). The economic burden of Alzheimer’s and related dementias in the U.S., which includes both formal and informal care costs and lost wages, is expected to rise from $305 billion (95% UR: $278–$333 billion) in 2020 to $2.2 trillion (95% UR: $1.3–$3.5 trillion) by 2060 (Nandi et al., 2024). While the at-risk population is expected to double over the next 30 to 40 years, the economic burden is projected to increase sevenfold, driven by rising healthcare costs, longer survival rates with neurodegenerative diseases, growing demand for long-term care, and the increasing prevalence of comorbid conditions among the elderly.

In the United States, Parkinson’s disease affected approximately one million people in 2020, with the number projected to increase to 1.2 million by 2030 (Marras et al., 2018). The annual economic burden of Parkinson’s disease is estimated at $52 billion, expected to rise to $80 billion by 2037 (Yang et al., 2020). In 2019, the U.S. reported the third-highest absolute number of migraine cases globally, with 3.8 million cases (95% CI: 3.43M - 4.37M) (Fan et al., 2023), and nearly 1 million individuals have been diagnosed with multiple sclerosis as of 2019 (Nelson et al., 2019).

Despite these trends, the pathogenesis of most neurodegenerative disorders remains poorly understood, and effective treatments are limited. Current diagnoses rely largely on clinical symptoms—such as cognitive, behavioural, speech, and motor impairments—supplemented by imaging studies to identify affected anatomical regions. However, reliable biomarkers for disease progression and definitive diagnostic tests are scarce, making early diagnosis and targeted treatment difficult.

The increasing availability of real-world data (RWD), particularly electronic health records (EHR), offers a powerful tool for advancing our understanding of diseases biology and disease pathways (Evans, 2016; Kataria & Ravindran, 2020; Tang et al., 2024). EHR data provides detailed, longitudinal clinical reporting on symptoms, treatments, and disease trajectories, enabling a comprehensive view of disease progression over time. When combined with neuroimaging, lab results, and other data sources, EHR data holds great potential for clinical trials research (Carrigan et al., 2020; Cowie et al., 2017; Hernán & Robins, 2016), uncovering new insights into disease mechanisms(Glicksberg et al., 2016; Tang et al., 2022; Woldemariam et al., 2023), identify subgroups in highly heterogeneous diseases such as Alzheimer’s (Parikh et al., 2021; Wang et al., 2020; Xu et al., 2020), identifying response to drugs (Shortreed et al., 2016; Voss et al., 2017), target identification (Davitte et al., 2022; Taubes et al., 2021), and informing personalized therapeutic strategies. The generation of RWE from the analysis of large-scale data sets that are representative of healthcare service delivery in routine settings provides a complementary source of data—as compared with more controlled clinical trials—which can support the assessment and approval of new treatments (Klonoff, 2020). RWE can also be used to guide decision-making for healthcare providers and payers since it more accurately represents how certain treatments are used in routine care and the associated outcomes (Roberts & Ferguson, 2021).

While large-scale real-world data (RWD) datasets based on electronic health records (EHR) exist for the general population (Carrigan et al., 2020; Johnson et al., 2023; *Northwestern Medicine Enterprise Data Warehouse (NMEDW)*) and also in specific fields such as oncology (*Overview of the SEER Program*), or neuropsychiatry (Patel et al., 2022), comprehensive neurology-focused datasets remain scarce and often lack the longitudinal depth necessary for studying the progression of neurodegenerative diseases. To address this gap, we have compiled what is expected to be one of the largest neurology-focused EHR datasets, drawing on longitudinal clinical and imaging data from leading outpatient neurology practices across the United States. This manuscript presents the initial subset of data from one of the largest outpatient neurology clinics in the United States of America. It provides an overview of the dataset, its variables, and early applications, with the aim of supporting future research in identifying disease progression trends, tracking symptom evolution, and discovering biomarkers that can improve the diagnosis and treatment of neurodegenerative diseases.

## Cohort description

### Participants and setting

The NeuroDiscovery AI dataset is a comprehensive collection of de-identified data from patients receiving care at various outpatient neurology practices across the United States. This dataset, in its complete form, is expected to be one of the largest neurology-focused EHR datasets in the USA, offering an invaluable resource for neurological research. The current manuscript describes the initial subset of data from this extensive database, which includes records from multiple large neurologic outpatient clinics across the United States. The dataset consists of de-identified Electronic Health Records (EHR) data from both outpatient visits and telemedicine services, along with associated imaging data and lab results, collected over a 15-year period (2008–2024).

The dataset includes 14,797 distinct ICD codes with records from 355,791 unique patients. Adhering to the OMOP data model recommendation (*Data Standardization – OHDSI*), active patients are defined as those with at least one encounter within the last 548 days (∼1.5 years) from the date of query and not reported as deceased. Based on this definition, 95,612 patients (∼ 27%) of the total cohort are classified as active.

Patients in the dataset have a median of 4 encounters, with a median follow-up duration of 3.35 months, reflecting both short-term and longer-term care engagements. A detailed summary of the descriptive statistics is provided in Table 2.

### EHR de-identification procedure

In accordance with the HIPAA Safe Harbor method, all 18 recommended Patient Health Information (PHI) elements were de-identified by either masking or removing them from the dataset. The following patient identifiers were completely removed or masked: patient names, all geographic subdivisions smaller than a state, telephone numbers, vehicle identifiers and serial numbers, fax numbers, device identifiers and serial numbers, email addresses, Uniform Resource Locators (URLs), social security numbers, Internet Protocol (IP) addresses, medical record numbers, biometric identifiers, health plan beneficiary numbers, full-face photographs, account numbers, certificate/license numbers, and other unique identifying numbers, characteristics, or codes.

Additionally, hospital-specific details such as facility IDs, specialty facility details, addresses, phone numbers, and fax numbers were de-identified. These elements were either replaced with generic tags (e.g., <Patient Name>, <Phone Number>, <Account Number>, <Hospital Name>, <Hospital Address>) or transformed to mask the sensitive information. For example, geographic information, such as zip codes, was truncated to retain only the first three digits, preserving gross demographic data while protecting individual privacy. Patient and encounter IDs were also mapped to completely different, unique values, ensuring no residual link to the original identifiers.

Dates were de-identified to ensure patient privacy while retaining their analytic value. The patient’s Date of Birth (DOB) was recorded by year only, with the month and day removed to minimize re-identification risk. To further protect patient privacy and prevent identification through exact encounter, test, or death dates, while also preserving the internal sequence of events for research purposes, we applied a random offset method. Each date was shifted by a random offset between −38 to −30 days or +30 to +38 days, with the specific offset unique to each patient. This method ensures that the chronological order of events is maintained, facilitating longitudinal analyses, while the original dates remain protected.

### Data movement and Transformation

The de-identification process was performed within the client’s server environment before the data was transferred to NeuroDiscovery AI servers.

De-identified data from the client server is securely transferred to the NeuroDiscovery server, where it is stored as compressed (zipped) XML flat files in a data lake. During transit, encryption protocols and access controls are employed to ensure data security. A comprehensive Medallion architecture is used to transform and store the data, ensuring that raw data undergoes meticulous cleansing, transformation, and enrichment, ultimately producing a refined dataset that is ready for advanced downstream analytics. The data transformation process at the various layers is outlined below:

1. *Bronze Layer:* In the first step, data from blob storage is transformed into JSON files and stored in the NoSQL database layer, which is optimized for handling semi-structured data. This layer, referred to as the *Bronze layer*, contains raw data in its most granular form, allowing for the capture of semi-structured information that can be easily accessed and processed.
2. *Silver Layer:* The next step moves the data into a hybrid layer that combines NoSQL and relational databases (RDBMS), supporting both JSON files and structured data. This *Silver layer* is where the data is cleaned to remove duplicates, correct errors, and ensure data quality. During this phase, appropriate schemas and tables are defined, transforming the raw data into a more structured and organized format. ETL (Extract, Transform, Load) tools are used to integrate data from NoSQL databases into RDBMS systems, ensuring seamless movement and transformation of data.
3. *Gold Layer:* The final step is the *Gold layer*, which is a relational database optimized for structured data that conforms to a Unified Data Model (UDM). This layer supports complex queries and analytics, providing a unified view of integrated, curated data sets. The *Gold layer* represents the most refined version of the data, which is ready for use in advanced downstream analytics.

At each stage of this transformation process, data validation is performed to ensure data quality across five key dimensions: completeness, correctness, concordance, plausibility, and currency. These criteria ensure that the data is accurate, timely, and reliable for subsequent analysis.

**Figure 1.**
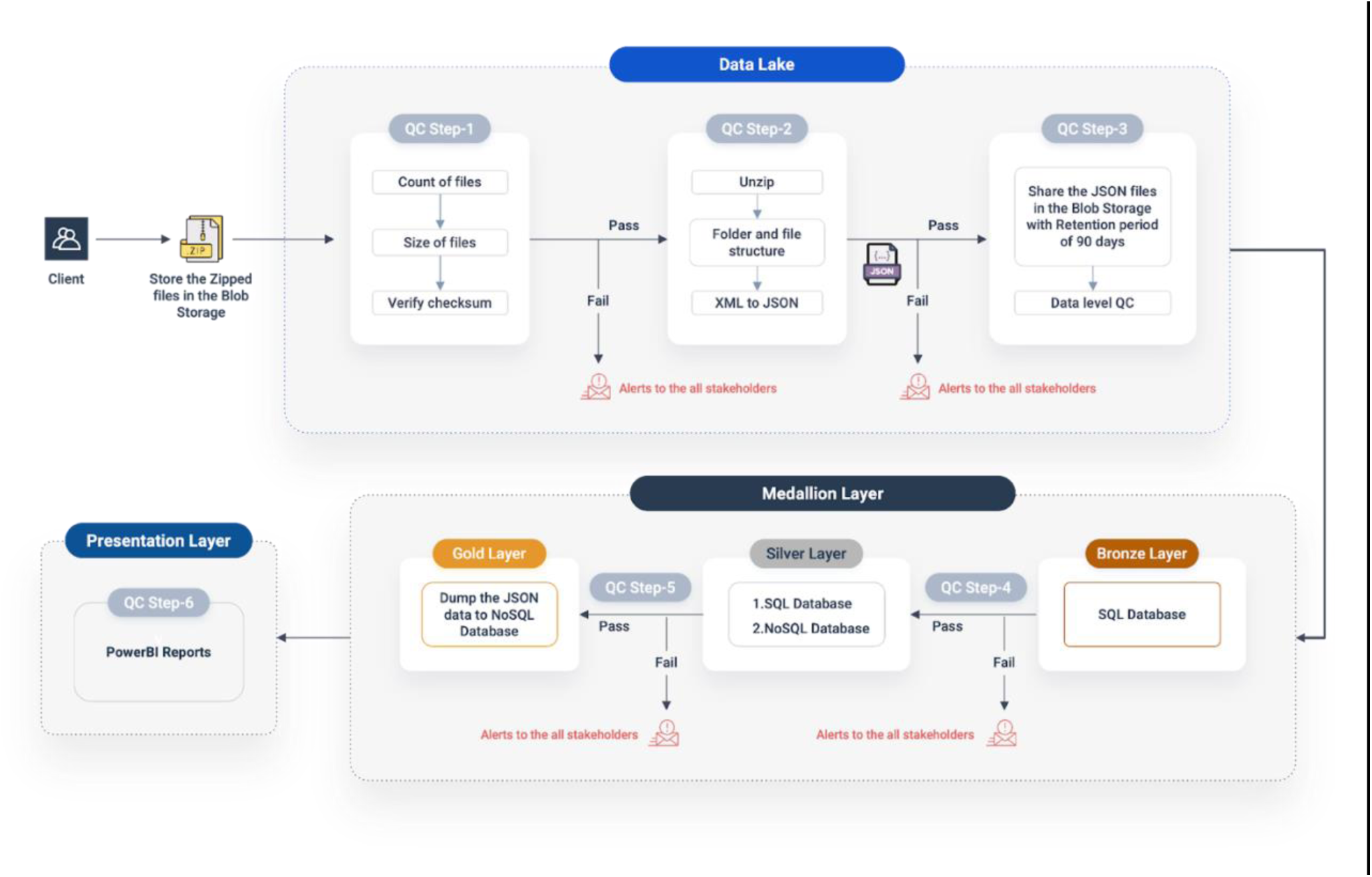
Overview of Data Movement, Transformation, and Quality Control (QC) Process. The workflow illustrates the Medallion architecture for data storage and transformation, along with the quality control framework applied at each stage of the process.

### Imaging de-Identification procedure

The data on the client server were assessed to understand the distribution of data including imaging studies, patient records, and associated metadata. After comprehensive assessment and bookkeeping, PACS files were transferred from the client PACS server to a staging server(mini PACS) for de-identification. A REST API was employed to anonymize patient-level information. In accordance with DICOM PS3.6 2024c recommendations, both direct and indirect identifiers were removed from all DICOM data and associated metadata using a combination of dynamic data masking techniques and data access controls.

Built-in plugins from Orthanc PACS were utilized to de-identify metadata for Patient Level Information, Study Level Information, Series Level Information, Equipment Level Information, and Institution Level Information. Table 1 gives a comprehensive view of all the de-identified elements and the method used to de-identify them. Additionally, tools such as DCMTK, OpenCV, and Tesseract were used to de-identify embedded PHI which is burned into the image pixel data.

**Table 1:** List of de-identified elements in imaging data.

| PHI element | De-identification |
| --- | --- |
| *Patient Level Information* |  |
| Patient Name | Removed |
| Patient ID | Replaced with NeuroDiscovery ID |
| Date of Birth | Removed |
| Sex | Removed |
| Other Patient IDs | Removed |
| Other Patient Names | Removed |
| Ethnic Group | Removed |
| Contact Details | Removed |
| *Study Level Information* |  |
| Study Date | Offset applied |
| Study Time | Removed |
| Referring Physician Name | Removed |
| Accession Number | Removed |
| Study ID | Removed |
| Study Description | Removed |
| *Series Level Information* |  |
| Series Date | Offset applied |
| Series Time | Removed |
| Performing Physician Name | Removed |
| *Institutional Level Information* |  |
| Institution Name | Removed |
| Institution Department Name | Removed |
| Performing Physician Name | Removed |
| *Equipment Level Information* |  |
| Device Serial Number | Removed |
| Secondary Capture Device Manufacturer | Removed |

Once the data is de-identified, a combination of manual and automated techniques is employed to validate the de-identification process. First, Mini PACS’s web interface is used to manually inspect the metadata for any remaining PHI elements that were not properly de-identified. Additionally, the Mini PACS De-identification Plugin’s REST API is used to query the modified metadata and ensure that all PHI fields have been handled appropriately. Following validation, the data is securely transferred to the cloud-based PACS server hosted by NeuroDiscovery AI.

### Data variables

The data model we use has been internally designed with input from clinical neurologists to ensure that all relevant components routinely collected during patient visits are captured comprehensively. To facilitate data consistency and usability, we have standardized the various data elements, ensuring uniform semantics and structure across the dataset. The variable structure is derived from a unified data model, which organizes the data into a relational database. This structure allows for efficient data querying and retrieval, either through a predefined set of database queries available in the front-end interface or through direct SQL queries.

#### Structured data

The structured data includes a comprehensive range of variables covering patient demographics, encounter details, clinical assessments, and treatment plans (see supplementary tables S1-S7 for a complete list of data variables and their descriptions). Key demographic information includes de-identified patient IDs, encounter dates, age, gender, and location, along with social and lifestyle factors such as tobacco and alcohol use, exercise, and occupation. Clinical data captures ICD9/ICD10 diagnosis codes, procedure codes (CPT), and patient-reported information, including chief complaints, medical and surgical history, and current medications. Additionally, the dataset includes prescribed medications, procedures, vitals, immunizations, and diagnostic tests.

#### Unstructured data

NLP techniques were used to extract relevant information from notes such as physician notes, treatment plans, assessment, and progression notes. Relevant information from neurological and mental status assessments, covering speech, memory, motor function, reflexes, and cranial nerve evaluations, and other routine scores such as Mini Mental Status Examination (MMSE), Montreal Cognitive Assessment (MoCA), Unified Parkinson’s Disease Rating Scale (UPDRS) are extracted from the assessment notes.

For this purpose, we plan to use two methods: a Classical Machine Learning (ML) approach, which applies traditional techniques to structured data like medications and ICD/CPT codes, and an Ensemble Approach combining ElasticSearch with Large Language Models (LLMs). The classical ML approach allows for rapid processing of structured data, while the ElasticSearch and LLMs method efficiently retrieves and interprets unstructured data, such as clinical notes, enabling precise extraction of clinical measures like MoCA scores.

#### Lab results

Various lab results, stored as PDF files on the client system, are processed through a series of steps to extract and store their contents in our database. First, the PDF files are converted into JPEG images using PyMuPDF. We then use CRAFT (Character Region Awareness for Text detection) to detect and localize text regions in the images. These detected regions are merged and passed to PyTesseract and EasyOCR for text extraction. The extracted text is refined using Large Language Models (LLMs), which improve accuracy by correcting misread characters, resolving ambiguities, and validating context-based information like units, medical terms, and numerical values. Finally, the output, in JSON format, is validated to ensure that the data conforms to expected medical formats, units checked, and completeness verified, any inconsistencies are flagged and reviewed manually, before integration into our structured database.

#### Data privacy

To ensure adherence to HIPAA regulations and protect patient privacy, the following measures have been implemented. In addition to the de-identification of all PHI elements, all data is encrypted both in transit and at rest ensuring security at all stages of its lifecycle.

Access to data is controlled using Azure Role-Based Access Control (RBAC) and conditional access policies (KeyClock). Detailed logs of access and activity on sensitive data are maintained using Azure Monitor and Azure Security Center, allowing for continuous monitoring and auditing of data usage, with potential security issues being detected and addressed in real time.

#### Data security

The data security framework of the NeuroDiscovery AI platform has been designed to comply with stringent security standards, ensuring that patient data is fully protected.

All data in transit, including communications between databases and backend services, is encrypted using Transport Layer Security (TLS). Data at rest is encrypted using Azure Storage Service Encryption (SSE), which automatically encrypts data stored in Azure Blob Storage, Azure Files, and other Azure storage services.

### Front-end interface

The de-identified data at NeuroDiscovery AI is used to power various applications such as dashboard, cohort builder, and data interrogation tools.

#### Dashboard

Users can interact with the data within NeuroDiscovery’s platform using our comprehensive dashboard, built using power BI. With dashboards, we aim to transform complex patient data into insightful and interactive visualization, empowering healthcare providers and stakeholders with actionable insights into patient demographics, clinical outcomes, and overall healthcare performance. The report generated using the dashboards will help identify trends and patterns, supporting data-driven decision-making to enhance patient care and improve the accessibility and usability of patient data.

**Figure 2:**
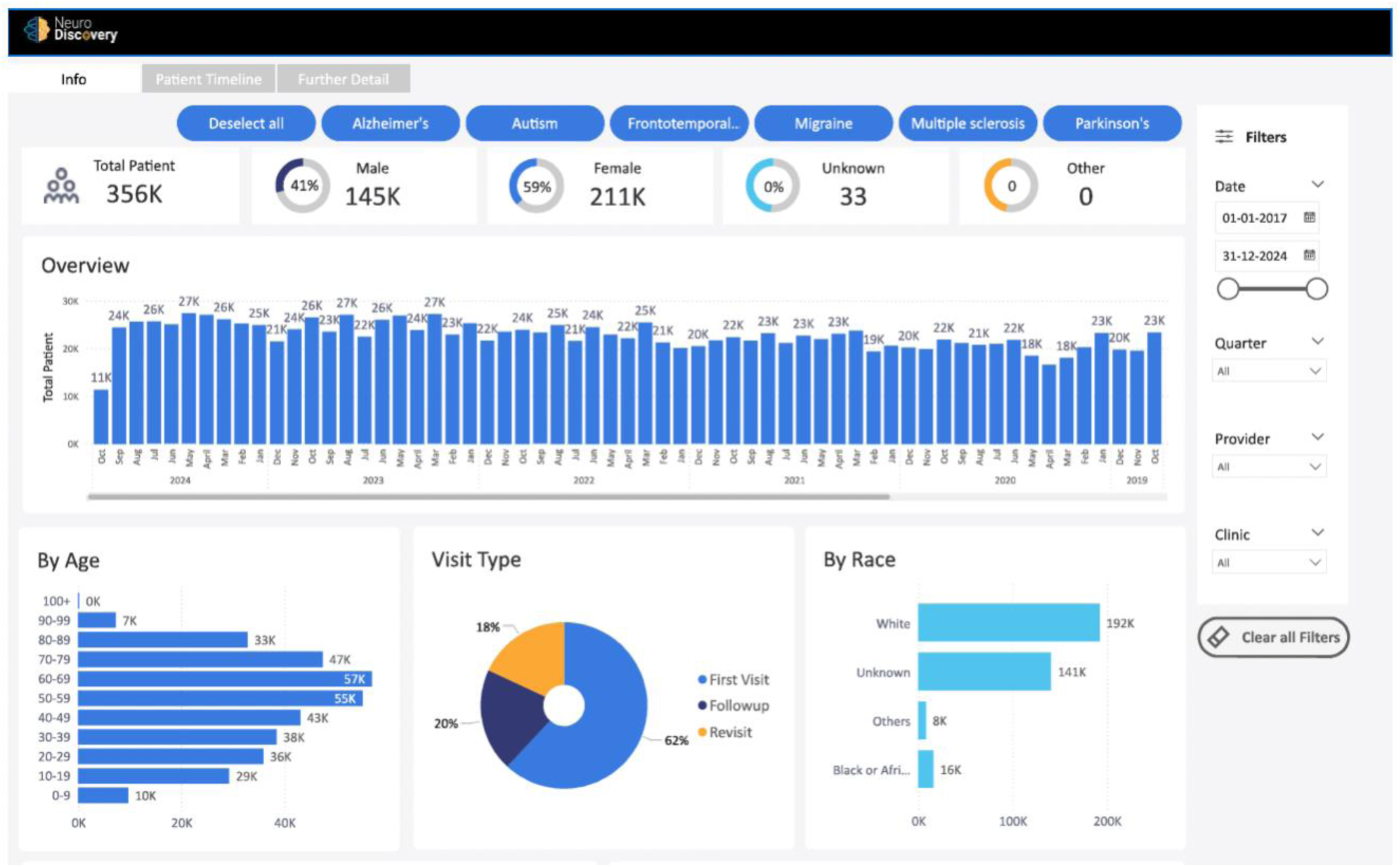
A representative image of the Data Dashboard.

#### Cohort builder

Another key application of the NeuroDiscovery dataset is the cohort builder tool, which enables the creation of highly specific patient cohorts. Leveraging a comprehensive dataset of active neurology patients, the platform integrates longitudinal electronic health records (EHRs), neuroimaging data, genomic data, and real-world evidence (RWE) to ensure that curated cohorts are representative of real-world clinical settings.

The cohort builder utilizes advanced machine learning algorithms, ElasticSearch, and large language models to optimize cohort selection, ensuring robust statistical power and minimizing biases in sample selection. Users can apply dynamic filtering, adjust inclusion and exclusion criteria, and visualize cohort characteristics in real time, offering a flexible and efficient approach to cohort development. This tool allows for the rapid identification of patient groups while maintaining data quality and relevance.

The tool can generate two types of outputs based on the specified inclusion and exclusion criteria: (A) Patient-specific lists, which detail all patients meeting the criteria. (B) Encounter-specific lists, which focus on specific patient encounters where the criteria are met.

#### Data interrogation tool

One of the key strengths of having a large dataset is the ability to query it effectively for relevant patient information. To enhance the efficiency, speed, and precision of data interrogation, we developed a Retrieval-Augmented Generation (RAG) pipeline. This tool enables healthcare practitioners to extract meaningful insights, streamlining clinical decision-making by providing quick access to critical patient data.

The architecture integrates embedding models, large language models (LLMs), and a vector database for seamless querying and retrieval. Large encounter records are segmented into manageable chunks, optimized for the context lengths supported by LLMs such as OpenAI’s Text Embedding v3, Biolord, or PubMedBERT, to generate dense data representations. These embeddings, along with associated metadata, such as encounter dates, categories, ranges, and chunk indices, are ingested into ChromaDB for persistent storage and efficient retrieval.

When a query is received, the algorithm retrieves the top-k most relevant chunks, ranked by relevance, ensuring only the most pertinent information is processed. The chat history is also incorporated to maintain continuity in conversations. These retrieved chunks, along with the user query, form an instruction-following prompt, which is then fed into an LLM, such as GPT-4o-mini, LLaMa8B, LLaMa70B, or Mistral-small, to generate a concise, accurate response.

#### Disclaimer on Figures

Figures 3 and 4 in this manuscript contain screenshots of the tool interface and are intended solely for illustrative purposes to demonstrate the front-end design and functionality. All names, dates, and other textual elements shown in these figures are fictitious and used as placeholders. They do not represent actual individuals, patients, or study participants, and no real identifiable information is presented.

**Figure 3:**
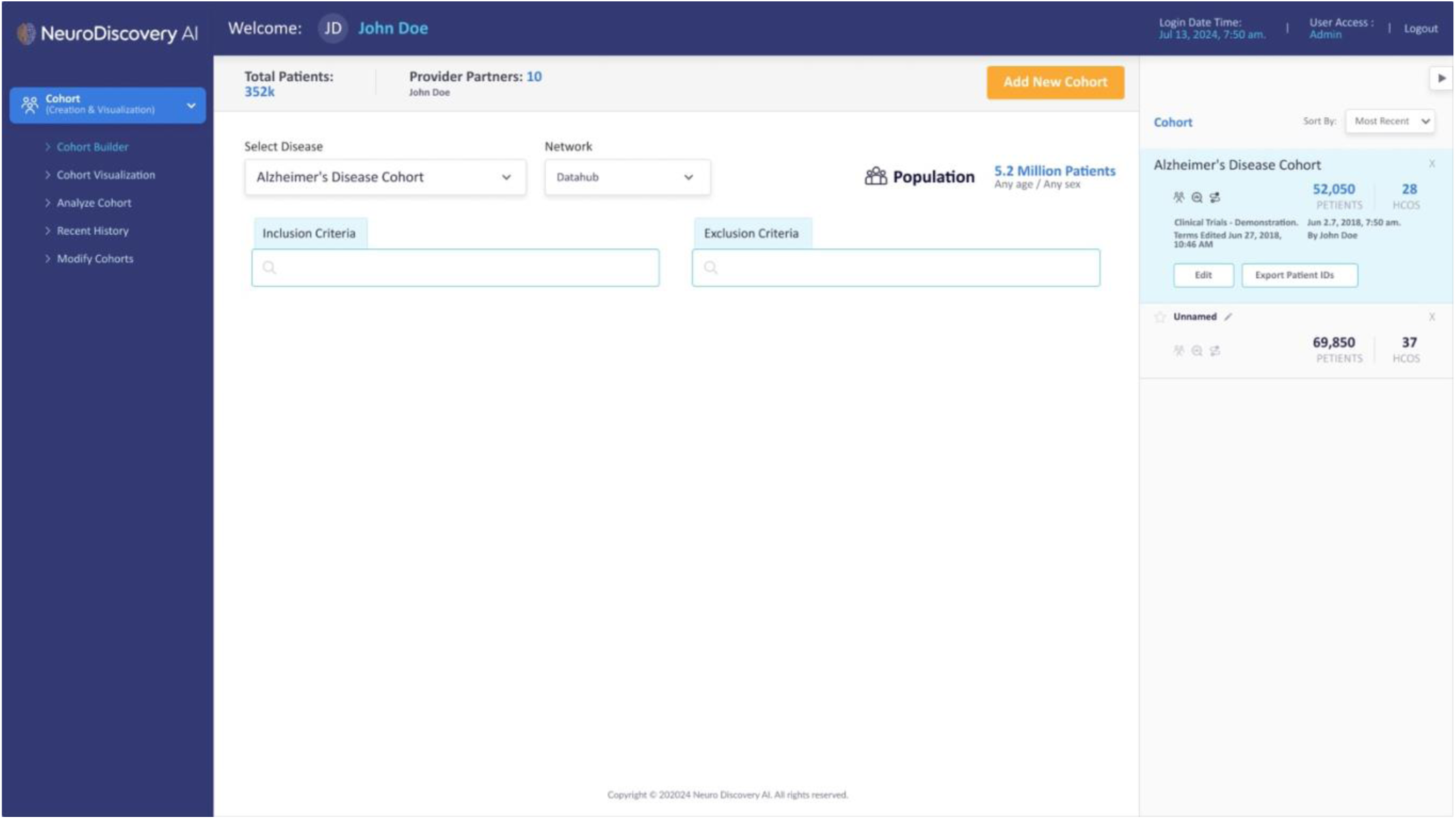
A representative image of the Cohort Builder Tool.

**Figure 4:**
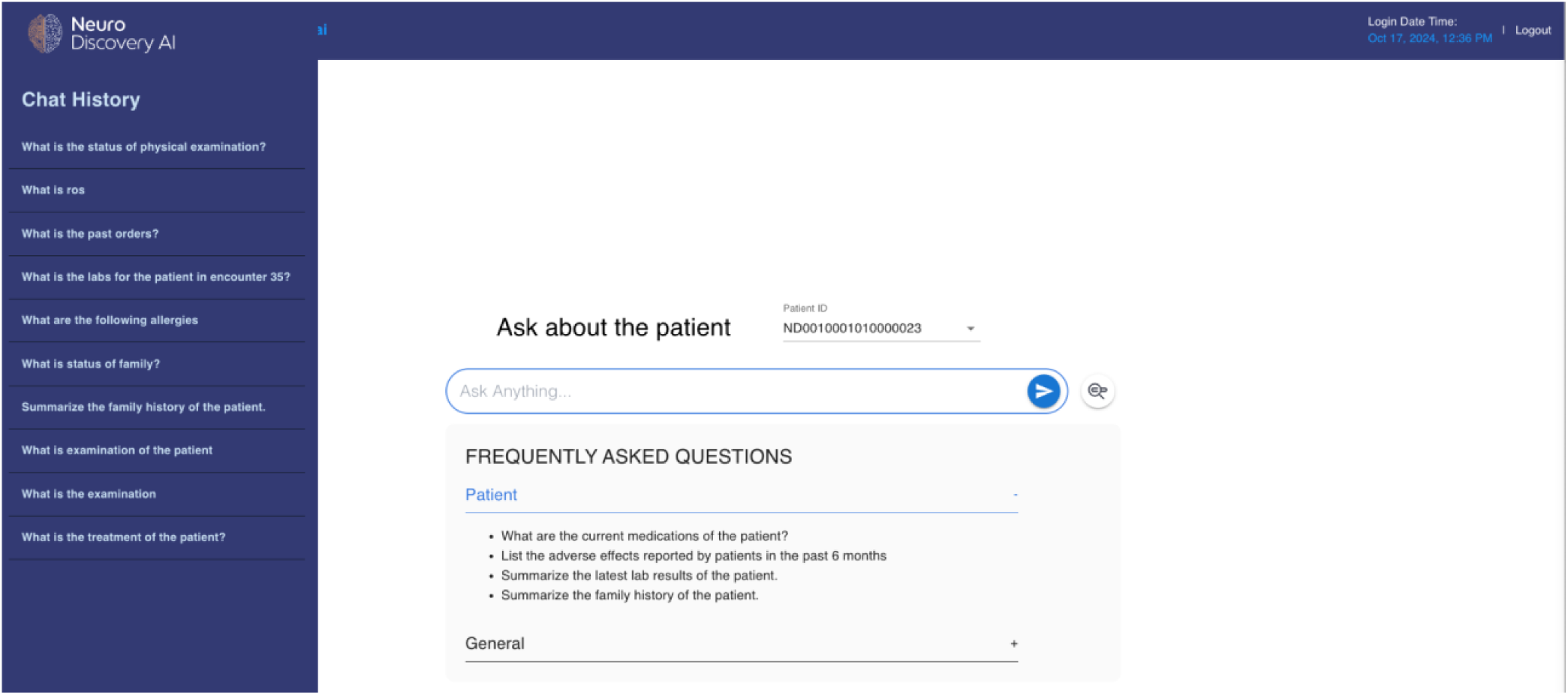
A representative image of the Data Interrogation Tool.

The sample and patient identifiers shown in Figure 4 are locally generated within our team and are not known or accessible to anyone outside the research team. These identifiers do not carry any risk of re-identification.

As such, no actual personal, clinical, or confidential data is depicted in these figures. This clarification is provided in accordance with journal policy on the use of representative images and placeholder identifiers.

### Participant characteristics

The current dataset comprises 355,791 patients, of whom 95,612 are classified as active as of October 15, 2024. Active patients are defined as those having at least one encounter within the past 1.5 years and not flagged as deceased. The dataset exhibits a female majority, with 59.27% of patients identifying as female. Racial distribution is predominantly white (53.89%), and over 40.06% of individuals are aged 60 or older at their most recent encounter.

When compared to other large EHR or registry-based datasets, such as the neuropsychiatry-focused NeuroBlu (Patel et al., 2022), Alzheimer’s disease focused dataset from The National Alzheimer’s Coordinating Center (NACC) (*UDS Demographics and Diagnoses*), Shaip (*EHR Datasets for Machine Learning | Electronic Health Records*) a provider for unstructured data across multiple diseases, the NeuroDiscovery AI dataset shows comparable demographic patterns.

Specifically, the female majority observed in NeuroDiscovery AI’s dataset aligns with NeuroBlu (51.10%), Shaip (54%), and NACC (65%). Additionally, NeuroDiscovery AI’s dataset features a higher representation of older age groups, with 16.01% of patients aged 60–69 and 13.31% aged 70–79, compared to NeuroBlu, which has only 5.50% and 2.70% in these age categories, respectively.

Table 2 and Figure 5 summarizes the demographics of the patients characteristics in the dataset.

**Table 2:** Patient Demographics.

| Total Patients |  | % |
| --- | --- | --- |
| Patient count | 355,791 | 100% |
| Female | 210,878 | 59.27% |
| Male | 144,880 | 40.72% |
| Unknown | 33 | 0.01% |
| Active Patients |  |  |
| Patient counts | 95,612 | 100% |
| Female | 60,204 | 62.97% |
| Male | 35,380 | 37.00% |
| Unknown | 28 | 0.03% |
| Age on last encounter (Total patients) |  |  |
| 0-9 | 9,643 | 2.71% |
| 10-19 | 29,247 | 8.22% |
| 20-29 | 35,841 | 10.07% |
| 30-39 | 38,418 | 10.80% |
| 40-49 | 43,031 | 12.09% |
| 50-59 | 55,127 | 15.49% |
| 60-69 | 56,976 | 16.01% |
| 70-79 | 47,356 | 13.31% |
| 80-89 | 32,806 | 9.22% |
| 90-99 | 7,230 | 2.03% |
| 100+ | 116 | 0.03% |

**Figure 5:**
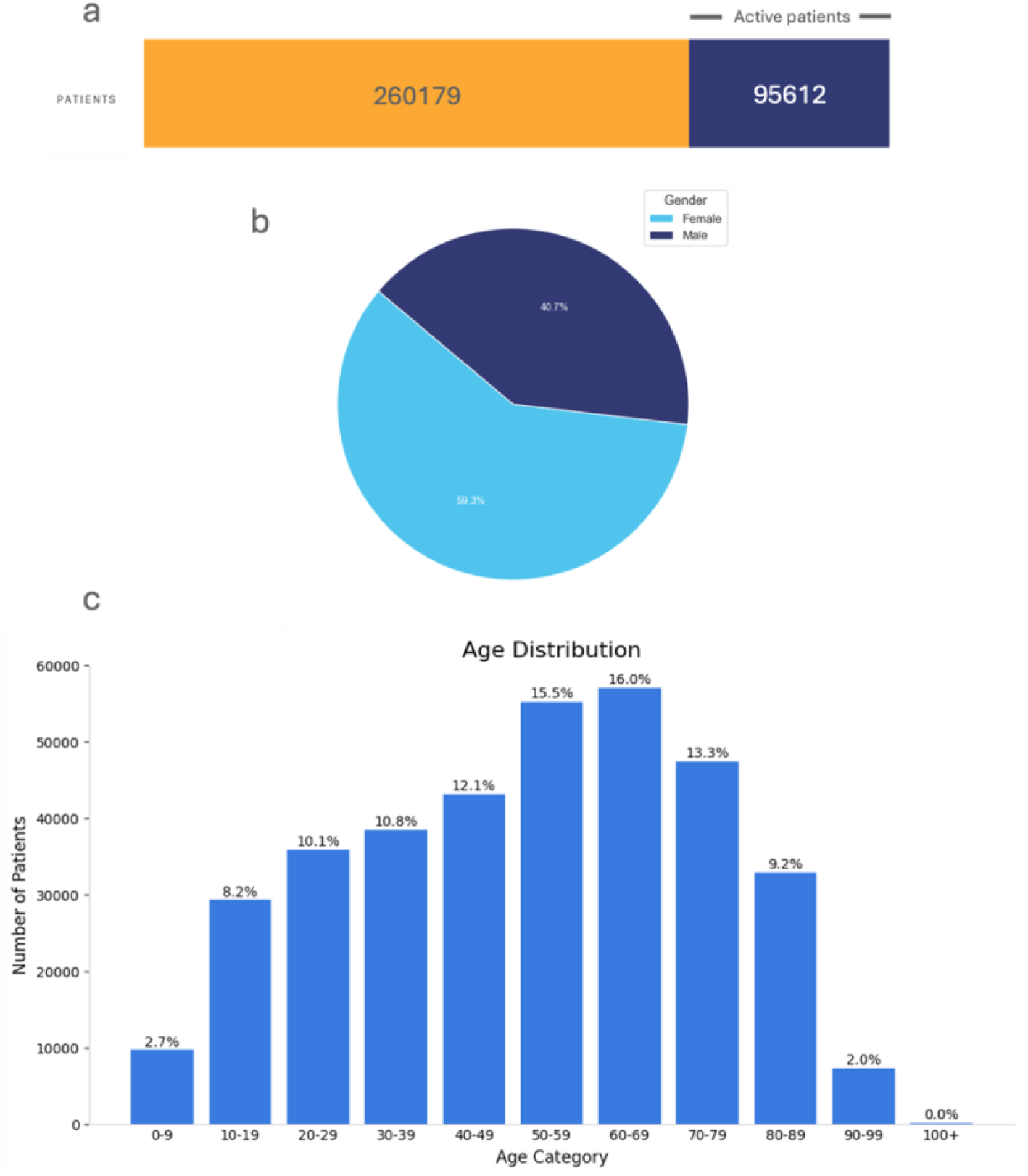
Overview of patient demographic distribution. (a) Distribution of historical and active patients, (b) distribution by gender, (c) distribution by age.

#### Encounter details

Individuals in the dataset had a median of 4 recorded encounters, increasing to a median of 9 encounters for active patients. For the majority of patients, the number of encounters ranged from 1 to 36, as indicated by the 5th to 95th percentile (Table 3 and Figure 6), indicating a highly skewed distribution, with skewness of 7.45. To further understand this variation, we analyzed the continuous observation period, defined as the time span between consecutive visits, provided that each visit is no more than 548 days apart (*Data Standardization – OHDSI*, n.d.-b). This approach emphasizes the period for continuous patient engagement and provides a more meaningful measure of observation period. The median observation period across the dataset was 2.06 months, with most patients (5th to 95th percentile) having an observation period ranging from 1 day to over 75.98 months (Table 3 and Figure 7). This distribution was also highly skewed with skewness of 3.25.

**Figure 6:**
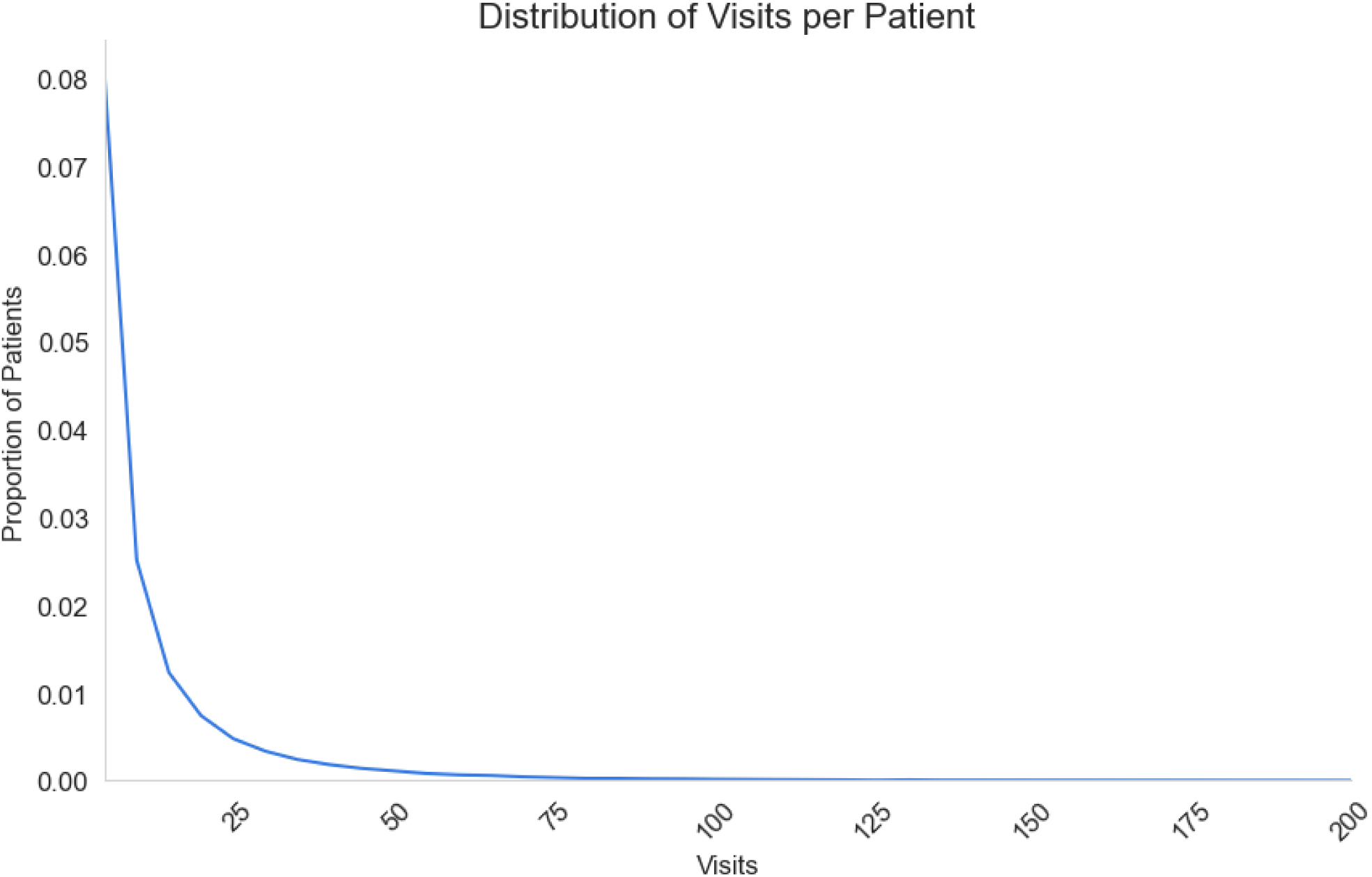
The distribution of number of visits per patients.

**Figure 7:**
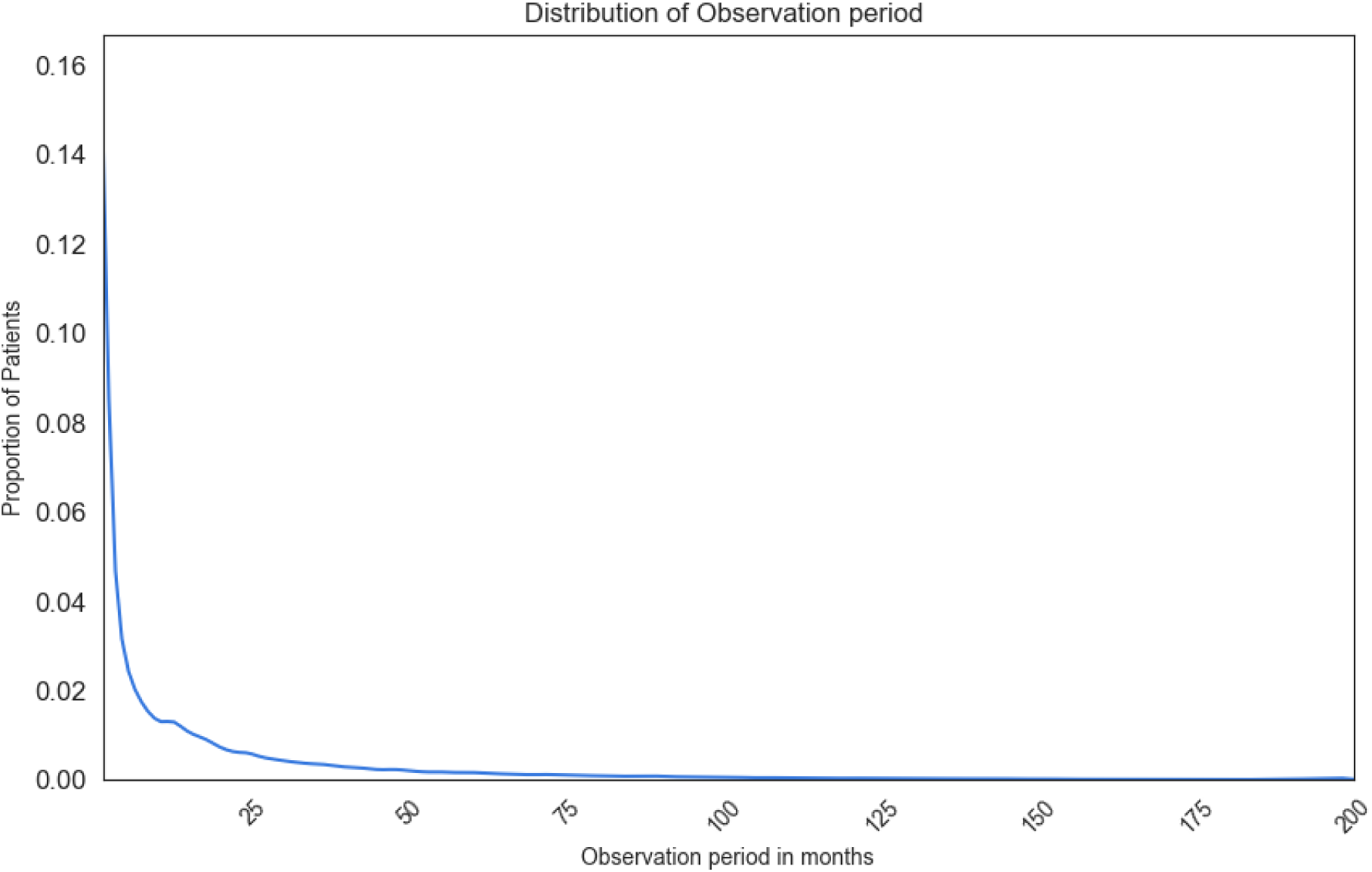
The distribution of observation duration per patients.

**Table 3:** Encounter details within dataset.

|  | Total Patients | Active Patients |
| --- | --- | --- |
| **Encounter Per Patient** |  |  |
| Mean (SD) | 9.23 (17.50) | 18.35 (27.58) |
| Median (IQR) | 4 (1-9) | 9 (3-22) |
| 5 and 95 Percentile | 1-36 | 1-66 |
| Skew | 7.45 |  |
| **Observation Period (Months)** |  |  |
| Mean (SD) | 14.71 (29.27) | 27.41 (41.63) |
| Median (IQR) | 2.06 (0.03 – 14.94) | 8.57 (0.49 – 35.64) |
| 5 and 95 Percentile | 0.03 – 75.98 | 0.03 – 123.91 |
| Skew | 3.25 |  |

| Follow-up Period for patients (Months) |  |  |
| --- | --- | --- |
| Mean (SD) | 18.76 (33.59) | 39.56 (48.30) |
| Median (IQR) | 3.35 (0.03 – 20.99) | 19.54 (2.85 – 59.85) |
| 5 and 95 Percentile | 0.03 – 93.33 | 0.03 – 149.17 |
| Skew | 2.71 |  |
| Patients with more than 2 years of FUP | 22.87% | 45.78% |

There was a notable increase over time in the proportion of patients with more than two encounters per year, rising from 60% in 2008 to 67% in 2023 (Figure 9), suggesting improved continuity of care. This trend is also reflected in the follow-up period, calculated as the cumulative length of each patient’s observation periods (*Data Standardization – OHDSI*, n.d.-a). A patient may have multiple observation periods, depending on their interactions with the healthcare system, and the follow-up period represents the sum of all these periods. The median follow-up duration was 3.35 months for all patients and 19.54 months for active patients, indicating sustained engagement in care. Although the distribution of the total follow-up period was highly skewed, with nearly 25% of individuals having just one encounter, more than 25% of patients (3rd quartile) had a follow-up period of over 20 months, with some active patients having follow-up durations of over 12 years (Table 3 and Figure 8). Table 3 provides a detailed summary of these metrics.

**Figure 8:**
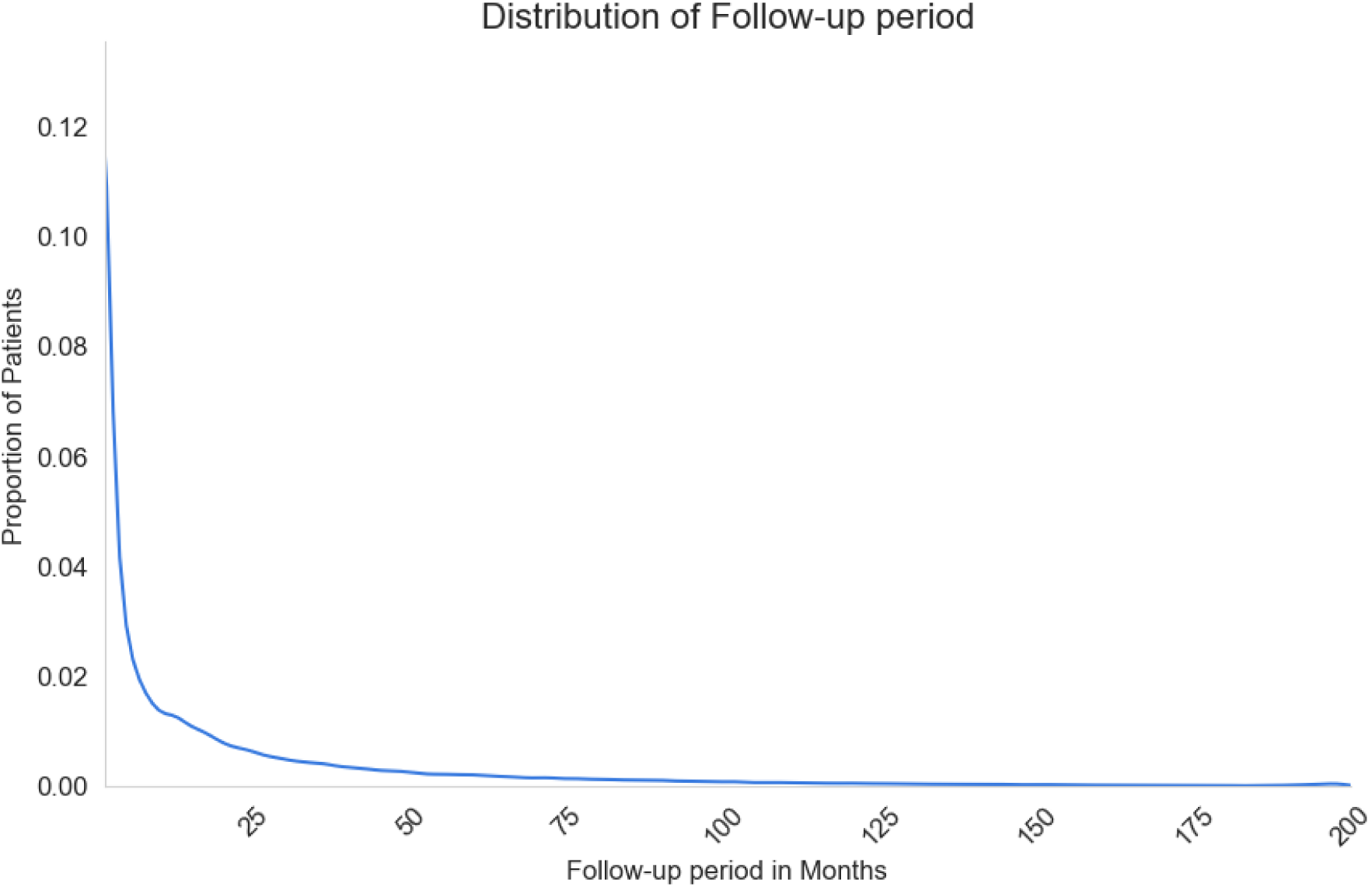
Distribution of followup duration per patients.

**Figure 9:**
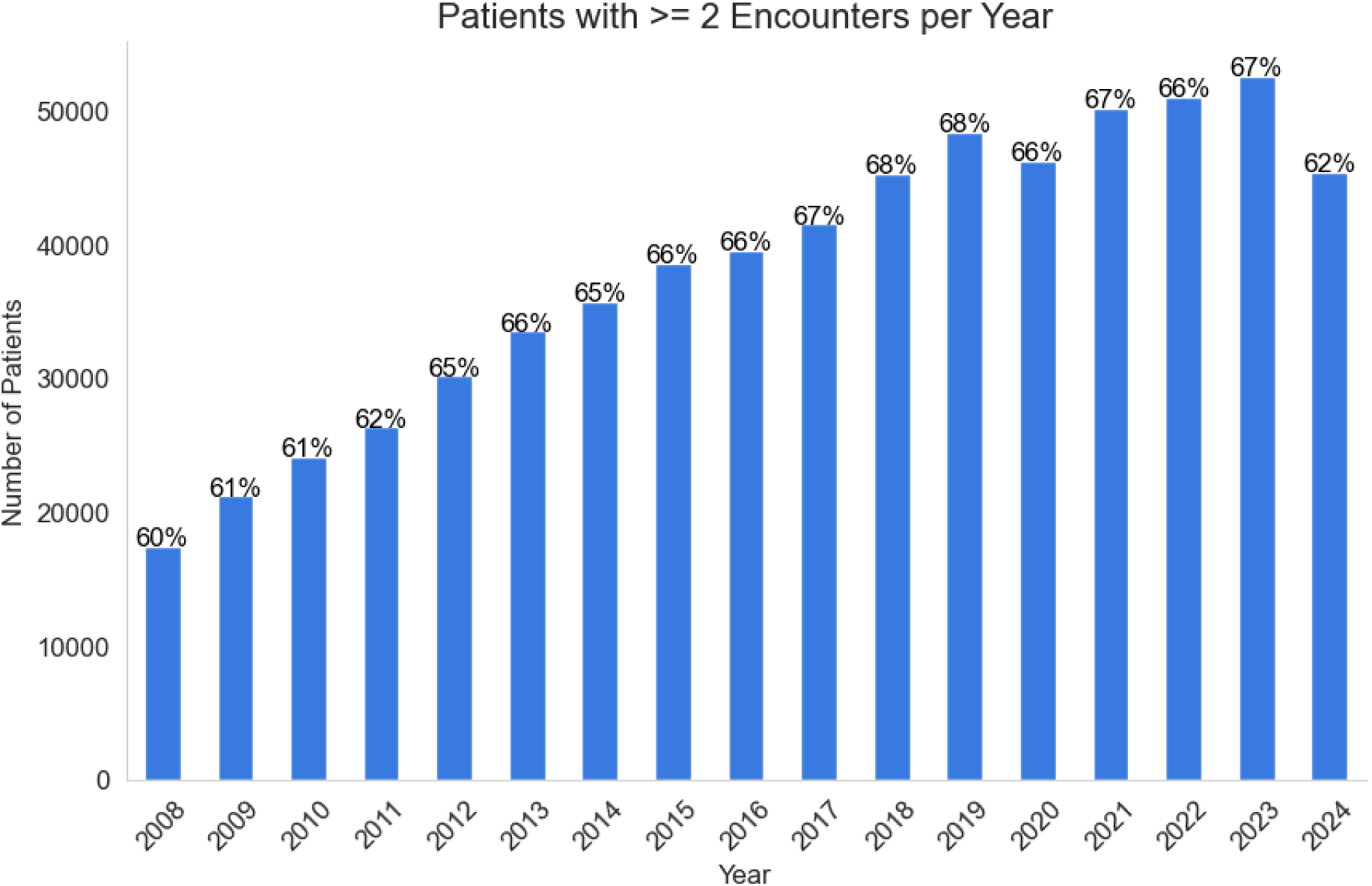
Change in number of patients with consecutive visits over years.

#### Diagnosis distribution

Table 4 provides a detailed distribution of patient counts across the top neurological conditions, highlighting disease prevalence and gender differences. The most frequent diagnosis is Migraine and Headache, representing 27.87% of all patient records, followed by Dizziness and Giddiness at 15.29%. A strong female predominance is noted for both conditions, with 73.02% of migraine cases and 68% of vertigo cases in women, consistent with studies reporting 2 to 3 times higher prevalence in women for migraines (Burch et al., 2021; Cohen et al., 2024) and 66% female prevalence in vertigo (Ruthberg et al., 2021).

**Table 4:** Description of some of the top neurological conditions.

| **Disease** | **Total Patients** | **Active Patients** | **Age (at first diagnosis)**<br>**Mean (SD)** | **Age Median (IQR)** | **% Male** |
| --- | --- | --- | --- | --- | --- |
| Migraine and Headache | 99,145 | 36,629 | 35 (18) | 33 (19 - 49) | 26.98% |
| Dizziness and Giddiness | 54,415 | 20,156 | 49 (22) | 51 (31 - 66) | 32.41% |
| Cerebral Infarction (Stroke) | 19,112 | 4,407 | 68 (16) | 70 (58 - 80) | 45.41% |
| Epilepsy | 12,078 | 5,456 | 43 (22) | 43 (24 - 60) | 48.10% |
| Unspecified Dementia | 9,027 | 3,196 | 79 (9) | 80 (74 - 85) | 39.85% |
| Parkinson's Disease | 6,737 | 1,941 | 73 (11) | 74 (66 - 80) | 57.22% |
| Alzheimer's Disease | 5,106 | 1,501 | 79 (9) | 80 (74 - 85) | 34.86% |
| Multiple Sclerosis | 4,507 | 2,137 | 48 (14) | 48 (37 - 58) | 25.47% |
| Autism | 1,603 | 936 | 16 (11) | 14 (8 - 21) | 75.42% |
| Frontotemporal Dementia | 859 | 149 | 74 (10) | 75 (68 - 80) | 45.63% |
| Amyotrophic Lateral Sclerosis | 649 | 90 | 66 (12) | 66 (59 - 75) | 54.85% |
| Traumatic Brain Injury | 267 | 121 | 56 (21) | 58 (39 - 73) | 58.43% |

Similarly, Multiple Sclerosis shows a female majority with 74.53% of cases in women, higher than the previously reported 69% (Walton et al., 2020), with the mean age of diagnosis in our dataset being 48 years, compared to the reported 32 years.

The mean age of stroke in our dataset is 78 years, slightly higher than the reported 66-70 years (Madsen et al., 2017), with a comparable gender distribution (45% males). For Alzheimer’s and dementia, the average age is 80 years, similar to the reported 73-81 years (Monfared et al., 2023), with a similar female predominance (60-65% vs. 55-63%).

Autism and Parkinson’s Disease show a higher proportion of male patients, at 75.42% and 57.22%, respectively. In Parkinson’s disease, our data reflects a mean age of 73 years and 57% males, consistent with previous studies (Devraj et al., 2024; Orozco et al., 2020). For ALS, our dataset shows 56% males, closely aligning with the 60% reported by the National ALS Registry (Raymond et al., 2019).

For a comprehensive summary, refer to Table 4. A full list of ICD codes and detailed descriptions of the top conditions can be found in the supplementary Table S8.

#### Missing data

Age was available for all patients in the dataset. Data were missing for the following variables:

1. Gender: 33 patients (0.01%).
2. Race: 140,512 patients (39.49%).
3. Date of birth: No Missing values

#### Imaging studies

The current imaging dataset includes a total of 650,231 imaging studies, collected over a period of 25 years, and conducted on 223,272 unique individuals. The predominant imaging modality is Magnetic Resonance Imaging (MRI), which constitutes approximately 72% of all studies. This is followed by Computed Tomography (CT) scans, accounting for 19% of the dataset. A detailed distribution of imaging modalities is provided in Table 5.

**Table 5:** Distribution of Imaging Modalities in the Dataset.

| Description | Modality | Count | % |
| --- | --- | --- | --- |
| Magnetic Resonance Imaging | MR | 377598 | 72.14 |
| Computed Tomography | CT | 90002 | 17.20 |
| Computed Radiography | CR | 4458 | 0.85 |
| Digital Radiography | DX | 23625 | 4.51 |
| Ultrasound | US | 24312 | 4.65 |
| X-ray Angiography | XA | 2786 | 0.53 |
| Nuclear Medicine | NM | 108 | 0.02063 |
| Positron Emission Tomography (PET) | PT | 135 | 0.02579 |
| Occupational Therapy Imaging | OT | 46 | 0.00879 |
| Radio Fluoroscopy | RF | 119 | 0.02274 |
| Document/Image (non-diagnostic) | DOC | 97 | 0.01853 |
| Key Object Selection (reference marker) | KO | 6 | 0.00115 |
| Presentation State | PR | 11 | 0.00210 |
| Mammography | MG | 2 | 0.00038 |
| Zonal Radiography | ZR | 5 | 0.00096 |
| Videofluoroscopy | VF | 26 | 0.00497 |
| General X-ray | XRAY | 3 | 0.00057 |
| Not Available/Applicable | N/A | 1 | 0.00019 |
| Hard Copy (scanned image) | HC | 4 | 0.00076 |
| X-ray (unspecified) | XR | 2 | 0.00038 |
| Audio | AU | 1 | 0.00019 |
| Secondary Capture | SC | 26 | 0.00497 |
| Structured Report | SR | 15 | 0.00287 |
| Positron Emission Tomography | PET | 2 | 0.00038 |

The majority of imaging studies were performed on individuals aged 50 to 70 years, representing approximately 40% of all scans in the dataset. A detailed age-wise distribution of imaging studies is presented in Table 6 and Figure 10.

**Table 6:** Distribution of imaging studies by age.

| Age on study date | Counts | % |
| --- | --- | --- |
| 00-09 | 8,786 | 1.41 |
| 10-19 | 39,600 | 6.34 |
| 20-29 | 45,626 | 7.30 |
| 30-39 | 63,144 | 10.11 |
| 40-49 | 83,084 | 13.30 |
| 50-59 | 1,18,633 | 18.99 |
| 60-69 | 1,18,154 | 18.91 |
| 70-79 | 97,520 | 15.61 |
| 80-89 | 46,674 | 7.47 |
| 90-99 | 3,448 | 0.55 |
| >100 | 54 | 0.009 |

**Figure 10.**
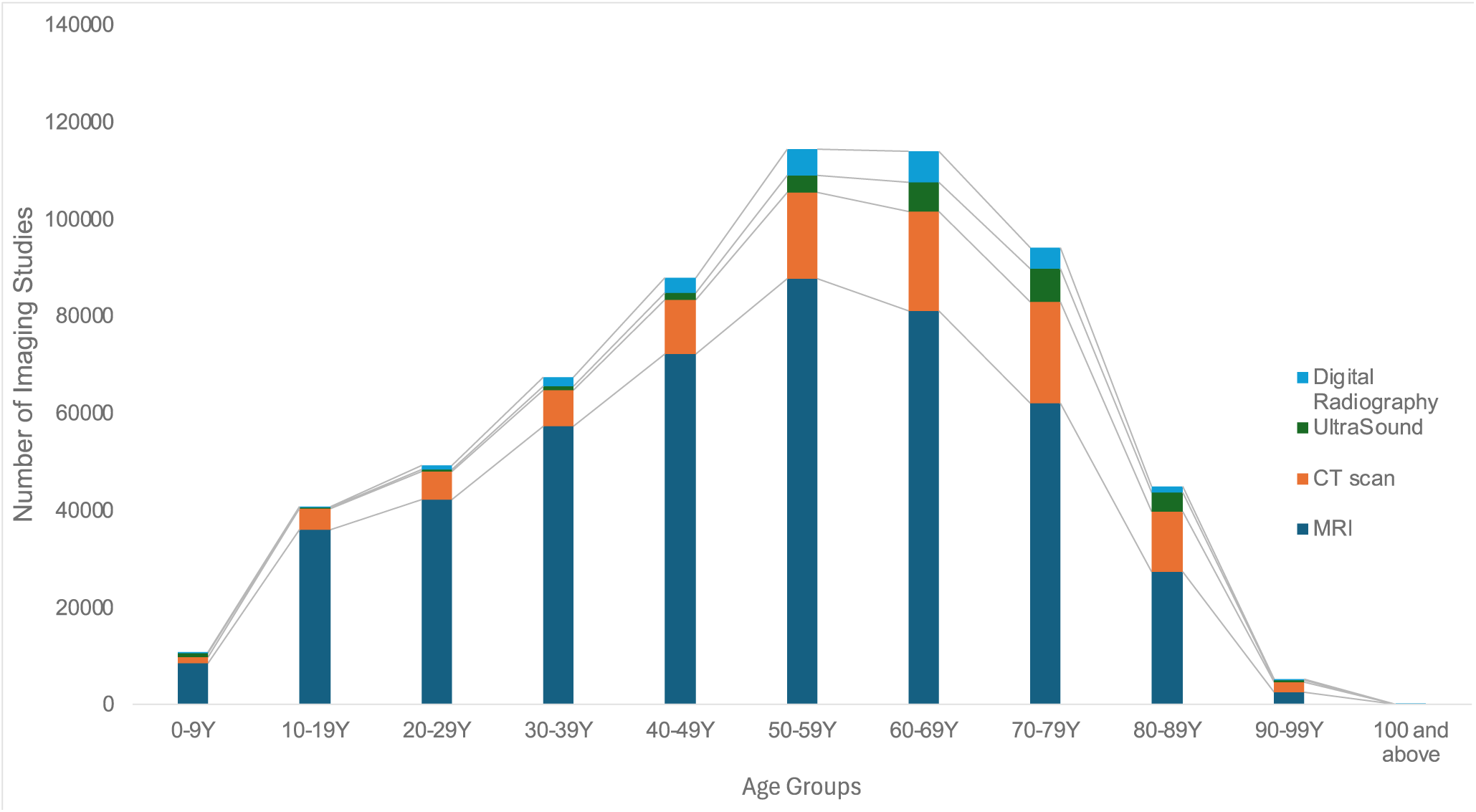
Distribution of Imaging Modalities by Age Group.

## Strengths and limitations

The NeuroDiscovery AI dataset’s large sample size is a key strength, significantly enhancing the power of statistical analyses across a wide array of neurological conditions. Rigorous data cleaning and processing, combined with robust security measures, ensure high-quality data that maintains patient privacy. The inclusion of both structured and unstructured data—including sociodemographics, clinical notes, and imaging reports—offers a comprehensive view of patient care. The follow-up duration has a median of 3.3 months with a heavily skewed right tail, allowing the analysis of both short-term and long-term patient outcomes. While 25% of patients have visited the facility only once, 25% are followed for over 20 months, with some extending beyond 7.75 years, enabling the study of immediate and extended clinical trajectories. This variation provides a unique opportunity to study long-term neurological outcomes, treatment responses, and disease progression patterns, facilitating research on questions not easily addressed through randomized controlled trials or prospective studies, such as the progression of neurodegenerative diseases (Davis et al., 2018; Lladó et al., 2021; Monfared et al., 2023), impact of comorbidities (Magyari & Sorensen, 2020), polypharmacy effects (Maust et al., 2021), and identification of early predictors of disease onset and severity (Li et al., 2023).

The dataset is not without limitations. As EHR data is primarily used for billing and clinical documentation, its level of detail may not always align with research needs, introducing variability and missingness in certain data elements (Kim et al., 2019). The inherent heterogeneity and incompleteness of EHR data may also affect data quality, and some variables may not fully comply with the FAIR (Findable, Accessible, Interoperable, and Reusable) principles (Kim et al., 2019). Unstructured data, while valuable, poses challenges in terms of extraction and completeness, leading to potential inaccuracies.

Moreover, this manuscript covers only an initial subset of the data, which may limit the generalizability of findings. Future expansions to include data from a wider range of neurology clinic providers will be critical to increasing the dataset’s representativeness and applicability across diverse patient populations.

## Future plans

The NeuroDiscovery AI dataset is continuously evolving to expand both its scope and functionality. Ongoing efforts to incorporate data from a wider range of providers will substantially increase the dataset’s size, and we anticipate that, upon completion, it will become one of the largest neurology-focused datasets available. A critical focus is also being placed on enhancing the dataset’s diversity, aiming for better representation across racial and ethnic groups to ensure more comprehensive and equitable insights.

A major priority is to further develop the platform into a comprehensive solution for data analysis. By integrating additional functionalities into the dashboard, we aim to create a one-stop solution for both clinical and research-based analytics.

The introduction of the cohort builder and data interrogation tools is expected to transform how clinical researchers and practitioners operate. These tools will enable more intuitive and powerful curation of patient cohorts and facilitate the extraction of relevant insights. Ultimately, they are poised to revolutionize clinical workflows by enabling faster, more accurate, and data-driven decision-making.

In addition to these technical enhancements, future work will focus on identifying and communicating clinically important trends in neurological conditions, particularly those with the potential to have a significant impact on healthcare practices. By analyzing both short- and long-term patient outcomes, we aim to uncover valuable insights into disease progression, treatment effectiveness, and emerging health risks in neurology.

## Data Availability

The data used in this study are not publicly available due to institutional and patient privacy considerations. However, we welcome research collaborations. If you are a neurology provider practice or health system in the United States and are interested in collaborating on biomarker discovery, novel target identification, multi-omics analysis, or other areas of clinical research, please contact us at.

## Collaborations

If you are a Neurology Provider Practice of Health System in USA and would like to collaborate on cutting edge research in the areas of Biomarker Discovery, Novel Target Discovery, Multi-OMICS Analysis or other areas of clinical research, please reach out to us at.

## Acknowledgements

We sincerely thank the members of the NeuroDiscovery AI team—Divya Veena, Garima Bajoria, Parinita Maithil, Sayantani Garai, Sneha K, Soorykant Suman, Divyansh Rajput, Jayant Verma, Alvin K Aloshyous, Devendra Patel, Ketki Suresh Khapare, Paras Mishra, and Vamsi Chandra Kasivajjala—for their dedicated efforts in building this database and contributing to various sections of the manuscript.

We are deeply grateful to our advisors—Prof. Howard Friedman and Dr. Ajith Thomas—for their invaluable guidance and constructive feedback, which greatly enhanced the quality of the manuscript. Finally, we extend our appreciation to all individuals at NeuroDiscovery AI for their collective contributions toward the development, refinement, and completion of this project.

We acknowledge the use of AI tools to improve the language of the manuscript.

