## Supplementary Material for "NeuroDiscovery AI database: Comprehensive EHR dataset for Neurology"

*Author Information*

1 NeuroDiscovery AI INC, GA, United States

2 Centennial High School, Frisco, TX, 75035

3 Fulton Science Academy Private School, Alpharetta, GA, 30004

Corresponding author; email: selveshwari{at}neurodiscovery.ai

### Supplementary Materials

#### Encounter Details

Table S1. provides an overview of the key variables captured for each patient encounter, including de-identified identifiers, appointment details, and demographic information.

| Variable Name | Variable Description |
|---|---|
| Patient ID | Each patient is assigned a unique ND ID, ensuring consistency across the database while maintaining de-identification. |
| Encounter ID | Each encounter is assigned a unique ID, recorded sequentially for each patient to capture the continuity of care. |
| Encounter Date | The date associated with each encounter, recorded with a pre-determined offset to ensure patient privacy while preserving the chronological sequence of events. |
| State of Residence | Patients' address details are limited to the state of residence to protect personal privacy. |
| Age | The patient's age at the time of the encounter. |
| Gender | The recorded gender of the patient. |
| Observation Period | The time span between the patient's first and last recorded encounter, ensuring that all consecutive visits are no more than 1.5 years apart. |
| Follow-up period | The cumulative duration over which the patient's details are recorded, sum of all observation periods. |

#### Clinical Information

Table S2. provides an overview of the essential clinical information recorded during each patient encounter, including diagnosis codes (ICD-9/ICD-10), procedure codes (CPT), and general clinical assessments such as vitals, height, weight, and BMI.

| Variable Name | Variable Description |
|---|---|
| Diagnosis codes (ICD9 / ICD 10) | International Classification of Diseases (ICD) codes used to document patient diagnoses. Diagnoses are recorded as both ICD9 and ICD10 codes. |
| CPT codes | Current Procedural Terminology (CPT) codes used to document medical procedures and billing information during the encounter. |

| Variable Name | Variable Description |
|---|---|
| **Vitals** | Key vital signs recorded during the encounter |
| Height | |
| Weight | |
| BMI | |
| Heart Rate | |
| Blood Pressure | |
| **General Assessment** | Overall clinical evaluation of patients including the following components |
| Appearance | |
| Eyes | |
| Heart | |
| Extremities | |

#### Neurological Assessment

Table S3. provides an overview of the neurological assessments performed during each patient encounter, including mental status examinations (MMSE), evaluations of speech, language, memory, motor function, reflexes, and cranial nerve assessments. These variables offer comprehensive insights into the neurological health of patients and assist in tracking disease progression, cognitive function, and motor abilities over time.

| Variable Name | Variable Description |
|---|---|
| **Neurological Examination** | Comprehensive evaluation of the patient's neurological health, covering motor and sensory functions. Some of the routinely recorded neurological exams are listed below |
| Mental Status Examination | |
| Speech | |
| Language | |
| Memory | |

| Variable Name | Variable Description |
|---|---|
| Fund of Knowledge | |
| Muscle Tone and Bulk | |
| Reflex | |
| Coordination | |
| Gait and Station | |
| Sensory Function | |
| **Cranial Nerve Examination** | Comprehensive assessment of cranial nerves II, III, IV, V, VI, VII, VIII, IX, X, XI, XII to detect abnormalities. |

#### Patient-Reported Variables

Table S4. provides an overview of the key patient-reported information documented during each encounter, including details on the visit type, chief complaints, current medications, and relevant medical and family histories.

| Variable Name | Variable Description |
|---|---|
| Visit Type | The category of the patient visit, such as first visit, routine check-up, or follow-up. |
| Chief Complaint | The primary reason or symptom reported by the patient that prompted the visit. |
| Current Medication | A list of medications the patient is actively taking at the time of the encounter, including dosage and frequency. |
| Allergies | Reported allergies to medications, foods, or environmental factors |
| Medical History | A detailed history of the patient's past and current medical conditions, including chronic illnesses. |
| Surgical History | Record of any past surgeries the patient has undergone, including the type and date of surgery. |
| Family History | Information about medical conditions prevalent in the patient's immediate family, which may indicate genetic risks. |

#### Social and Lifestyle History

Table S5. provides an overview of the key social and lifestyle factors reported by patients during each encounter, including information on tobacco, alcohol, drug use, and other lifestyle habits such as exercise, driving status, and employment history.

| **Variable Name** | **Variable Description** |
|---|---|
| Tobacco Use | Information on the patient’s current or past use of tobacco products, including frequency and duration. |
| Alcohol Use | Details regarding the patient’s alcohol consumption habits, including frequency and quantity. |
| Drug Use | Information about the use of recreational or illicit drugs. |
| Caffeine Use | Details on the patient’s consumption of caffeinated beverages, such as coffee or energy drinks, including frequency. |
| Medical Marijuana Use | Information on the patient’s use of medical marijuana |
| Exercise | The patient’s exercise habits, including type, frequency |
| Driving Status | Information on whether the patient is currently able to drive. |
| Fall History | Record of any recent falls experienced by the patient. |
| Marital Status | The patient’s current marital status. |
| Employment Status | Information on the patient’s current or past employment status, including occupation and work history. |
| Educational Status | The highest level of education completed by the patient, which may be relevant to cognitive or social assessments. |

#### Clinical History and Doctor’s Notes

Table S6. provides an overview of the key variables related to the patient’s clinical history and the doctor’s notes, with a focus on the history of present illness (HPI) and the physician's assessments.

| Variable Name | Variable Description |
|---|---|
| History of Present Illness (HPI) | A detailed narrative recorded by the physician that outlines the patient's current symptoms, the progression of the illness, and any relevant factors that may have contributed to the onset or worsening of the condition. |
| Doctor’s Notes under Assessments | Physician’s comprehensive assessment of the patient’s condition, including diagnostic impressions, clinical findings, and recommended treatment plans or next steps. |

#### Treatment Plan and Follow-Up

Table S7. provides an overview of the key variables that outline the patient’s treatment plan and follow-up care, including prescribed medications, procedures, counseling, and diagnostic tests such as laboratory and imaging orders.

| Variable Name | Variable Description |
|---|---|
| Treatment Plan | Physician’s recommended course of treatment |
| Medications | List of prescribed medications, including dosage, frequency, and duration, as part of the treatment plan. |
| Procedure | Details of any procedures recommended |
| Counseling | Information on any patient counseling sessions recommended, including topics like smoking, healthy living, diet, and exercise. |
| Laboratory Tests | Orders for lab tests, including CBC, metabolic panels, etc., |
| Imaging Order | Requests for imaging studies, such as X-rays, MRIs, or CT scans. |

#### Table S8. ICD codes and their descriptions

| Disease | ICD type | ICD Code | Description |
|---|---|---|---|
| Alzheimer's disease | ICD 10 | G30 | Alzheimer's disease |
| Alzheimer's disease | ICD 10 | G30.0 | Alzheimer's disease with early onset |
| Alzheimer's disease | ICD 10 | G30.1 | Alzheimer's disease with late onset |
| Alzheimer's disease | ICD 10 | G30.8 | Other Alzheimer's disease |
| Alzheimer's disease | ICD 10 | G30.9 | Alzheimer's disease, unspecified |
| Alzheimer's disease | ICD 9 | 331.0 | Alzheimer's disease |
| Parkinson's disease | ICD 10 | G20 | Parkinson's disease |
| Parkinson's disease | ICD 10 | G20.A | Parkinson's disease without dyskinesia |
| Parkinson's disease | ICD 10 | G20.A1 | Parkinson's disease without dyskinesia without mention of fluctuations |
| Parkinson's disease | ICD 10 | G20.A2 | Parkinson's disease without dyskinesia with fluctuations |
| Parkinson's disease | ICD 10 | G20.B | Parkinson's disease with dyskinesia |
| Parkinson's disease | ICD 10 | G20.B1 | Parkinson's disease with dyskinesia without mention of fluctuations |
| Parkinson's disease | ICD 10 | G20.B2 | Parkinson's disease with dyskinesia with fluctuations |
| Parkinson's disease | ICD 10 | G20.C | Parkinsonism, unspecified |

| | | | |
|---|---|---|---|
| Parkinson's disease | ICD 9 | 332 | Parkinson's disease |
| Parkinson's disease | ICD 9 | 332.0 | Paralysis agitans |
| Parkinson's disease | ICD 9 | 332.1 | Secondary parkinsonism |
| Multiple sclerosis | ICD 10 | G35 | Multiple sclerosis |
| Multiple sclerosis | ICD 9 | 340 | Multiple sclerosis |
| Autism | ICD 10 | F84.0 | Autistic disorder |
| Autism | ICD 9 | 299.0 | Autistic disorder |
| Autism | ICD 9 | 299.00 | Autistic disorder, current or active state |
| Autism | ICD 9 | 299.01 | Autistic disorder, residual state |
| Frontotemporal dementia | ICD 10 | G31.0 | Frontotemporal dementia |
| Frontotemporal dementia | ICD 10 | G31.01 | Pick's disease |
| Frontotemporal dementia | ICD 10 | G31.09 | Other frontotemporal neurocognitive disorder |
| Frontotemporal dementia | ICD 9 | 331.1 | Frontotemporal dementia |
| Frontotemporal dementia | ICD 9 | 331.11 | Pick's disease |
| Frontotemporal dementia | ICD 9 | 331.19 | Other frontotemporal dementia |
| Unspecified dementia | ICD 10 | F03 | Unspecified dementia |
| Unspecified dementia | ICD 10 | F03.9 | Unspecified dementia, unspecified severity |

| | | | |
|---|---|---|---|
| Unspecified dementia | ICD 10 | F03.90 | Unspecified dementia, unspecified severity, without behavioral disturbance, psychotic disturbance, mood disturbance, and anxiety |
| Unspecified dementia | ICD 10 | F03.91 | Unspecified dementia, unspecified severity, with behavioral disturbance |
| Unspecified dementia | ICD 10 | F03.911 | Unspecified dementia, unspecified severity, with agitation |
| Unspecified dementia | ICD 10 | F03.918 | Unspecified dementia, unspecified severity, with other behavioral disturbance |
| Unspecified dementia | ICD 10 | F03.92 | Unspecified dementia, unspecified severity, with psychotic disturbance |
| Unspecified dementia | ICD 10 | F03.93 | Unspecified dementia, unspecified severity, with mood disturbance |
| Unspecified dementia | ICD 10 | F03.94 | Unspecified dementia, unspecified severity, with anxiety |
| Unspecified dementia | ICD 10 | F03.A | Unspecified dementia, mild |
| Unspecified dementia | ICD 10 | F03.A0 | Unspecified dementia, mild, without behavioral disturbance, psychotic disturbance, mood disturbance, and anxiety |
| Unspecified dementia | ICD 10 | F03.A1 | Unspecified dementia, mild, with behavioral disturbance |
| Unspecified dementia | ICD 10 | F03.A11 | Unspecified dementia, mild, with agitation |
| Unspecified dementia | ICD 10 | F03.A18 | Unspecified dementia, mild, with other behavioral disturbance |
| Unspecified dementia | ICD 10 | F03.A2 | Unspecified dementia, mild, with psychotic disturbance |
| Unspecified dementia | ICD 10 | F03.A3 | Unspecified dementia, mild, with mood disturbance |
| Unspecified dementia | ICD 10 | F03.A4 | Unspecified dementia, mild, with anxiety |
| Unspecified dementia | ICD 10 | F03.B | Unspecified dementia, moderate |

| | | | |
|---|---|---|---|
| Unspecified dementia | ICD 10 | F03.B0 | Unspecified dementia, moderate, without behavioral disturbance, psychotic disturbance, mood disturbance, and anxiety |
| Unspecified dementia | ICD 10 | F03.B1 | Unspecified dementia, moderate, with behavioral disturbance |
| Unspecified dementia | ICD 10 | F03.B11 | Unspecified dementia, moderate, with agitation |
| Unspecified dementia | ICD 10 | F03.B18 | Unspecified dementia, moderate, with other behavioral disturbance |
| Unspecified dementia | ICD 10 | F03.B2 | Unspecified dementia, moderate, with psychotic disturbance |
| Unspecified dementia | ICD 10 | F03.B3 | Unspecified dementia, moderate, with mood disturbance |
| Unspecified dementia | ICD 10 | F03.B4 | Unspecified dementia, moderate, with anxiety |
| Unspecified dementia | ICD 10 | F03.C | Unspecified dementia, severe |
| Unspecified dementia | ICD 10 | F03.C0 | Unspecified dementia, severe, without behavioral disturbance, psychotic disturbance, mood disturbance, and anxiety |
| Unspecified dementia | ICD 10 | F03.C1 | Unspecified dementia, severe, with behavioral disturbance |
| Unspecified dementia | ICD 10 | F03.C11 | Unspecified dementia, severe, with agitation |
| Unspecified dementia | ICD 10 | F03.C18 | Unspecified dementia, severe, with other behavioral disturbance |
| Unspecified dementia | ICD 10 | F03.C2 | Unspecified dementia, severe, with psychotic disturbance |
| Unspecified dementia | ICD 10 | F03.C3 | Unspecified dementia, severe, with mood disturbance |
| Unspecified dementia | ICD 10 | F03.C4 | Unspecified dementia, severe, with anxiety |

| | | | |
|---|---|---|---|
| Unspecified dementia | ICD 9 | 294.2 | Dementia, unspecified |
| Unspecified dementia | ICD 9 | 294.20 | Dementia, unspecified, without behavioral disturbance |
| Unspecified dementia | ICD 9 | 294.21 | Dementia, unspecified, with behavioral disturbance |
| Migraine and Headache | ICD 10 | G43 | Migraine |
| Migraine and Headache | ICD 10 | G43.0 | Migraine without aura |
| Migraine and Headache | ICD 10 | G43.00 | Migraine without aura, not intractable |
| Migraine and Headache | ICD 10 | G43.001 | Migraine without aura, with status migrainosus |
| Migraine and Headache | ICD 10 | G43.009 | Migraine without aura, without status migrainosus |
| Migraine and Headache | ICD 10 | G43.01 | Migraine without aura, intractable |
| Migraine and Headache | ICD 10 | G43.011 | Migraine without aura, intractable, with status migrainosus |
| Migraine and Headache | ICD 10 | G43.019 | Migraine without aura, intractable, without status migrainosus |
| Migraine and Headache | ICD 10 | G43.1 | Migraine with aura |
| Migraine and Headache | ICD 10 | G43.10 | Migraine with aura, not intractable |
| Migraine and Headache | ICD 10 | G43.101 | Migraine with aura, not intractable, with status migrainosus |
| Migraine and Headache | ICD 10 | G43.109 | Migraine with aura, not intractable, without status migrainosus |
| Migraine and Headache | ICD 10 | G43.11 | Migraine with aura, intractable |

| | | | |
|---|---|---|---|
| Migraine and Headache | ICD 10 | G43.111 | Migraine with aura, intractable, with status migrainosus |
| Migraine and Headache | ICD 10 | G43.119 | Migraine with aura, intractable, without status migrainosus |
| Migraine and Headache | ICD 10 | G43.4 | Hemiplegic migraine |
| Migraine and Headache | ICD 10 | G43.40 | Hemiplegic migraine, not intractable |
| Migraine and Headache | ICD 10 | G43.401 | Hemiplegic migraine, not intractable, with status migrainosus |
| Migraine and Headache | ICD 10 | G43.409 | Hemiplegic migraine, not intractable, without status migrainosus |
| Migraine and Headache | ICD 10 | G43.41 | Hemiplegic migraine, intractable |
| Migraine and Headache | ICD 10 | G43.411 | Hemiplegic migraine, intractable, with status migrainosus |
| Migraine and Headache | ICD 10 | G43.419 | Hemiplegic migraine, intractable, without status migrainosus |
| Migraine and Headache | ICD 10 | G43.5 | Persistent migraine aura without cerebral infarction |
| Migraine and Headache | ICD 10 | G43.50 | Persistent migraine aura without cerebral infarction, not intractable |
| Migraine and Headache | ICD 10 | G43.501 | Persistent migraine aura without cerebral infarction, not intractable, with status migrainosus |
| Migraine and Headache | ICD 10 | G43.509 | Persistent migraine aura without cerebral infarction, not intractable, without status migrainosus |
| Migraine and Headache | ICD 10 | G43.51 | Persistent migraine aura without cerebral infarction, intractable |
| Migraine and Headache | ICD 10 | G43.511 | Persistent migraine aura without cerebral infarction, intractable, with status migrainosus |
| Migraine and Headache | ICD 10 | G43.519 | Persistent migraine aura without cerebral infarction, intractable, without status migrainosus |

| | | | |
|---|---|---|---|
| Migraine and Headache | ICD 10 | G43.6 | Persistent migraine aura with cerebral infarction |
| Migraine and Headache | ICD 10 | G43.60 | Persistent migraine aura with cerebral infarction, not intractable |
| Migraine and Headache | ICD 10 | G43.601 | Persistent migraine aura with cerebral infarction, not intractable, with status migrainosus |
| Migraine and Headache | ICD 10 | G43.609 | Persistent migraine aura with cerebral infarction, not intractable, without status migrainosus |
| Migraine and Headache | ICD 10 | G43.61 | Persistent migraine aura with cerebral infarction, intractable |
| Migraine and Headache | ICD 10 | G43.611 | Persistent migraine aura with cerebral infarction, intractable, with status migrainosus |
| Migraine and Headache | ICD 10 | G43.619 | Persistent migraine aura with cerebral infarction, intractable, without status migrainosus |
| Migraine and Headache | ICD 10 | G43.7 | Chronic migraine without aura |
| Migraine and Headache | ICD 10 | G43.70 | Chronic migraine without aura, not intractable |
| Migraine and Headache | ICD 10 | G43.701 | Chronic migraine without aura, not intractable, with status migrainosus |
| Migraine and Headache | ICD 10 | G43.709 | Chronic migraine without aura, not intractable, without status migrainosus |
| Migraine and Headache | ICD 10 | G43.71 | Chronic migraine without aura, intractable |
| Migraine and Headache | ICD 10 | G43.711 | Chronic migraine without aura, intractable, with status migrainosus |
| Migraine and Headache | ICD 10 | G43.719 | Chronic migraine without aura, intractable, without status migrainosus |
| Migraine and Headache | ICD 10 | G43.A | Cyclical vomiting |
| Migraine and Headache | ICD 10 | G43.A0 | Cyclical vomiting in migraine, not intractable |

| | | | |
|---|---|---|---|
| Migraine and Headache | ICD 10 | G43.A1 | Cyclical vomiting in migraine, intractable |
| Migraine and Headache | ICD 10 | G43.B | Ophthalmoplegic migraine |
| Migraine and Headache | ICD 10 | G43.B0 | Ophthalmoplegic migraine, not intractable |
| Migraine and Headache | ICD 10 | G43.B1 | Ophthalmoplegic migraine, intractable |
| Migraine and Headache | ICD 10 | G43.C | Periodic headache syndromes in child or adult |
| Migraine and Headache | ICD 10 | G43.C0 | Periodic headache syndromes in child or adult, not intractable |
| Migraine and Headache | ICD 10 | G43.C1 | Periodic headache syndromes in child or adult, intractable |
| Migraine and Headache | ICD 10 | G43.D | Abdominal migraine |
| Migraine and Headache | ICD 10 | G43.D0 | Abdominal migraine, not intractable |
| Migraine and Headache | ICD 10 | G43.D1 | Abdominal migraine, intractable |
| Migraine and Headache | ICD 10 | G43.8 | Other migraine |
| Migraine and Headache | ICD 10 | G43.80 | Other migraine, not intractable |
| Migraine and Headache | ICD 10 | G43.801 | Other migraine, not intractable, with status migrainosus |
| Migraine and Headache | ICD 10 | G43.809 | Other migraine, not intractable, without status migrainosus |
| Migraine and Headache | ICD 10 | G43.81 | Other migraine, intractable |
| Migraine and Headache | ICD 10 | G43.811 | Other migraine, intractable, with status migrainosus |

| | | | |
|---|---|---|---|
| Migraine and Headache | ICD 10 | G43.819 | Other migraine, intractable, without status migrainosus |
| Migraine and Headache | ICD 10 | G43.82 | Menstrual migraine, not intractable |
| Migraine and Headache | ICD 10 | G43.821 | Menstrual migraine, not intractable, with status migrainosus |
| Migraine and Headache | ICD 10 | G43.829 | Menstrual migraine, not intractable, without status migrainosus |
| Migraine and Headache | ICD 10 | G43.83 | Menstrual migraine, intractable |
| Migraine and Headache | ICD 10 | G43.831 | Menstrual migraine, intractable, with status migrainosus |
| Migraine and Headache | ICD 10 | G43.839 | Menstrual migraine, intractable, without status migrainosus |
| Migraine and Headache | ICD 10 | G43.9 | Migraine, unspecified |
| Migraine and Headache | ICD 10 | G43.90 | Migraine, unspecified, not intractable |
| Migraine and Headache | ICD 10 | G43.901 | Migraine, unspecified, not intractable, with status migrainosus |
| Migraine and Headache | ICD 10 | G43.909 | Migraine, unspecified, not intractable, without status migrainosus |
| Migraine and Headache | ICD 10 | G43.91 | Migraine, unspecified, intractable |
| Migraine and Headache | ICD 10 | G43.911 | Migraine, unspecified, intractable, with status migrainosus |
| Migraine and Headache | ICD 10 | G43.919 | Migraine, unspecified, intractable, without status migrainosus |
| Migraine and Headache | ICD 10 | G43.E | Chronic migraine with aura |
| Migraine and Headache | ICD 10 | G43.E0 | Chronic migraine with aura, not intractable |

| | | | |
|---|---|---|---|
| Migraine and Headache | ICD 10 | G43.E01 | Chronic migraine with aura, not intractable, with status migrainosus |
| Migraine and Headache | ICD 10 | G43.E09 | Chronic migraine with aura, not intractable, without status migrainosus |
| Migraine and Headache | ICD 10 | G43.E1 | Chronic migraine with aura, intractable |
| Migraine and Headache | ICD 10 | G43.E11 | Chronic migraine with aura, intractable, with status migrainosus |
| Migraine and Headache | ICD 10 | G43.E19 | Chronic migraine with aura, intractable, without status migrainosus |
| Migraine and Headache | ICD 10 | G44 | Other headache syndromes |
| Migraine and Headache | ICD 10 | G44.0 | Cluster headaches and other trigeminal autonomic cephalgias (TAC) |
| Migraine and Headache | ICD 10 | G44.00 | Cluster headache syndrome, unspecified |
| Migraine and Headache | ICD 10 | G44.001 | Cluster headache syndrome, intractable |
| Migraine and Headache | ICD 10 | G44.009 | Cluster headache syndrome, not intractable |
| Migraine and Headache | ICD 10 | G44.01 | Episodic cluster headache |
| Migraine and Headache | ICD 10 | G44.011 | Episodic cluster headache, intractable |
| Migraine and Headache | ICD 10 | G44.019 | Episodic cluster headache, not intractable |
| Migraine and Headache | ICD 10 | G44.02 | Chronic cluster headache |
| Migraine and Headache | ICD 10 | G44.021 | Chronic cluster headache, intractable |
| Migraine and Headache | ICD 10 | G44.029 | Chronic cluster headache, not intractable |

| | | | |
|---|---|---|---|
| Migraine and Headache | ICD 10 | G44.03 | Episodic paroxysmal hemicrania |
| Migraine and Headache | ICD 10 | G44.031 | Episodic paroxysmal hemicrania, intractable |
| Migraine and Headache | ICD 10 | G44.039 | Episodic paroxysmal hemicrania, not intractable |
| Migraine and Headache | ICD 10 | G44.04 | Chronic paroxysmal hemicrania |
| Migraine and Headache | ICD 10 | G44.041 | Chronic paroxysmal hemicrania, intractable |
| Migraine and Headache | ICD 10 | G44.049 | Chronic paroxysmal hemicrania, not intractable |
| Migraine and Headache | ICD 10 | G44.05 | Short-lasting unilateral neuralgiform headache with conjunctival injection and tearing (SUNCT) |
| Migraine and Headache | ICD 10 | G44.051 | SUNCT, intractable |
| Migraine and Headache | ICD 10 | G44.059 | SUNCT, not intractable |
| Migraine and Headache | ICD 10 | G44.09 | Other trigeminal autonomic cephalgias (TAC) |
| Migraine and Headache | ICD 10 | G44.091 | Other TAC, intractable |
| Migraine and Headache | ICD 10 | G44.099 | Other TAC, not intractable |
| Migraine and Headache | ICD 10 | G44.1 | Vascular headache, not elsewhere classified |
| Migraine and Headache | ICD 10 | G44.2 | Tension-type headache |
| Migraine and Headache | ICD 10 | G44.20 | Tension-type headache, unspecified |
| Migraine and Headache | ICD 10 | G44.201 | Tension-type headache, intractable |

| | | | |
|---|---|---|---|
| Migraine and Headache | ICD 10 | G44.209 | Tension-type headache, not intractable |
| Migraine and Headache | ICD 10 | G44.21 | Episodic tension-type headache |
| Migraine and Headache | ICD 10 | G44.211 | Episodic tension-type headache, intractable |
| Migraine and Headache | ICD 10 | G44.219 | Episodic tension-type headache, not intractable |
| Migraine and Headache | ICD 10 | G44.22 | Chronic tension-type headache |
| Migraine and Headache | ICD 10 | G44.221 | Chronic tension-type headache, intractable |
| Migraine and Headache | ICD 10 | G44.229 | Chronic tension-type headache, not intractable |
| Migraine and Headache | ICD 10 | G44.3 | Post-traumatic headache |
| Migraine and Headache | ICD 10 | G44.30 | Post-traumatic headache, unspecified |
| Migraine and Headache | ICD 10 | G44.301 | Post-traumatic headache, intractable |
| Migraine and Headache | ICD 10 | G44.309 | Post-traumatic headache, not intractable |
| Migraine and Headache | ICD 10 | G44.31 | Acute post-traumatic headache |
| Migraine and Headache | ICD 10 | G44.311 | Acute post-traumatic headache, intractable |
| Migraine and Headache | ICD 10 | G44.319 | Acute post-traumatic headache, not intractable |
| Migraine and Headache | ICD 10 | G44.32 | Chronic post-traumatic headache |
| Migraine and Headache | ICD 10 | G44.321 | Chronic post-traumatic headache, intractable |

| | | | |
|---|---|---|---|
| Migraine and Headache | ICD 10 | G44.329 | Chronic post-traumatic headache, not intractable |
| Migraine and Headache | ICD 10 | G44.4 | Drug-induced headache, not elsewhere classified |
| Migraine and Headache | ICD 10 | G44.40 | Drug-induced headache, not intractable |
| Migraine and Headache | ICD 10 | G44.41 | Drug-induced headache, intractable |
| Migraine and Headache | ICD 10 | G44.5 | Complicated headache syndromes |
| Migraine and Headache | ICD 10 | G44.51 | Hemicrania continua |
| Migraine and Headache | ICD 10 | G44.52 | New daily persistent headache (NDPH) |
| Migraine and Headache | ICD 10 | G44.53 | Primary thunderclap headache |
| Migraine and Headache | ICD 10 | G44.59 | Other complicated headache syndrome |
| Migraine and Headache | ICD 10 | G44.8 | Other specified headache syndromes |
| Migraine and Headache | ICD 10 | G44.81 | Hypnic headache |
| Migraine and Headache | ICD 10 | G44.82 | Headache associated with sexual activity |
| Migraine and Headache | ICD 10 | G44.83 | Primary cough headache |
| Migraine and Headache | ICD 10 | G44.84 | Primary exertional headache |
| Migraine and Headache | ICD 10 | G44.85 | Primary stabbing headache |
| Migraine and Headache | ICD 10 | G44.86 | Cervicogenic headache |

| | | | |
|---|---|---|---|
| Migraine and Headache | ICD 10 | G44.89 | Other headache syndrome |
| Migraine and Headache | ICD 9 | 346 | Migraine |
| Migraine and Headache | ICD 9 | 346.0 | Migraine with aura |
| Migraine and Headache | ICD 9 | 346.00 | Migraine with aura, without mention of intractable migraine, without mention of status migrainosus |
| Migraine and Headache | ICD 9 | 346.01 | Migraine with aura, with intractable migraine, so stated, without mention of status migrainosus |
| Migraine and Headache | ICD 9 | 346.02 | Migraine with aura, without mention of intractable migraine, with status migrainosus |
| Migraine and Headache | ICD 9 | 346.03 | Migraine with aura, with intractable migraine, so stated, with status migrainosus |
| Migraine and Headache | ICD 9 | 346.1 | Migraine without aura |
| Migraine and Headache | ICD 9 | 346.10 | Migraine without aura, without mention of intractable migraine, without mention of status migrainosus |
| Migraine and Headache | ICD 9 | 346.11 | Migraine without aura, with intractable migraine, so stated, without mention of status migrainosus |
| Migraine and Headache | ICD 9 | 346.12 | Migraine without aura, without mention of intractable migraine, with status migrainosus |
| Migraine and Headache | ICD 9 | 346.13 | Migraine without aura, with intractable migraine, so stated, with status migrainosus |
| Migraine and Headache | ICD 9 | 346.2 | Variants of migraine, not elsewhere classified |
| Migraine and Headache | ICD 9 | 346.20 | Variants of migraine, not elsewhere classified, without mention of intractable migraine, without mention of status migrainosus |
| Migraine and Headache | ICD 9 | 346.21 | Variants of migraine, not elsewhere classified, with intractable migraine, so stated, without mention of status migrainosus |

| | | | |
|---|---|---|---|
| Migraine and Headache | ICD 9 | 346.22 | Variants of migraine, not elsewhere classified, without mention of intractable migraine, with status migrainosus |
| Migraine and Headache | ICD 9 | 346.23 | Variants of migraine, not elsewhere classified, with intractable migraine, so stated, with status migrainosus |
| Migraine and Headache | ICD 9 | 346.3 | Hemiplegic migraine |
| Migraine and Headache | ICD 9 | 346.30 | Hemiplegic migraine, without mention of intractable migraine, without mention of status migrainosus |
| Migraine and Headache | ICD 9 | 346.31 | Hemiplegic migraine, with intractable migraine, so stated, without mention of status migrainosus |
| Migraine and Headache | ICD 9 | 346.32 | Hemiplegic migraine, without mention of intractable migraine, with status migrainosus |
| Migraine and Headache | ICD 9 | 346.33 | Hemiplegic migraine, with intractable migraine, so stated, with status migrainosus |
| Migraine and Headache | ICD 9 | 346.4 | Menstrual migraine |
| Migraine and Headache | ICD 9 | 346.40 | Menstrual migraine, without mention of intractable migraine, without mention of status migrainosus |
| Migraine and Headache | ICD 9 | 346.41 | Menstrual migraine, with intractable migraine, so stated, without mention of status migrainosus |
| Migraine and Headache | ICD 9 | 346.42 | Menstrual migraine, without mention of intractable migraine, with status migrainosus |
| Migraine and Headache | ICD 9 | 346.43 | Menstrual migraine, with intractable migraine, so stated, with status migrainosus |
| Migraine and Headache | ICD 9 | 346.5 | Persistent migraine aura without cerebral infarction |
| Migraine and Headache | ICD 9 | 346.50 | Persistent migraine aura without cerebral infarction, without mention of intractable migraine, without mention of status migrainosus |
| Migraine and Headache | ICD 9 | 346.51 | Persistent migraine aura without cerebral infarction, with intractable migraine, so stated, without mention of status migrainosus |

| | | | |
|---|---|---|---|
| Migraine and Headache | ICD 9 | 346.52 | Persistent migraine aura without cerebral infarction, without mention of intractable migraine, with status migrainosus |
| Migraine and Headache | ICD 9 | 346.53 | Persistent migraine aura without cerebral infarction, with intractable migraine, so stated, with status migrainosus |
| Migraine and Headache | ICD 9 | 346.6 | Persistent migraine aura with cerebral infarction |
| Migraine and Headache | ICD 9 | 346.60 | Persistent migraine aura with cerebral infarction, without mention of intractable migraine, without mention of status migrainosus |
| Migraine and Headache | ICD 9 | 346.61 | Persistent migraine aura with cerebral infarction, with intractable migraine, so stated, without mention of status migrainosus |
| Migraine and Headache | ICD 9 | 346.62 | Persistent migraine aura with cerebral infarction, without mention of intractable migraine, with status migrainosus |
| Migraine and Headache | ICD 9 | 346.63 | Persistent migraine aura with cerebral infarction, with intractable migraine, so stated, with status migrainosus |
| Migraine and Headache | ICD 9 | 346.7 | Chronic migraine without aura |
| Migraine and Headache | ICD 9 | 346.70 | Chronic migraine without aura, without mention of intractable migraine, without mention of status migrainosus |
| Migraine and Headache | ICD 9 | 346.71 | Chronic migraine without aura, with intractable migraine, so stated, without mention of status migrainosus |
| Migraine and Headache | ICD 9 | 346.72 | Chronic migraine without aura, without mention of intractable migraine, with status migrainosus |
| Migraine and Headache | ICD 9 | 346.73 | Chronic migraine without aura, with intractable migraine, so stated, with status migrainosus |
| Migraine and Headache | ICD 9 | 346.8 | Other forms of migraine |
| Migraine and Headache | ICD 9 | 346.80 | Other forms of migraine, without mention of intractable migraine, without mention of status migrainosus |
| Migraine and Headache | ICD 9 | 346.81 | Other forms of migraine, with intractable migraine, so stated, without mention of status migrainosus |

| | | | |
|---|---|---|---|
| Migraine and Headache | ICD 9 | 346.82 | Other forms of migraine, without mention of intractable migraine, with status migrainosus |
| Migraine and Headache | ICD 9 | 346.83 | Other forms of migraine, with intractable migraine, so stated, with status migrainosus |
| Migraine and Headache | ICD 9 | 346.9 | Migraine unspecified |
| Migraine and Headache | ICD 9 | 346.90 | Migraine, unspecified, without mention of intractable migraine, without mention of status migrainosus |
| Migraine and Headache | ICD 9 | 346.91 | Migraine, unspecified, with intractable migraine, so stated, without mention of status migrainosus |
| Migraine and Headache | ICD 9 | 346.92 | Migraine, unspecified, without mention of intractable migraine, with status migrainosus |
| Migraine and Headache | ICD 9 | 346.93 | Migraine, unspecified, with intractable migraine, so stated, with status migrainosus |
| Epilepsy | ICD 10 | G40 | Epilepsy and recurrent seizures |
| Epilepsy | ICD 10 | G40.0 | Localization-related (focal) (partial) idiopathic epilepsy and epileptic syndromes with seizures of localized onset |
| Epilepsy | ICD 10 | G40.00 | Localization-related (focal) (partial) idiopathic epilepsy and epileptic syndromes with seizures of localized onset, not intractable |
| Epilepsy | ICD 10 | G40.001 | Localization-related (focal) (partial) idiopathic epilepsy and epileptic syndromes with seizures of localized onset, with status epilepticus |
| Epilepsy | ICD 10 | G40.009 | Localization-related (focal) (partial) idiopathic epilepsy and epileptic syndromes with seizures of localized onset, without status epilepticus |
| Epilepsy | ICD 10 | G40.01 | Localization-related (focal) (partial) idiopathic epilepsy and epileptic syndromes with seizures of localized onset, intractable |
| Epilepsy | ICD 10 | G40.011 | Localization-related (focal) (partial) idiopathic epilepsy and epileptic syndromes with seizures of localized onset, intractable, with status epilepticus |

| | | | |
|---|---|---|---|
| Epilepsy | ICD 10 | G40.019 | Localization-related (focal) (partial) idiopathic epilepsy and epileptic syndromes with seizures of localized onset, intractable, without status epilepticus |
| Epilepsy | ICD 10 | G40.1 | Localization-related (focal) (partial) symptomatic epilepsy and epileptic syndromes with simple partial seizures |
| Epilepsy | ICD 10 | G40.10 | Localization-related (focal) (partial) symptomatic epilepsy and epileptic syndromes with simple partial seizures, not intractable |
| Epilepsy | ICD 10 | G40.101 | Localization-related (focal) (partial) symptomatic epilepsy and epileptic syndromes with simple partial seizures, not intractable, with status epilepticus |
| Epilepsy | ICD 10 | G40.109 | Localization-related (focal) (partial) symptomatic epilepsy and epileptic syndromes with simple partial seizures, not intractable, without status epilepticus |
| Epilepsy | ICD 10 | G40.11 | Localization-related (focal) (partial) symptomatic epilepsy and epileptic syndromes with simple partial seizures, intractable |
| Epilepsy | ICD 10 | G40.111 | Localization-related (focal) (partial) symptomatic epilepsy and epileptic syndromes with simple partial seizures, intractable, with status epilepticus |
| Epilepsy | ICD 10 | G40.119 | Localization-related (focal) (partial) symptomatic epilepsy and epileptic syndromes with simple partial seizures, intractable, without status epilepticus |
| Epilepsy | ICD 10 | G40.2 | Localization-related (focal) (partial) symptomatic epilepsy and epileptic syndromes with complex partial seizures |
| Epilepsy | ICD 10 | G40.20 | Localization-related (focal) (partial) symptomatic epilepsy and epileptic syndromes with complex partial seizures, not intractable |
| Epilepsy | ICD 10 | G40.201 | Localization-related (focal) (partial) symptomatic epilepsy and epileptic syndromes with complex partial seizures, not intractable, with status epilepticus |
| Epilepsy | ICD 10 | G40.209 | Localization-related (focal) (partial) symptomatic epilepsy and epileptic syndromes with complex partial seizures, not intractable, without status epilepticus |

| | | | |
|---|---|---|---|
| Epilepsy | ICD 10 | G40.21 | Localization-related (focal) (partial) symptomatic epilepsy and epileptic syndromes with complex partial seizures, intractable |
| Epilepsy | ICD 10 | G40.211 | Localization-related (focal) (partial) symptomatic epilepsy and epileptic syndromes with complex partial seizures, intractable, with status epilepticus |
| Epilepsy | ICD 10 | G40.219 | Localization-related (focal) (partial) symptomatic epilepsy and epileptic syndromes with complex partial seizures, intractable, without status epilepticus |
| Epilepsy | ICD 10 | G40.3 | Generalized idiopathic epilepsy and epileptic syndromes |
| Epilepsy | ICD 10 | G40.30 | Generalized idiopathic epilepsy and epileptic syndromes, not intractable |
| Epilepsy | ICD 10 | G40.301 | Generalized idiopathic epilepsy and epileptic syndromes, not intractable, with status epilepticus |
| Epilepsy | ICD 10 | G40.309 | Generalized idiopathic epilepsy and epileptic syndromes, not intractable, without status epilepticus |
| Epilepsy | ICD 10 | G40.31 | Generalized idiopathic epilepsy and epileptic syndromes, intractable |
| Epilepsy | ICD 10 | G40.311 | Generalized idiopathic epilepsy and epileptic syndromes, intractable, with status epilepticus |
| Epilepsy | ICD 10 | G40.319 | Generalized idiopathic epilepsy and epileptic syndromes, intractable, without status epilepticus |
| Epilepsy | ICD 10 | G40.A | Absence epileptic syndrome |
| Epilepsy | ICD 10 | G40.A0 | Absence epileptic syndrome, not intractable |
| Epilepsy | ICD 10 | G40.A01 | Absence epileptic syndrome, not intractable, with status epilepticus |
| Epilepsy | ICD 10 | G40.A09 | Absence epileptic syndrome, not intractable, without status epilepticus |
| Epilepsy | ICD 10 | G40.A1 | Absence epileptic syndrome, intractable |
| Epilepsy | ICD 10 | G40.A11 | Absence epileptic syndrome, intractable, with status epilepticus |

| | | | |
|---|---|---|---|
| Epilepsy | ICD 10 | G40.A19 | Absence epileptic syndrome, intractable, without status epilepticus |
| Epilepsy | ICD 10 | G40.B | Juvenile myoclonic epilepsy [impulsive petit mal] |
| Epilepsy | ICD 10 | G40.B0 | Juvenile myoclonic epilepsy, not intractable |
| Epilepsy | ICD 10 | G40.B01 | Juvenile myoclonic epilepsy, not intractable, with status epilepticus |
| Epilepsy | ICD 10 | G40.B09 | Juvenile myoclonic epilepsy, not intractable, without status epilepticus |
| Epilepsy | ICD 10 | G40.B1 | Juvenile myoclonic epilepsy, intractable |
| Epilepsy | ICD 10 | G40.B11 | Juvenile myoclonic epilepsy, intractable, with status epilepticus |
| Epilepsy | ICD 10 | G40.B19 | Juvenile myoclonic epilepsy, intractable, without status epilepticus |
| Epilepsy | ICD 10 | G40.C | Lafora progressive myoclonus epilepsy |
| Epilepsy | ICD 10 | G40.C0 | Lafora progressive myoclonus epilepsy, not intractable |
| Epilepsy | ICD 10 | G40.C01 | Lafora progressive myoclonus epilepsy, not intractable, with status epilepticus |
| Epilepsy | ICD 10 | G40.C09 | Lafora progressive myoclonus epilepsy, not intractable, without status epilepticus |
| Epilepsy | ICD 10 | G40.C1 | Lafora progressive myoclonus epilepsy, intractable |
| Epilepsy | ICD 10 | G40.C11 | Lafora progressive myoclonus epilepsy, intractable, with status epilepticus |
| Epilepsy | ICD 10 | G40.C19 | Lafora progressive myoclonus epilepsy, intractable, without status epilepticus |
| Epilepsy | ICD 10 | G40.4 | Other generalized epilepsy and epileptic syndromes |
| Epilepsy | ICD 10 | G40.40 | Other generalized epilepsy and epileptic syndromes, not intractable |
| Epilepsy | ICD 10 | G40.401 | Other generalized epilepsy and epileptic syndromes, not intractable, with status epilepticus |

| | | | |
|---|---|---|---|
| Epilepsy | ICD 10 | G40.409 | Other generalized epilepsy and epileptic syndromes, not intractable, without status epilepticus |
| Epilepsy | ICD 10 | G40.41 | Other generalized epilepsy and epileptic syndromes, intractable |
| Epilepsy | ICD 10 | G40.411 | Other generalized epilepsy and epileptic syndromes, intractable, with status epilepticus |
| Epilepsy | ICD 10 | G40.419 | Other generalized epilepsy and epileptic syndromes, intractable, without status epilepticus |
| Epilepsy | ICD 10 | G40.42 | Cyclin-Dependent Kinase-Like 5 Deficiency Disorder |
| Epilepsy | ICD 10 | G40.5 | Epileptic seizures related to external causes |
| Epilepsy | ICD 10 | G40.50 | Epileptic seizures related to external causes, not intractable |
| Epilepsy | ICD 10 | G40.501 | Epileptic seizures related to external causes, not intractable, with status epilepticus |
| Epilepsy | ICD 10 | G40.509 | Epileptic seizures related to external causes, not intractable, without status epilepticus |
| Epilepsy | ICD 10 | G40.8 | Other epilepsy and recurrent seizures |
| Epilepsy | ICD 10 | G40.80 | Other epilepsy |
| Epilepsy | ICD 10 | G40.801 | Other epilepsy, not intractable, with status epilepticus |
| Epilepsy | ICD 10 | G40.802 | Other epilepsy, not intractable, without status epilepticus |
| Epilepsy | ICD 10 | G40.803 | Other epilepsy, intractable, with status epilepticus |
| Epilepsy | ICD 10 | G40.804 | Other epilepsy, intractable, without status epilepticus |
| Epilepsy | ICD 10 | G40.81 | Lennox-Gastaut syndrome |
| Epilepsy | ICD 10 | G40.811 | Lennox-Gastaut syndrome, not intractable, with status epilepticus |
| Epilepsy | ICD 10 | G40.812 | Lennox-Gastaut syndrome, not intractable, without status epilepticus |
| Epilepsy | ICD 10 | G40.813 | Lennox-Gastaut syndrome, intractable, with status epilepticus |

| | | | |
|---|---|---|---|
| Epilepsy | ICD 10 | G40.814 | Lennox-Gastaut syndrome, intractable, without status epilepticus |
| Epilepsy | ICD 10 | G40.82 | Epileptic spasms |
| Epilepsy | ICD 10 | G40.821 | Epileptic spasms, not intractable, with status epilepticus |
| Epilepsy | ICD 10 | G40.822 | Epileptic spasms, not intractable, without status epilepticus |
| Epilepsy | ICD 10 | G40.823 | Epileptic spasms, intractable, with status epilepticus |
| Epilepsy | ICD 10 | G40.824 | Epileptic spasms, intractable, without status epilepticus |
| Epilepsy | ICD 10 | G40.83 | Dravet syndrome |
| Epilepsy | ICD 10 | G40.833 | Dravet syndrome, intractable, with status epilepticus |
| Epilepsy | ICD 10 | G40.834 | Dravet syndrome, intractable, without status epilepticus |
| Epilepsy | ICD 10 | G40.84 | KCNQ2-related epilepsy |
| Epilepsy | ICD 10 | G40.841 | KCNQ2-related epilepsy, not intractable, with status epilepticus |
| Epilepsy | ICD 10 | G40.842 | KCNQ2-related epilepsy, not intractable, without status epilepticus |
| Epilepsy | ICD 10 | G40.843 | KCNQ2-related epilepsy, intractable, with status epilepticus |
| Epilepsy | ICD 10 | G40.844 | KCNQ2-related epilepsy, intractable, without status epilepticus |
| Epilepsy | ICD 10 | G40.89 | Other seizures |
| Epilepsy | ICD 10 | G40.9 | Epilepsy, unspecified |
| Epilepsy | ICD 10 | G40.90 | Epilepsy, unspecified, not intractable |
| Epilepsy | ICD 10 | G40.901 | Epilepsy, unspecified, not intractable, with status epilepticus |
| Epilepsy | ICD 10 | G40.909 | Epilepsy, unspecified, not intractable, without status epilepticus |
| Epilepsy | ICD 10 | G40.91 | Epilepsy, unspecified, intractable |
| Epilepsy | ICD 10 | G40.911 | Epilepsy, unspecified, intractable, with status epilepticus |
| Epilepsy | ICD 10 | G40.919 | Epilepsy, unspecified, intractable, without status epilepticus |

| | | | |
|---|---|---|---|
| Epilepsy | ICD 9 | 345 | Epilepsy and recurrent seizures |
| Epilepsy | ICD 9 | 345.0 | Generalized nonconvulsive epilepsy |
| Epilepsy | ICD 9 | 345.00 | Generalized nonconvulsive epilepsy, without mention of intractable epilepsy |
| Epilepsy | ICD 9 | 345.01 | Generalized nonconvulsive epilepsy, with intractable epilepsy |
| Epilepsy | ICD 9 | 345.1 | Generalized convulsive epilepsy |
| Epilepsy | ICD 9 | 345.10 | Generalized convulsive epilepsy, without mention of intractable epilepsy |
| Epilepsy | ICD 9 | 345.11 | Generalized convulsive epilepsy, with intractable epilepsy |
| Epilepsy | ICD 9 | 345.2 | Petit mal status |
| Epilepsy | ICD 9 | 345.3 | Grand mal status |
| Epilepsy | ICD 9 | 345.4 | Localization-related (focal) (partial) epilepsy and epileptic syndromes with complex partial seizures |
| Epilepsy | ICD 9 | 345.40 | Localization-related (focal) (partial) epilepsy and epileptic syndromes with complex partial seizures, without mention of intractable epilepsy |
| Epilepsy | ICD 9 | 345.41 | Localization-related (focal) (partial) epilepsy and epileptic syndromes with complex partial seizures, with intractable epilepsy |
| Epilepsy | ICD 9 | 345.5 | Localization-related (focal) (partial) epilepsy and epileptic syndromes with simple partial seizures |
| Epilepsy | ICD 9 | 345.50 | Localization-related (focal) (partial) epilepsy and epileptic syndromes with simple partial seizures, without mention of intractable epilepsy |
| Epilepsy | ICD 9 | 345.51 | Localization-related (focal) (partial) epilepsy and epileptic syndromes with simple partial seizures, with intractable epilepsy |
| Epilepsy | ICD 9 | 345.6 | Infantile spasms |
| Epilepsy | ICD 9 | 345.60 | Infantile spasms, without mention of intractable epilepsy |
| Epilepsy | ICD 9 | 345.61 | Infantile spasms, with intractable epilepsy |

| | | | |
|---|---|---|---|
| Epilepsy | ICD 9 | 345.7 | Epilepsia partialis continua |
| Epilepsy | ICD 9 | 345.70 | Epilepsia partialis continua, without mention of intractable epilepsy |
| Epilepsy | ICD 9 | 345.71 | Epilepsia partialis continua, with intractable epilepsy |
| Epilepsy | ICD 9 | 345.8 | Other forms of epilepsy and recurrent seizures |
| Epilepsy | ICD 9 | 345.80 | Other forms of epilepsy and recurrent seizures, without mention of intractable epilepsy |
| Epilepsy | ICD 9 | 345.81 | Other forms of epilepsy and recurrent seizures, with intractable epilepsy |
| Epilepsy | ICD 9 | 345.9 | Epilepsy unspecified |
| Epilepsy | ICD 9 | 345.90 | Epilepsy, unspecified, without mention of intractable epilepsy |
| Epilepsy | ICD 9 | 345.91 | Epilepsy, unspecified, with intractable epilepsy |
| Dizziness and giddiness | ICD 10 | R42 | Dizziness and giddiness |
| Dizziness and giddiness | ICD 9 | 780.4 | Dizziness and giddiness |
| Brain Traumatic Injury | ICD 10 | S06.2 | Diffuse traumatic brain injury |
| Brain Traumatic Injury | ICD 10 | S06.2X | Diffuse traumatic brain injury |
| Brain Traumatic Injury | ICD 10 | S06.2X0 | Diffuse traumatic brain injury without loss of consciousness |
| Brain Traumatic Injury | ICD 10 | S06.2X0A | Diffuse traumatic brain injury without loss of consciousness, initial encounter |
| Brain Traumatic Injury | ICD 10 | S06.2X0D | Diffuse traumatic brain injury without loss of consciousness, subsequent encounter |
| Brain Traumatic Injury | ICD 10 | S06.2X0S | Diffuse traumatic brain injury without loss of consciousness, sequela |
| Brain Traumatic Injury | ICD 10 | S06.2X1 | Diffuse traumatic brain injury with loss of consciousness of 30 minutes or less |

| | | | |
|---|---|---|---|
| Brain Traumatic Injury | ICD 10 | S06.2X1A | Diffuse traumatic brain injury with loss of consciousness of 30 minutes or less, initial encounter |
| Brain Traumatic Injury | ICD 10 | S06.2X1D | Diffuse traumatic brain injury with loss of consciousness of 30 minutes or less, subsequent encounter |
| Brain Traumatic Injury | ICD 10 | S06.2X1S | Diffuse traumatic brain injury with loss of consciousness of 30 minutes or less, sequela |
| Brain Traumatic Injury | ICD 10 | S06.2X2 | Diffuse traumatic brain injury with loss of consciousness of 31 minutes to 59 minutes |
| Brain Traumatic Injury | ICD 10 | S06.2X2A | Diffuse traumatic brain injury with loss of consciousness of 31 minutes to 59 minutes, initial encounter |
| Brain Traumatic Injury | ICD 10 | S06.2X2D | Diffuse traumatic brain injury with loss of consciousness of 31 minutes to 59 minutes, subsequent encounter |
| Brain Traumatic Injury | ICD 10 | S06.2X2S | Diffuse traumatic brain injury with loss of consciousness of 31 minutes to 59 minutes, sequela |
| Brain Traumatic Injury | ICD 10 | S06.2X3 | Diffuse traumatic brain injury with loss of consciousness of 1 hour to 5 hours 59 minutes |
| Brain Traumatic Injury | ICD 10 | S06.2X3A | Diffuse traumatic brain injury with loss of consciousness of 1 hour to 5 hours 59 minutes, initial encounter |
| Brain Traumatic Injury | ICD 10 | S06.2X3D | Diffuse traumatic brain injury with loss of consciousness of 1 hour to 5 hours 59 minutes, subsequent encounter |
| Brain Traumatic Injury | ICD 10 | S06.2X3S | Diffuse traumatic brain injury with loss of consciousness of 1 hour to 5 hours 59 minutes, sequela |
| Brain Traumatic Injury | ICD 10 | S06.2X4 | Diffuse traumatic brain injury with loss of consciousness of 6 hours to 24 hours |
| Brain Traumatic Injury | ICD 10 | S06.2X4A | Diffuse traumatic brain injury with loss of consciousness of 6 hours to 24 hours, initial encounter |
| Brain Traumatic Injury | ICD 10 | S06.2X4D | Diffuse traumatic brain injury with loss of consciousness of 6 hours to 24 hours, subsequent encounter |
| Brain Traumatic Injury | ICD 10 | S06.2X4S | Diffuse traumatic brain injury with loss of consciousness of 6 hours to 24 hours, sequela |
| Brain Traumatic Injury | ICD 10 | S06.2X5 | Diffuse traumatic brain injury with loss of consciousness greater than 24 hours with return to pre-existing conscious levels |

| | | | |
|---|---|---|---|
| Brain Traumatic Injury | ICD 10 | S06.2X5A | Diffuse traumatic brain injury with loss of consciousness greater than 24 hours, initial encounter |
| Brain Traumatic Injury | ICD 10 | S06.2X5D | Diffuse traumatic brain injury with loss of consciousness greater than 24 hours, subsequent encounter |
| Brain Traumatic Injury | ICD 10 | S06.2X5S | Diffuse traumatic brain injury with loss of consciousness greater than 24 hours, sequela |
| Brain Traumatic Injury | ICD 10 | S06.2X6 | Diffuse traumatic brain injury with loss of consciousness greater than 24 hours without return to pre-existing conscious level with patient surviving |
| Brain Traumatic Injury | ICD 10 | S06.2X6A | Diffuse traumatic brain injury with loss of consciousness greater than 24 hours without return to pre-existing conscious level, initial encounter |
| Brain Traumatic Injury | ICD 10 | S06.2X6D | Diffuse traumatic brain injury with loss of consciousness greater than 24 hours without return to pre-existing conscious level, subsequent encounter |
| Brain Traumatic Injury | ICD 10 | S06.2X6S | Diffuse traumatic brain injury with loss of consciousness greater than 24 hours without return to pre-existing conscious level, sequela |
| Brain Traumatic Injury | ICD 10 | S06.2X7 | Diffuse traumatic brain injury with loss of consciousness of any duration with death due to brain injury prior to regaining consciousness |
| Brain Traumatic Injury | ICD 10 | S06.2X7A | Diffuse traumatic brain injury with loss of consciousness of any duration with death due to brain injury prior to regaining consciousness, initial encounter |
| Brain Traumatic Injury | ICD 10 | S06.2X8 | Diffuse traumatic brain injury with loss of consciousness of any duration with death due to other cause prior to regaining consciousness |
| Brain Traumatic Injury | ICD 10 | S06.2X8A | Diffuse traumatic brain injury with loss of consciousness of any duration with death due to other cause prior to regaining consciousness, initial encounter |
| Brain Traumatic Injury | ICD 10 | S06.2XA | Diffuse traumatic brain injury with loss of consciousness status unknown |
| Brain Traumatic Injury | ICD 10 | S06.2XAA | Diffuse traumatic brain injury with loss of consciousness status unknown, initial encounter |

| | | | |
|---|---|---|---|
| Brain Traumatic Injury | ICD 10 | S06.2XAD | Diffuse traumatic brain injury with loss of consciousness status unknown, subsequent encounter |
| Brain Traumatic Injury | ICD 10 | S06.2XAS | Diffuse traumatic brain injury with loss of consciousness status unknown, sequela |
| Brain Traumatic Injury | ICD 10 | S06.2X9 | Diffuse traumatic brain injury with loss of consciousness of unspecified duration |
| Brain Traumatic Injury | ICD 10 | S06.2X9A | Diffuse traumatic brain injury with loss of consciousness of unspecified duration, initial encounter |
| Brain Traumatic Injury | ICD 10 | S06.2X9D | Diffuse traumatic brain injury with loss of consciousness of unspecified duration, subsequent encounter |
| Brain Traumatic Injury | ICD 10 | S06.2X9S | Diffuse traumatic brain injury with loss of consciousness of unspecified duration, sequela |
| Brain Traumatic Injury | ICD 10 | S06.3 | Focal traumatic brain injury |
| Brain Traumatic Injury | ICD 10 | S06.30 | Unspecified focal traumatic brain injury |
| Brain Traumatic Injury | ICD 10 | S06.300 | Unspecified focal traumatic brain injury without loss of consciousness |
| Brain Traumatic Injury | ICD 10 | S06.300A | Unspecified focal traumatic brain injury without loss of consciousness, initial encounter |
| Brain Traumatic Injury | ICD 10 | S06.300D | Unspecified focal traumatic brain injury without loss of consciousness, subsequent encounter |
| Brain Traumatic Injury | ICD 10 | S06.300S | Unspecified focal traumatic brain injury without loss of consciousness, sequela |
| Brain Traumatic Injury | ICD 10 | S06.301 | Unspecified focal traumatic brain injury with loss of consciousness of 30 minutes or less |
| Brain Traumatic Injury | ICD 10 | S06.301A | Unspecified focal traumatic brain injury with loss of consciousness of 30 minutes or less, initial encounter |
| Brain Traumatic Injury | ICD 10 | S06.301D | Unspecified focal traumatic brain injury with loss of consciousness of 30 minutes or less, subsequent encounter |
| Brain Traumatic Injury | ICD 10 | S06.301S | Unspecified focal traumatic brain injury with loss of consciousness of 30 minutes or less, sequela |

| | | | |
|---|---|---|---|
| Brain Traumatic Injury | ICD 10 | S06.302 | Unspecified focal traumatic brain injury with loss of consciousness of 31 minutes to 59 minutes |
| Brain Traumatic Injury | ICD 10 | S06.302A | Unspecified focal traumatic brain injury with loss of consciousness of 31 minutes to 59 minutes, initial encounter |
| Brain Traumatic Injury | ICD 10 | S06.302D | Unspecified focal traumatic brain injury with loss of consciousness of 31 minutes to 59 minutes, subsequent encounter |
| Brain Traumatic Injury | ICD 10 | S06.302S | Unspecified focal traumatic brain injury with loss of consciousness of 31 minutes to 59 minutes, sequela |
| Brain Traumatic Injury | ICD 10 | S06.303 | Unspecified focal traumatic brain injury with loss of consciousness of 1 hour to 5 hours 59 minutes |
| Brain Traumatic Injury | ICD 10 | S06.303A | Unspecified focal traumatic brain injury with loss of consciousness of 1 hour to 5 hours 59 minutes, initial encounter |
| Brain Traumatic Injury | ICD 10 | S06.303D | Unspecified focal traumatic brain injury with loss of consciousness of 1 hour to 5 hours 59 minutes, subsequent encounter |
| Brain Traumatic Injury | ICD 10 | S06.303S | Unspecified focal traumatic brain injury with loss of consciousness of 1 hour to 5 hours 59 minutes, sequela |
| Brain Traumatic Injury | ICD 10 | S06.304 | Unspecified focal traumatic brain injury with loss of consciousness of 6 hours to 24 hours |
| Brain Traumatic Injury | ICD 10 | S06.304A | Unspecified focal traumatic brain injury with loss of consciousness of 6 hours to 24 hours, initial encounter |
| Brain Traumatic Injury | ICD 10 | S06.304D | Unspecified focal traumatic brain injury with loss of consciousness of 6 hours to 24 hours, subsequent encounter |
| Brain Traumatic Injury | ICD 10 | S06.304S | Unspecified focal traumatic brain injury with loss of consciousness of 6 hours to 24 hours, sequela |
| Brain Traumatic Injury | ICD 10 | S06.305 | Unspecified focal traumatic brain injury with loss of consciousness greater than 24 hours with return to pre-existing conscious level |
| Brain Traumatic Injury | ICD 10 | S06.305A | Unspecified focal traumatic brain injury with loss of consciousness greater than 24 hours with return to pre-existing conscious level, initial encounter |

| | | | |
|---|---|---|---|
| Brain Traumatic Injury | ICD 10 | S06.305D | Unspecified focal traumatic brain injury with loss of consciousness greater than 24 hours with return to pre-existing conscious level, subsequent encounter |
| Brain Traumatic Injury | ICD 10 | S06.305S | Unspecified focal traumatic brain injury with loss of consciousness greater than 24 hours with return to pre-existing conscious level, sequela |
| Brain Traumatic Injury | ICD 10 | S06.306 | Unspecified focal traumatic brain injury with loss of consciousness greater than 24 hours without return to pre-existing conscious level with patient surviving |
| Brain Traumatic Injury | ICD 10 | S06.306A | Unspecified focal traumatic brain injury with loss of consciousness greater than 24 hours without return to pre-existing conscious level with patient surviving, initial encounter |
| Brain Traumatic Injury | ICD 10 | S06.306D | Unspecified focal traumatic brain injury with loss of consciousness greater than 24 hours without return to pre-existing conscious level with patient surviving, subsequent encounter |
| Brain Traumatic Injury | ICD 10 | S06.306S | Unspecified focal traumatic brain injury with loss of consciousness greater than 24 hours without return to pre-existing conscious level with patient surviving, sequela |
| Brain Traumatic Injury | ICD 10 | S06.307 | Unspecified focal traumatic brain injury with loss of consciousness of any duration with death due to brain injury prior to regaining consciousness |
| Brain Traumatic Injury | ICD 10 | S06.307A | Unspecified focal traumatic brain injury with loss of consciousness of any duration with death due to brain injury prior to regaining consciousness, initial encounter |
| Brain Traumatic Injury | ICD 10 | S06.308 | Unspecified focal traumatic brain injury with loss of consciousness of any duration with death due to other cause prior to regaining consciousness |
| Brain Traumatic Injury | ICD 10 | S06.308A | Unspecified focal traumatic brain injury with loss of consciousness of any duration with death due to other cause prior to regaining consciousness, initial encounter |
| Brain Traumatic Injury | ICD 10 | S06.30A | Unspecified focal traumatic brain injury with loss of consciousness status unknown |
| Brain Traumatic Injury | ICD 10 | S06.30AA | Unspecified focal traumatic brain injury with loss of consciousness status unknown, initial encounter |

| | | | |
|---|---|---|---|
| Brain Traumatic Injury | ICD 10 | S06.30AD | Unspecified focal traumatic brain injury with loss of consciousness status unknown, subsequent encounter |
| Brain Traumatic Injury | ICD 10 | S06.30AS | Unspecified focal traumatic brain injury with loss of consciousness status unknown, sequela |
| Brain Traumatic Injury | ICD 10 | S06.309 | Unspecified focal traumatic brain injury with loss of consciousness of unspecified duration |
| Brain Traumatic Injury | ICD 10 | S06.309A | Unspecified focal traumatic brain injury with loss of consciousness of unspecified duration, initial encounter |
| Brain Traumatic Injury | ICD 10 | S06.309D | Unspecified focal traumatic brain injury with loss of consciousness of unspecified duration, subsequent encounter |
| Brain Traumatic Injury | ICD 10 | S06.309S | Unspecified focal traumatic brain injury with loss of consciousness of unspecified duration, sequela |
| Brain Traumatic Injury | ICD 10 | S06.31 | Contusion and laceration of right cerebrum |
| Brain Traumatic Injury | ICD 10 | S06.310 | Contusion and laceration of right cerebrum without loss of consciousness |
| Brain Traumatic Injury | ICD 10 | S06.310A | Contusion and laceration of right cerebrum without loss of consciousness, initial encounter |
| Brain Traumatic Injury | ICD 10 | S06.310D | Contusion and laceration of right cerebrum without loss of consciousness, subsequent encounter |
| Brain Traumatic Injury | ICD 10 | S06.310S | Contusion and laceration of right cerebrum without loss of consciousness, sequela |
| Brain Traumatic Injury | ICD 10 | S06.311 | Contusion and laceration of right cerebrum with loss of consciousness of 30 minutes or less |
| Brain Traumatic Injury | ICD 10 | S06.311A | Contusion and laceration of right cerebrum with loss of consciousness of 30 minutes or less, initial encounter |
| Brain Traumatic Injury | ICD 10 | S06.311D | Contusion and laceration of right cerebrum with loss of consciousness of 30 minutes or less, subsequent encounter |
| Brain Traumatic Injury | ICD 10 | S06.311S | Contusion and laceration of right cerebrum with loss of consciousness of 30 minutes or less, sequela |
| Brain Traumatic Injury | ICD 10 | S06.312 | Contusion and laceration of right cerebrum with loss of consciousness of 31 minutes to 59 minutes |

| | | | |
|---|---|---|---|
| Brain Traumatic Injury | ICD 10 | S06.312A | Contusion and laceration of right cerebrum with loss of consciousness of 31 minutes to 59 minutes, initial encounter |
| Brain Traumatic Injury | ICD 10 | S06.312D | Contusion and laceration of right cerebrum with loss of consciousness of 31 minutes to 59 minutes, subsequent encounter |
| Brain Traumatic Injury | ICD 10 | S06.312S | Contusion and laceration of right cerebrum with loss of consciousness of 31 minutes to 59 minutes, sequela |
| Brain Traumatic Injury | ICD 10 | S06.313 | Contusion and laceration of right cerebrum with loss of consciousness of 1 hour to 5 hours 59 minutes |
| Brain Traumatic Injury | ICD 10 | S06.313A | Contusion and laceration of right cerebrum with loss of consciousness of 1 hour to 5 hours 59 minutes, initial encounter |
| Brain Traumatic Injury | ICD 10 | S06.313D | Contusion and laceration of right cerebrum with loss of consciousness of 1 hour to 5 hours 59 minutes, subsequent encounter |
| Brain Traumatic Injury | ICD 10 | S06.313S | Contusion and laceration of right cerebrum with loss of consciousness of 1 hour to 5 hours 59 minutes, sequela |
| Brain Traumatic Injury | ICD 10 | S06.314 | Contusion and laceration of right cerebrum with loss of consciousness of 6 hours to 24 hours |
| Brain Traumatic Injury | ICD 10 | S06.314A | Contusion and laceration of right cerebrum with loss of consciousness of 6 hours to 24 hours, initial encounter |
| Brain Traumatic Injury | ICD 10 | S06.314D | Contusion and laceration of right cerebrum with loss of consciousness of 6 hours to 24 hours, subsequent encounter |
| Brain Traumatic Injury | ICD 10 | S06.314S | Contusion and laceration of right cerebrum with loss of consciousness of 6 hours to 24 hours, sequela |
| Brain Traumatic Injury | ICD 10 | S06.315 | Contusion and laceration of right cerebrum with loss of consciousness greater than 24 hours with return to pre-existing conscious level |
| Brain Traumatic Injury | ICD 10 | S06.315A | Contusion and laceration of right cerebrum with loss of consciousness greater than 24 hours with return to pre-existing conscious level, initial encounter |
| Brain Traumatic Injury | ICD 10 | S06.315D | Contusion and laceration of right cerebrum with loss of consciousness greater than 24 hours with return to pre-existing conscious level, subsequent encounter |

| | | | |
|---|---|---|---|
| Brain Traumatic Injury | ICD 10 | S06.315S | Contusion and laceration of right cerebrum with loss of consciousness greater than 24 hours with return to pre-existing conscious level, sequela |
| Brain Traumatic Injury | ICD 10 | S06.316 | Contusion and laceration of right cerebrum with loss of consciousness greater than 24 hours without return to pre-existing conscious level with patient surviving |
| Brain Traumatic Injury | ICD 10 | S06.316A | Contusion and laceration of right cerebrum with loss of consciousness greater than 24 hours without return to pre-existing conscious level with patient surviving, initial encounter |
| Brain Traumatic Injury | ICD 10 | S06.316D | Contusion and laceration of right cerebrum with loss of consciousness greater than 24 hours without return to pre-existing conscious level with patient surviving, subsequent encounter |
| Brain Traumatic Injury | ICD 10 | S06.316S | Contusion and laceration of right cerebrum with loss of consciousness greater than 24 hours without return to pre-existing conscious level with patient surviving, sequela |
| Brain Traumatic Injury | ICD 10 | S06.317 | Contusion and laceration of right cerebrum with loss of consciousness of any duration with death due to brain injury prior to regaining consciousness |
| Brain Traumatic Injury | ICD 10 | S06.317A | Contusion and laceration of right cerebrum with loss of consciousness of any duration with death due to brain injury prior to regaining consciousness, initial encounter |
| Brain Traumatic Injury | ICD 10 | S06.318 | Contusion and laceration of right cerebrum with loss of consciousness of any duration with death due to other cause prior to regaining consciousness |
| Brain Traumatic Injury | ICD 10 | S06.318A | Contusion and laceration of right cerebrum with loss of consciousness of any duration with death due to other cause prior to regaining consciousness, initial encounter |
| Brain Traumatic Injury | ICD 10 | S06.31A | Contusion and laceration of right cerebrum with loss of consciousness status unknown |
| Brain Traumatic Injury | ICD 10 | S06.31AA | Contusion and laceration of right cerebrum with loss of consciousness status unknown, initial encounter |
| Brain Traumatic Injury | ICD 10 | S06.31AD | Contusion and laceration of right cerebrum with loss of consciousness status unknown, subsequent encounter |

| | | | |
|---|---|---|---|
| Brain Traumatic Injury | ICD 10 | S06.31AS | Contusion and laceration of right cerebrum with loss of consciousness status unknown, sequela |
| Brain Traumatic Injury | ICD 10 | S06.319 | Contusion and laceration of right cerebrum with loss of consciousness of unspecified duration |
| Brain Traumatic Injury | ICD 10 | S06.319A | Contusion and laceration of right cerebrum with loss of consciousness of unspecified duration, initial encounter |
| Brain Traumatic Injury | ICD 10 | S06.319D | Contusion and laceration of right cerebrum with loss of consciousness of unspecified duration, subsequent encounter |
| Brain Traumatic Injury | ICD 10 | S06.319S | Contusion and laceration of right cerebrum with loss of consciousness of unspecified duration, sequela |
| Brain Traumatic Injury | ICD 10 | S06.32 | Contusion and laceration of left cerebrum |
| Brain Traumatic Injury | ICD 10 | S06.320 | Contusion and laceration of left cerebrum without loss of consciousness |
| Brain Traumatic Injury | ICD 10 | S06.320A | Contusion and laceration of left cerebrum without loss of consciousness, initial encounter |
| Brain Traumatic Injury | ICD 10 | S06.320D | Contusion and laceration of left cerebrum without loss of consciousness, subsequent encounter |
| Brain Traumatic Injury | ICD 10 | S06.320S | Contusion and laceration of left cerebrum without loss of consciousness, sequela |
| Brain Traumatic Injury | ICD 10 | S06.321 | Contusion and laceration of left cerebrum with loss of consciousness of 30 minutes or less |
| Brain Traumatic Injury | ICD 10 | S06.321A | Contusion and laceration of left cerebrum with loss of consciousness of 30 minutes or less, initial encounter |
| Brain Traumatic Injury | ICD 10 | S06.321D | Contusion and laceration of left cerebrum with loss of consciousness of 30 minutes or less, subsequent encounter |
| Brain Traumatic Injury | ICD 10 | S06.321S | Contusion and laceration of left cerebrum with loss of consciousness of 30 minutes or less, sequela |
| Brain Traumatic Injury | ICD 10 | S06.322 | Contusion and laceration of left cerebrum with loss of consciousness of 31 minutes to 59 minutes |
| Brain Traumatic Injury | ICD 10 | S06.322A | Contusion and laceration of left cerebrum with loss of consciousness of 31 minutes to 59 minutes, initial encounter |

| | | | |
|---|---|---|---|
| Brain Traumatic Injury | ICD 10 | S06.322D | Contusion and laceration of left cerebrum with loss of consciousness of 31 minutes to 59 minutes, subsequent encounter |
| Brain Traumatic Injury | ICD 10 | S06.322S | Contusion and laceration of left cerebrum with loss of consciousness of 31 minutes to 59 minutes, sequela |
| Brain Traumatic Injury | ICD 10 | S06.323 | Contusion and laceration of left cerebrum with loss of consciousness of 1 hour to 5 hours 59 minutes |
| Brain Traumatic Injury | ICD 10 | S06.323A | Contusion and laceration of left cerebrum with loss of consciousness of 1 hour to 5 hours 59 minutes, initial encounter |
| Brain Traumatic Injury | ICD 10 | S06.323D | Contusion and laceration of left cerebrum with loss of consciousness of 1 hour to 5 hours 59 minutes, subsequent encounter |
| Brain Traumatic Injury | ICD 10 | S06.323S | Contusion and laceration of left cerebrum with loss of consciousness of 1 hour to 5 hours 59 minutes, sequela |
| Brain Traumatic Injury | ICD 10 | S06.324 | Contusion and laceration of left cerebrum with loss of consciousness of 6 hours to 24 hours |
| Brain Traumatic Injury | ICD 10 | S06.324A | Contusion and laceration of left cerebrum with loss of consciousness of 6 hours to 24 hours, initial encounter |
| Brain Traumatic Injury | ICD 10 | S06.324D | Contusion and laceration of left cerebrum with loss of consciousness of 6 hours to 24 hours, subsequent encounter |
| Brain Traumatic Injury | ICD 10 | S06.324S | Contusion and laceration of left cerebrum with loss of consciousness of 6 hours to 24 hours, sequela |
| Brain Traumatic Injury | ICD 10 | S06.325 | Contusion and laceration of left cerebrum with loss of consciousness greater than 24 hours with return to pre-existing conscious level |
| Brain Traumatic Injury | ICD 10 | S06.325A | Contusion and laceration of left cerebrum with loss of consciousness greater than 24 hours with return to pre-existing conscious level, initial encounter |
| Brain Traumatic Injury | ICD 10 | S06.325D | Contusion and laceration of left cerebrum with loss of consciousness greater than 24 hours with return to pre-existing conscious level, subsequent encounter |
| Brain Traumatic Injury | ICD 10 | S06.325S | Contusion and laceration of left cerebrum with loss of consciousness greater than 24 hours with return to pre-existing conscious level, sequela |

| | | | |
|---|---|---|---|
| Brain Traumatic Injury | ICD 10 | S06.326 | Contusion and laceration of left cerebrum with loss of consciousness greater than 24 hours without return to pre-existing conscious level with patient surviving |
| Brain Traumatic Injury | ICD 10 | S06.326A | Contusion and laceration of left cerebrum with loss of consciousness greater than 24 hours without return to pre-existing conscious level with patient surviving, initial encounter |
| Brain Traumatic Injury | ICD 10 | S06.326D | Contusion and laceration of left cerebrum with loss of consciousness greater than 24 hours without return to pre-existing conscious level with patient surviving, subsequent encounter |
| Brain Traumatic Injury | ICD 10 | S06.326S | Contusion and laceration of left cerebrum with loss of consciousness greater than 24 hours without return to pre-existing conscious level with patient surviving, sequela |
| Brain Traumatic Injury | ICD 10 | S06.327 | Contusion and laceration of left cerebrum with loss of consciousness of any duration with death due to brain injury prior to regaining consciousness |
| Brain Traumatic Injury | ICD 10 | S06.327A | Contusion and laceration of left cerebrum with loss of consciousness of any duration with death due to brain injury prior to regaining consciousness, initial encounter |
| Brain Traumatic Injury | ICD 10 | S06.328 | Contusion and laceration of left cerebrum with loss of consciousness of any duration with death due to other cause prior to regaining consciousness |
| Brain Traumatic Injury | ICD 10 | S06.328A | Contusion and laceration of left cerebrum with loss of consciousness of any duration with death due to other cause prior to regaining consciousness, initial encounter |
| Brain Traumatic Injury | ICD 10 | S06.32A | Contusion and laceration of left cerebrum with loss of consciousness status unknown |
| Brain Traumatic Injury | ICD 10 | S06.32AA | Contusion and laceration of left cerebrum with loss of consciousness status unknown, initial encounter |
| Brain Traumatic Injury | ICD 10 | S06.32AD | Contusion and laceration of left cerebrum with loss of consciousness status unknown, subsequent encounter |
| Brain Traumatic Injury | ICD 10 | S06.32AS | Contusion and laceration of left cerebrum with loss of consciousness status unknown, sequela |
| Brain Traumatic Injury | ICD 10 | S06.329 | Contusion and laceration of left cerebrum with loss of consciousness of unspecified duration |

| | | | |
|---|---|---|---|
| Brain Traumatic Injury | ICD 10 | S06.329A | Contusion and laceration of left cerebrum with loss of consciousness of unspecified duration, initial encounter |
| Brain Traumatic Injury | ICD 10 | S06.329D | Contusion and laceration of left cerebrum with loss of consciousness of unspecified duration, subsequent encounter |
| Brain Traumatic Injury | ICD 10 | S06.329S | Contusion and laceration of left cerebrum with loss of consciousness of unspecified duration, sequela |
| Brain Traumatic Injury | ICD 10 | S06.33 | Contusion and laceration of cerebrum, unspecified |
| Brain Traumatic Injury | ICD 10 | S06.330 | Contusion and laceration of cerebrum, unspecified, without loss of consciousness |
| Brain Traumatic Injury | ICD 10 | S06.330A | Contusion and laceration of cerebrum, unspecified, without loss of consciousness, initial encounter |
| Brain Traumatic Injury | ICD 10 | S06.330D | Contusion and laceration of cerebrum, unspecified, without loss of consciousness, subsequent encounter |
| Brain Traumatic Injury | ICD 10 | S06.330S | Contusion and laceration of cerebrum, unspecified, without loss of consciousness, sequela |
| Brain Traumatic Injury | ICD 10 | S06.331 | Contusion and laceration of cerebrum, unspecified, with loss of consciousness of 30 minutes or less |
| Brain Traumatic Injury | ICD 10 | S06.331A | Contusion and laceration of cerebrum, unspecified, with loss of consciousness of 30 minutes or less, initial encounter |
| Brain Traumatic Injury | ICD 10 | S06.331D | Contusion and laceration of cerebrum, unspecified, with loss of consciousness of 30 minutes or less, subsequent encounter |
| Brain Traumatic Injury | ICD 10 | S06.331S | Contusion and laceration of cerebrum, unspecified, with loss of consciousness of 30 minutes or less, sequela |
| Brain Traumatic Injury | ICD 10 | S06.332 | Contusion and laceration of cerebrum, unspecified, with loss of consciousness of 31 minutes to 59 minutes |
| Brain Traumatic Injury | ICD 10 | S06.332A | Contusion and laceration of cerebrum, unspecified, with loss of consciousness of 31 minutes to 59 minutes, initial encounter |
| Brain Traumatic Injury | ICD 10 | S06.332D | Contusion and laceration of cerebrum, unspecified, with loss of consciousness of 31 minutes to 59 minutes, subsequent encounter |

| | | | |
|---|---|---|---|
| Brain Traumatic Injury | ICD 10 | S06.332S | Contusion and laceration of cerebrum, unspecified, with loss of consciousness of 31 minutes to 59 minutes, sequela |
| Brain Traumatic Injury | ICD 10 | S06.333 | Contusion and laceration of cerebrum, unspecified, with loss of consciousness of 1 hour to 5 hours 59 minutes |
| Brain Traumatic Injury | ICD 10 | S06.333A | Contusion and laceration of cerebrum, unspecified, with loss of consciousness of 1 hour to 5 hours 59 minutes, initial encounter |
| Brain Traumatic Injury | ICD 10 | S06.333D | Contusion and laceration of cerebrum, unspecified, with loss of consciousness of 1 hour to 5 hours 59 minutes, subsequent encounter |
| Brain Traumatic Injury | ICD 10 | S06.333S | Contusion and laceration of cerebrum, unspecified, with loss of consciousness of 1 hour to 5 hours 59 minutes, sequela |
| Brain Traumatic Injury | ICD 10 | S06.334 | Contusion and laceration of cerebrum, unspecified, with loss of consciousness of 6 hours to 24 hours |
| Brain Traumatic Injury | ICD 10 | S06.334A | Contusion and laceration of cerebrum, unspecified, with loss of consciousness of 6 hours to 24 hours, initial encounter |
| Brain Traumatic Injury | ICD 10 | S06.334D | Contusion and laceration of cerebrum, unspecified, with loss of consciousness of 6 hours to 24 hours, subsequent encounter |
| Brain Traumatic Injury | ICD 10 | S06.334S | Contusion and laceration of cerebrum, unspecified, with loss of consciousness of 6 hours to 24 hours, sequela |
| Brain Traumatic Injury | ICD 10 | S06.335 | Contusion and laceration of cerebrum, unspecified, with loss of consciousness greater than 24 hours with return to pre-existing conscious level |
| Brain Traumatic Injury | ICD 10 | S06.335A | Contusion and laceration of cerebrum, unspecified, with loss of consciousness greater than 24 hours with return to pre-existing conscious level, initial encounter |
| Brain Traumatic Injury | ICD 10 | S06.335D | Contusion and laceration of cerebrum, unspecified, with loss of consciousness greater than 24 hours with return to pre-existing conscious level, subsequent encounter |
| Brain Traumatic Injury | ICD 10 | S06.335S | Contusion and laceration of cerebrum, unspecified, with loss of consciousness greater than 24 hours with return to pre-existing conscious level, sequela |

| | | | |
|---|---|---|---|
| Brain Traumatic Injury | ICD 10 | S06.336 | Contusion and laceration of cerebrum, unspecified, with loss of consciousness greater than 24 hours without return to pre-existing conscious level with patient surviving |
| Brain Traumatic Injury | ICD 10 | S06.336A | Contusion and laceration of cerebrum, unspecified, with loss of consciousness greater than 24 hours without return to pre-existing conscious level with patient surviving, initial encounter |
| Brain Traumatic Injury | ICD 10 | S06.336D | Contusion and laceration of cerebrum, unspecified, with loss of consciousness greater than 24 hours without return to pre-existing conscious level with patient surviving, subsequent encounter |
| Brain Traumatic Injury | ICD 10 | S06.336S | Contusion and laceration of cerebrum, unspecified, with loss of consciousness greater than 24 hours without return to pre-existing conscious level with patient surviving, sequela |
| Brain Traumatic Injury | ICD 10 | S06.337 | Contusion and laceration of cerebrum, unspecified, with loss of consciousness of any duration with death due to brain injury prior to regaining consciousness |
| Brain Traumatic Injury | ICD 10 | S06.337A | Contusion and laceration of cerebrum, unspecified, with loss of consciousness of any duration with death due to brain injury prior to regaining consciousness, initial encounter |
| Brain Traumatic Injury | ICD 10 | S06.338 | Contusion and laceration of cerebrum, unspecified, with loss of consciousness of any duration with death due to other cause prior to regaining consciousness |
| Brain Traumatic Injury | ICD 10 | S06.338A | Contusion and laceration of cerebrum, unspecified, with loss of consciousness of any duration with death due to other cause prior to regaining consciousness, initial encounter |
| Brain Traumatic Injury | ICD 10 | S06.33A | Contusion and laceration of cerebrum, unspecified, with loss of consciousness status unknown |
| Brain Traumatic Injury | ICD 10 | S06.33AA | Contusion and laceration of cerebrum, unspecified, with loss of consciousness status unknown, initial encounter |
| Brain Traumatic Injury | ICD 10 | S06.33AD | Contusion and laceration of cerebrum, unspecified, with loss of consciousness status unknown, subsequent encounter |
| Brain Traumatic Injury | ICD 10 | S06.33AS | Contusion and laceration of cerebrum, unspecified, with loss of consciousness status unknown, sequela |
| Brain Traumatic Injury | ICD 10 | S06.339 | Contusion and laceration of cerebrum, unspecified, with loss of consciousness of unspecified duration |

| | | | |
|---|---|---|---|
| Brain Traumatic Injury | ICD 10 | S06.339A | Contusion and laceration of cerebrum, unspecified, with loss of consciousness of unspecified duration, initial encounter |
| Brain Traumatic Injury | ICD 10 | S06.339D | Contusion and laceration of cerebrum, unspecified, with loss of consciousness of unspecified duration, subsequent encounter |
| Brain Traumatic Injury | ICD 10 | S06.339S | Contusion and laceration of cerebrum, unspecified, with loss of consciousness of unspecified duration, sequela |
| Brain Traumatic Injury | ICD 10 | S06.34 | Traumatic hemorrhage of right cerebrum |
| Brain Traumatic Injury | ICD 10 | S06.340 | Traumatic hemorrhage of right cerebrum without loss of consciousness |
| Brain Traumatic Injury | ICD 10 | S06.340A | Traumatic hemorrhage of right cerebrum without loss of consciousness, initial encounter |
| Brain Traumatic Injury | ICD 10 | S06.340D | Traumatic hemorrhage of right cerebrum without loss of consciousness, subsequent encounter |
| Brain Traumatic Injury | ICD 10 | S06.340S | Traumatic hemorrhage of right cerebrum without loss of consciousness, sequela |
| Brain Traumatic Injury | ICD 10 | S06.341 | Traumatic hemorrhage of right cerebrum with loss of consciousness of 30 minutes or less |
| Brain Traumatic Injury | ICD 10 | S06.341A | Traumatic hemorrhage of right cerebrum with loss of consciousness of 30 minutes or less, initial encounter |
| Brain Traumatic Injury | ICD 10 | S06.341D | Traumatic hemorrhage of right cerebrum with loss of consciousness of 30 minutes or less, subsequent encounter |
| Brain Traumatic Injury | ICD 10 | S06.341S | Traumatic hemorrhage of right cerebrum with loss of consciousness of 30 minutes or less, sequela |
| Brain Traumatic Injury | ICD 10 | S06.342 | Traumatic hemorrhage of right cerebrum with loss of consciousness of 31 minutes to 59 minutes |
| Brain Traumatic Injury | ICD 10 | S06.342A | Traumatic hemorrhage of right cerebrum with loss of consciousness of 31 minutes to 59 minutes, initial encounter |
| Brain Traumatic Injury | ICD 10 | S06.342D | Traumatic hemorrhage of right cerebrum with loss of consciousness of 31 minutes to 59 minutes, subsequent encounter |

| | | | |
|---|---|---|---|
| Brain Traumatic Injury | ICD 10 | S06.342S | Traumatic hemorrhage of right cerebrum with loss of consciousness of 31 minutes to 59 minutes, sequela |
| Brain Traumatic Injury | ICD 10 | S06.343 | Traumatic hemorrhage of right cerebrum with loss of consciousness of 1 hour to 5 hours 59 minutes |
| Brain Traumatic Injury | ICD 10 | S06.343A | Traumatic hemorrhage of right cerebrum with loss of consciousness of 1 hour to 5 hours 59 minutes, initial encounter |
| Brain Traumatic Injury | ICD 10 | S06.343D | Traumatic hemorrhage of right cerebrum with loss of consciousness of 1 hour to 5 hours 59 minutes, subsequent encounter |
| Brain Traumatic Injury | ICD 10 | S06.343S | Traumatic hemorrhage of right cerebrum with loss of consciousness of 1 hour to 5 hours 59 minutes, sequela |
| Brain Traumatic Injury | ICD 10 | S06.344 | Traumatic hemorrhage of right cerebrum with loss of consciousness of 6 hours to 24 hours |
| Brain Traumatic Injury | ICD 10 | S06.344A | Traumatic hemorrhage of right cerebrum with loss of consciousness of 6 hours to 24 hours, initial encounter |
| Brain Traumatic Injury | ICD 10 | S06.344D | Traumatic hemorrhage of right cerebrum with loss of consciousness of 6 hours to 24 hours, subsequent encounter |
| Brain Traumatic Injury | ICD 10 | S06.344S | Traumatic hemorrhage of right cerebrum with loss of consciousness of 6 hours to 24 hours, sequela |
| Brain Traumatic Injury | ICD 10 | S06.345 | Traumatic hemorrhage of right cerebrum with loss of consciousness greater than 24 hours with return to pre-existing conscious level |
| Brain Traumatic Injury | ICD 10 | S06.345A | Traumatic hemorrhage of right cerebrum with loss of consciousness greater than 24 hours with return to pre-existing conscious level, initial encounter |
| Brain Traumatic Injury | ICD 10 | S06.345D | Traumatic hemorrhage of right cerebrum with loss of consciousness greater than 24 hours with return to pre-existing conscious level, subsequent encounter |
| Brain Traumatic Injury | ICD 10 | S06.345S | Traumatic hemorrhage of right cerebrum with loss of consciousness greater than 24 hours with return to pre-existing conscious level, sequela |
| Brain Traumatic Injury | ICD 10 | S06.346 | Traumatic hemorrhage of right cerebrum with loss of consciousness greater than 24 hours without return to pre-existing conscious level with patient surviving |

| | | | |
|---|---|---|---|
| Brain Traumatic Injury | ICD 10 | S06.346A | Traumatic hemorrhage of right cerebrum with loss of consciousness greater than 24 hours without return to pre-existing conscious level with patient surviving, initial encounter |
| Brain Traumatic Injury | ICD 10 | S06.346D | Traumatic hemorrhage of right cerebrum with loss of consciousness greater than 24 hours without return to pre-existing conscious level with patient surviving, subsequent encounter |
| Brain Traumatic Injury | ICD 10 | S06.346S | Traumatic hemorrhage of right cerebrum with loss of consciousness greater than 24 hours without return to pre-existing conscious level with patient surviving, sequela |
| Brain Traumatic Injury | ICD 10 | S06.347 | Traumatic hemorrhage of right cerebrum with loss of consciousness of any duration with death due to brain injury prior to regaining consciousness |
| Brain Traumatic Injury | ICD 10 | S06.347A | Traumatic hemorrhage of right cerebrum with loss of consciousness of any duration with death due to brain injury prior to regaining consciousness, initial encounter |
| Brain Traumatic Injury | ICD 10 | S06.348 | Traumatic hemorrhage of right cerebrum with loss of consciousness of any duration with death due to other cause prior to regaining consciousness |
| Brain Traumatic Injury | ICD 10 | S06.348A | Traumatic hemorrhage of right cerebrum with loss of consciousness of any duration with death due to other cause prior to regaining consciousness, initial encounter |
| Brain Traumatic Injury | ICD 10 | S06.34A | Traumatic hemorrhage of right cerebrum with loss of consciousness status unknown |
| Brain Traumatic Injury | ICD 10 | S06.34AA | Traumatic hemorrhage of right cerebrum with loss of consciousness status unknown, initial encounter |
| Brain Traumatic Injury | ICD 10 | S06.34AD | Traumatic hemorrhage of right cerebrum with loss of consciousness status unknown, subsequent encounter |
| Brain Traumatic Injury | ICD 10 | S06.34AS | Traumatic hemorrhage of right cerebrum with loss of consciousness status unknown, sequela |
| Brain Traumatic Injury | ICD 10 | S06.349 | Traumatic hemorrhage of right cerebrum with loss of consciousness of unspecified duration |
| Brain Traumatic Injury | ICD 10 | S06.349A | Traumatic hemorrhage of right cerebrum with loss of consciousness of unspecified duration, initial encounter |

| | | | |
|---|---|---|---|
| Brain Traumatic Injury | ICD 10 | S06.349D | Traumatic hemorrhage of right cerebrum with loss of consciousness of unspecified duration, subsequent encounter |
| Brain Traumatic Injury | ICD 10 | S06.349S | Traumatic hemorrhage of right cerebrum with loss of consciousness of unspecified duration, sequela |
| Brain Traumatic Injury | ICD 10 | S06.35 | Traumatic hemorrhage of left cerebrum |
| Brain Traumatic Injury | ICD 10 | S06.350 | Traumatic hemorrhage of left cerebrum without loss of consciousness |
| Brain Traumatic Injury | ICD 10 | S06.350A | Traumatic hemorrhage of left cerebrum without loss of consciousness, initial encounter |
| Brain Traumatic Injury | ICD 10 | S06.350D | Traumatic hemorrhage of left cerebrum without loss of consciousness, subsequent encounter |
| Brain Traumatic Injury | ICD 10 | S06.350S | Traumatic hemorrhage of left cerebrum without loss of consciousness, sequela |
| Brain Traumatic Injury | ICD 10 | S06.351 | Traumatic hemorrhage of left cerebrum with loss of consciousness of 30 minutes or less |
| Brain Traumatic Injury | ICD 10 | S06.351A | Traumatic hemorrhage of left cerebrum with loss of consciousness of 30 minutes or less, initial encounter |
| Brain Traumatic Injury | ICD 10 | S06.351D | Traumatic hemorrhage of left cerebrum with loss of consciousness of 30 minutes or less, subsequent encounter |
| Brain Traumatic Injury | ICD 10 | S06.351S | Traumatic hemorrhage of left cerebrum with loss of consciousness of 30 minutes or less, sequela |
| Brain Traumatic Injury | ICD 10 | S06.352 | Traumatic hemorrhage of left cerebrum with loss of consciousness of 31 minutes to 59 minutes |
| Brain Traumatic Injury | ICD 10 | S06.352A | Traumatic hemorrhage of left cerebrum with loss of consciousness of 31 minutes to 59 minutes, initial encounter |
| Brain Traumatic Injury | ICD 10 | S06.352D | Traumatic hemorrhage of left cerebrum with loss of consciousness of 31 minutes to 59 minutes, subsequent encounter |
| Brain Traumatic Injury | ICD 10 | S06.352S | Traumatic hemorrhage of left cerebrum with loss of consciousness of 31 minutes to 59 minutes, sequela |

| | | | |
|---|---|---|---|
| Brain Traumatic Injury | ICD 10 | S06.353 | Traumatic hemorrhage of left cerebrum with loss of consciousness of 1 hour to 5 hours 59 minutes |
| Brain Traumatic Injury | ICD 10 | S06.353A | Traumatic hemorrhage of left cerebrum with loss of consciousness of 1 hour to 5 hours 59 minutes, initial encounter |
| Brain Traumatic Injury | ICD 10 | S06.353D | Traumatic hemorrhage of left cerebrum with loss of consciousness of 1 hour to 5 hours 59 minutes, subsequent encounter |
| Brain Traumatic Injury | ICD 10 | S06.353S | Traumatic hemorrhage of left cerebrum with loss of consciousness of 1 hour to 5 hours 59 minutes, sequela |
| Brain Traumatic Injury | ICD 10 | S06.354 | Traumatic hemorrhage of left cerebrum with loss of consciousness of 6 hours to 24 hours |
| Brain Traumatic Injury | ICD 10 | S06.354A | Traumatic hemorrhage of left cerebrum with loss of consciousness of 6 hours to 24 hours, initial encounter |
| Brain Traumatic Injury | ICD 10 | S06.354D | Traumatic hemorrhage of left cerebrum with loss of consciousness of 6 hours to 24 hours, subsequent encounter |
| Brain Traumatic Injury | ICD 10 | S06.354S | Traumatic hemorrhage of left cerebrum with loss of consciousness of 6 hours to 24 hours, sequela |
| Brain Traumatic Injury | ICD 10 | S06.355 | Traumatic hemorrhage of left cerebrum with loss of consciousness greater than 24 hours with return to pre-existing conscious level |
| Brain Traumatic Injury | ICD 10 | S06.355A | Traumatic hemorrhage of left cerebrum with loss of consciousness greater than 24 hours with return to pre-existing conscious level, initial encounter |
| Brain Traumatic Injury | ICD 10 | S06.355D | Traumatic hemorrhage of left cerebrum with loss of consciousness greater than 24 hours with return to pre-existing conscious level, subsequent encounter |
| Brain Traumatic Injury | ICD 10 | S06.355S | Traumatic hemorrhage of left cerebrum with loss of consciousness greater than 24 hours with return to pre-existing conscious level, sequela |
| Brain Traumatic Injury | ICD 10 | S06.356 | Traumatic hemorrhage of left cerebrum with loss of consciousness greater than 24 hours without return to pre-existing conscious level with patient surviving |
| Brain Traumatic Injury | ICD 10 | S06.356A | Traumatic hemorrhage of left cerebrum with loss of consciousness greater than 24 hours without return to pre- |

| | | | |
|---|---|---|---|
| | | | existing conscious level with patient surviving, initial encounter |
| Brain Traumatic Injury | ICD 10 | S06.356D | Traumatic hemorrhage of left cerebrum with loss of consciousness greater than 24 hours without return to pre-existing conscious level with patient surviving, subsequent encounter |
| Brain Traumatic Injury | ICD 10 | S06.356S | Traumatic hemorrhage of left cerebrum with loss of consciousness greater than 24 hours without return to pre-existing conscious level with patient surviving, sequela |
| Brain Traumatic Injury | ICD 10 | S06.357 | Traumatic hemorrhage of left cerebrum with loss of consciousness of any duration with death due to brain injury prior to regaining consciousness |
| Brain Traumatic Injury | ICD 10 | S06.357A | Traumatic hemorrhage of left cerebrum with loss of consciousness of any duration with death due to brain injury prior to regaining consciousness, initial encounter |
| Brain Traumatic Injury | ICD 10 | S06.358 | Traumatic hemorrhage of left cerebrum with loss of consciousness of any duration with death due to other cause prior to regaining consciousness |
| Brain Traumatic Injury | ICD 10 | S06.358A | Traumatic hemorrhage of left cerebrum with loss of consciousness of any duration with death due to other cause prior to regaining consciousness, initial encounter |
| Brain Traumatic Injury | ICD 10 | S06.35A | Traumatic hemorrhage of left cerebrum with loss of consciousness status unknown |
| Brain Traumatic Injury | ICD 10 | S06.35AA | Traumatic hemorrhage of left cerebrum with loss of consciousness status unknown, initial encounter |
| Brain Traumatic Injury | ICD 10 | S06.35AD | Traumatic hemorrhage of left cerebrum with loss of consciousness status unknown, subsequent encounter |
| Brain Traumatic Injury | ICD 10 | S06.35AS | Traumatic hemorrhage of left cerebrum with loss of consciousness status unknown, sequela |
| Brain Traumatic Injury | ICD 10 | S06.359 | Traumatic hemorrhage of left cerebrum with loss of consciousness of unspecified duration |
| Brain Traumatic Injury | ICD 10 | S06.359A | Traumatic hemorrhage of left cerebrum with loss of consciousness of unspecified duration, initial encounter |

| | | | |
|---|---|---|---|
| Brain Traumatic Injury | ICD 10 | S06.359D | Traumatic hemorrhage of left cerebrum with loss of consciousness of unspecified duration, subsequent encounter |
| Brain Traumatic Injury | ICD 10 | S06.359S | Traumatic hemorrhage of left cerebrum with loss of consciousness of unspecified duration, sequela |
| Brain Traumatic Injury | ICD 10 | S06.36 | Traumatic hemorrhage of cerebrum, unspecified |
| Brain Traumatic Injury | ICD 10 | S06.360 | Traumatic hemorrhage of cerebrum, unspecified, without loss of consciousness |
| Brain Traumatic Injury | ICD 10 | S06.360A | Traumatic hemorrhage of cerebrum, unspecified, without loss of consciousness, initial encounter |
| Brain Traumatic Injury | ICD 10 | S06.360D | Traumatic hemorrhage of cerebrum, unspecified, without loss of consciousness, subsequent encounter |
| Brain Traumatic Injury | ICD 10 | S06.360S | Traumatic hemorrhage of cerebrum, unspecified, without loss of consciousness, sequela |
| Brain Traumatic Injury | ICD 10 | S06.361 | Traumatic hemorrhage of cerebrum, unspecified, with loss of consciousness of 30 minutes or less |
| Brain Traumatic Injury | ICD 10 | S06.361A | Traumatic hemorrhage of cerebrum, unspecified, with loss of consciousness of 30 minutes or less, initial encounter |
| Brain Traumatic Injury | ICD 10 | S06.361D | Traumatic hemorrhage of cerebrum, unspecified, with loss of consciousness of 30 minutes or less, subsequent encounter |
| Brain Traumatic Injury | ICD 10 | S06.361S | Traumatic hemorrhage of cerebrum, unspecified, with loss of consciousness of 30 minutes or less, sequela |
| Brain Traumatic Injury | ICD 10 | S06.362 | Traumatic hemorrhage of cerebrum, unspecified, with loss of consciousness of 31 minutes to 59 minutes |
| Brain Traumatic Injury | ICD 10 | S06.362A | Traumatic hemorrhage of cerebrum, unspecified, with loss of consciousness of 31 minutes to 59 minutes, initial encounter |
| Brain Traumatic Injury | ICD 10 | S06.362D | Traumatic hemorrhage of cerebrum, unspecified, with loss of consciousness of 31 minutes to 59 minutes, subsequent encounter |
| Brain Traumatic Injury | ICD 10 | S06.362S | Traumatic hemorrhage of cerebrum, unspecified, with loss of consciousness of 31 minutes to 59 minutes, sequela |

| | | | |
|---|---|---|---|
| Brain Traumatic Injury | ICD 10 | S06.363 | Traumatic hemorrhage of cerebrum, unspecified, with loss of consciousness of 1 hour to 5 hours 59 minutes |
| Brain Traumatic Injury | ICD 10 | S06.363A | Traumatic hemorrhage of cerebrum, unspecified, with loss of consciousness of 1 hour to 5 hours 59 minutes, initial encounter |
| Brain Traumatic Injury | ICD 10 | S06.363D | Traumatic hemorrhage of cerebrum, unspecified, with loss of consciousness of 1 hour to 5 hours 59 minutes, subsequent encounter |
| Brain Traumatic Injury | ICD 10 | S06.363S | Traumatic hemorrhage of cerebrum, unspecified, with loss of consciousness of 1 hour to 5 hours 59 minutes, sequela |
| Brain Traumatic Injury | ICD 10 | S06.364 | Traumatic hemorrhage of cerebrum, unspecified, with loss of consciousness of 6 hours to 24 hours |
| Brain Traumatic Injury | ICD 10 | S06.364A | Traumatic hemorrhage of cerebrum, unspecified, with loss of consciousness of 6 hours to 24 hours, initial encounter |
| Brain Traumatic Injury | ICD 10 | S06.364D | Traumatic hemorrhage of cerebrum, unspecified, with loss of consciousness of 6 hours to 24 hours, subsequent encounter |
| Brain Traumatic Injury | ICD 10 | S06.364S | Traumatic hemorrhage of cerebrum, unspecified, with loss of consciousness of 6 hours to 24 hours, sequela |
| Brain Traumatic Injury | ICD 10 | S06.365 | Traumatic hemorrhage of cerebrum, unspecified, with loss of consciousness greater than 24 hours with return to pre-existing conscious level |
| Brain Traumatic Injury | ICD 10 | S06.365A | Traumatic hemorrhage of cerebrum, unspecified, with loss of consciousness greater than 24 hours with return to pre-existing conscious level, initial encounter |
| Brain Traumatic Injury | ICD 10 | S06.365D | Traumatic hemorrhage of cerebrum, unspecified, with loss of consciousness greater than 24 hours with return to pre-existing conscious level, subsequent encounter |
| Brain Traumatic Injury | ICD 10 | S06.365S | Traumatic hemorrhage of cerebrum, unspecified, with loss of consciousness greater than 24 hours with return to pre-existing conscious level, sequela |
| Brain Traumatic Injury | ICD 10 | S06.366 | Traumatic hemorrhage of cerebrum, unspecified, with loss of consciousness greater than 24 hours without return to pre-existing conscious level with patient surviving |
| Brain Traumatic Injury | ICD 10 | S06.366A | Traumatic hemorrhage of cerebrum, unspecified, with loss of consciousness greater than 24 hours without return to pre- |

| | | | |
|---|---|---|---|
| | | | existing conscious level with patient surviving, initial encounter |
| Brain Traumatic Injury | ICD 10 | S06.366D | Traumatic hemorrhage of cerebrum, unspecified, with loss of consciousness greater than 24 hours without return to pre-existing conscious level with patient surviving, subsequent encounter |
| Brain Traumatic Injury | ICD 10 | S06.366S | Traumatic hemorrhage of cerebrum, unspecified, with loss of consciousness greater than 24 hours without return to pre-existing conscious level with patient surviving, sequela |
| Brain Traumatic Injury | ICD 10 | S06.367 | Traumatic hemorrhage of cerebrum, unspecified, with loss of consciousness of any duration with death due to brain injury prior to regaining consciousness |
| Brain Traumatic Injury | ICD 10 | S06.367A | Traumatic hemorrhage of cerebrum, unspecified, with loss of consciousness of any duration with death due to brain injury prior to regaining consciousness, initial encounter |
| Brain Traumatic Injury | ICD 10 | S06.368 | Traumatic hemorrhage of cerebrum, unspecified, with loss of consciousness of any duration with death due to other cause prior to regaining consciousness |
| Brain Traumatic Injury | ICD 10 | S06.368A | Traumatic hemorrhage of cerebrum, unspecified, with loss of consciousness of any duration with death due to other cause prior to regaining consciousness, initial encounter |
| Brain Traumatic Injury | ICD 10 | S06.36A | Traumatic hemorrhage of cerebrum, unspecified, with loss of consciousness status unknown |
| Brain Traumatic Injury | ICD 10 | S06.36AA | Traumatic hemorrhage of cerebrum, unspecified, with loss of consciousness status unknown, initial encounter |
| Brain Traumatic Injury | ICD 10 | S06.36AD | Traumatic hemorrhage of cerebrum, unspecified, with loss of consciousness status unknown, subsequent encounter |
| Brain Traumatic Injury | ICD 10 | S06.36AS | Traumatic hemorrhage of cerebrum, unspecified, with loss of consciousness status unknown, sequela |
| Brain Traumatic Injury | ICD 10 | S06.369 | Traumatic hemorrhage of cerebrum, unspecified, with loss of consciousness of unspecified duration |
| Brain Traumatic Injury | ICD 10 | S06.369A | Traumatic hemorrhage of cerebrum, unspecified, with loss of consciousness of unspecified duration, initial encounter |

| | | | |
|---|---|---|---|
| Brain Traumatic Injury | ICD 10 | S06.369D | Traumatic hemorrhage of cerebrum, unspecified, with loss of consciousness of unspecified duration, subsequent encounter |
| Brain Traumatic Injury | ICD 10 | S06.369S | Traumatic hemorrhage of cerebrum, unspecified, with loss of consciousness of unspecified duration, sequela |
| Brain Traumatic Injury | ICD 10 | S06.37 | Contusion, laceration, and hemorrhage of cerebellum |
| Brain Traumatic Injury | ICD 10 | S06.370 | Contusion, laceration, and hemorrhage of cerebellum without loss of consciousness |
| Brain Traumatic Injury | ICD 10 | S06.370A | Contusion, laceration, and hemorrhage of cerebellum without loss of consciousness, initial encounter |
| Brain Traumatic Injury | ICD 10 | S06.370D | Contusion, laceration, and hemorrhage of cerebellum without loss of consciousness, subsequent encounter |
| Brain Traumatic Injury | ICD 10 | S06.370S | Contusion, laceration, and hemorrhage of cerebellum without loss of consciousness, sequela |
| Brain Traumatic Injury | ICD 10 | S06.371 | Contusion, laceration, and hemorrhage of cerebellum with loss of consciousness of 30 minutes or less |
| Brain Traumatic Injury | ICD 10 | S06.371A | Contusion, laceration, and hemorrhage of cerebellum with loss of consciousness of 30 minutes or less, initial encounter |
| Brain Traumatic Injury | ICD 10 | S06.371D | Contusion, laceration, and hemorrhage of cerebellum with loss of consciousness of 30 minutes or less, subsequent encounter |
| Brain Traumatic Injury | ICD 10 | S06.371S | Contusion, laceration, and hemorrhage of cerebellum with loss of consciousness of 30 minutes or less, sequela |
| Brain Traumatic Injury | ICD 10 | S06.372 | Contusion, laceration, and hemorrhage of cerebellum with loss of consciousness of 31 minutes to 59 minutes |
| Brain Traumatic Injury | ICD 10 | S06.372A | Contusion, laceration, and hemorrhage of cerebellum with loss of consciousness of 31 minutes to 59 minutes, initial encounter |
| Brain Traumatic Injury | ICD 10 | S06.372D | Contusion, laceration, and hemorrhage of cerebellum with loss of consciousness of 31 minutes to 59 minutes, subsequent encounter |
| Brain Traumatic Injury | ICD 10 | S06.372S | Contusion, laceration, and hemorrhage of cerebellum with loss of consciousness of 31 minutes to 59 minutes, sequela |

| | | | |
|---|---|---|---|
| Brain Traumatic Injury | ICD 10 | S06.373 | Contusion, laceration, and hemorrhage of cerebellum with loss of consciousness of 1 hour to 5 hours 59 minutes |
| Brain Traumatic Injury | ICD 10 | S06.373A | Contusion, laceration, and hemorrhage of cerebellum with loss of consciousness of 1 hour to 5 hours 59 minutes, initial encounter |
| Brain Traumatic Injury | ICD 10 | S06.373D | Contusion, laceration, and hemorrhage of cerebellum with loss of consciousness of 1 hour to 5 hours 59 minutes, subsequent encounter |
| Brain Traumatic Injury | ICD 10 | S06.373S | Contusion, laceration, and hemorrhage of cerebellum with loss of consciousness of 1 hour to 5 hours 59 minutes, sequela |
| Brain Traumatic Injury | ICD 10 | S06.374 | Contusion, laceration, and hemorrhage of cerebellum with loss of consciousness of 6 hours to 24 hours |
| Brain Traumatic Injury | ICD 10 | S06.374A | Contusion, laceration, and hemorrhage of cerebellum with loss of consciousness of 6 hours to 24 hours, initial encounter |
| Brain Traumatic Injury | ICD 10 | S06.374D | Contusion, laceration, and hemorrhage of cerebellum with loss of consciousness of 6 hours to 24 hours, subsequent encounter |
| Brain Traumatic Injury | ICD 10 | S06.374S | Contusion, laceration, and hemorrhage of cerebellum with loss of consciousness of 6 hours to 24 hours, sequela |
| Brain Traumatic Injury | ICD 10 | S06.375 | Contusion, laceration, and hemorrhage of cerebellum with loss of consciousness greater than 24 hours with return to pre-existing conscious level |
| Brain Traumatic Injury | ICD 10 | S06.375A | Contusion, laceration, and hemorrhage of cerebellum with loss of consciousness greater than 24 hours with return to pre-existing conscious level, initial encounter |
| Brain Traumatic Injury | ICD 10 | S06.375D | Contusion, laceration, and hemorrhage of cerebellum with loss of consciousness greater than 24 hours with return to pre-existing conscious level, subsequent encounter |
| Brain Traumatic Injury | ICD 10 | S06.375S | Contusion, laceration, and hemorrhage of cerebellum with loss of consciousness greater than 24 hours with return to pre-existing conscious level, sequela |
| Brain Traumatic Injury | ICD 10 | S06.376 | Contusion, laceration, and hemorrhage of cerebellum with loss of consciousness greater than 24 hours without return to pre-existing conscious level with patient surviving |

| | | | |
|---|---|---|---|
| Brain Traumatic Injury | ICD 10 | S06.376A | Contusion, laceration, and hemorrhage of cerebellum with loss of consciousness greater than 24 hours without return to pre-existing conscious level with patient surviving, initial encounter |
| Brain Traumatic Injury | ICD 10 | S06.376D | Contusion, laceration, and hemorrhage of cerebellum with loss of consciousness greater than 24 hours without return to pre-existing conscious level with patient surviving, subsequent encounter |
| Brain Traumatic Injury | ICD 10 | S06.376S | Contusion, laceration, and hemorrhage of cerebellum with loss of consciousness greater than 24 hours without return to pre-existing conscious level with patient surviving, sequela |
| Brain Traumatic Injury | ICD 10 | S06.377 | Contusion, laceration, and hemorrhage of cerebellum with loss of consciousness of any duration with death due to brain injury prior to regaining consciousness |
| Brain Traumatic Injury | ICD 10 | S06.377A | Contusion, laceration, and hemorrhage of cerebellum with loss of consciousness of any duration with death due to brain injury prior to regaining consciousness, initial encounter |
| Brain Traumatic Injury | ICD 10 | S06.378 | Contusion, laceration, and hemorrhage of cerebellum with loss of consciousness of any duration with death due to other cause prior to regaining consciousness |
| Brain Traumatic Injury | ICD 10 | S06.378A | Contusion, laceration, and hemorrhage of cerebellum with loss of consciousness of any duration with death due to other cause prior to regaining consciousness, initial encounter |
| Brain Traumatic Injury | ICD 10 | S06.37A | Contusion, laceration, and hemorrhage of cerebellum with loss of consciousness status unknown |
| Brain Traumatic Injury | ICD 10 | S06.37AA | Contusion, laceration, and hemorrhage of cerebellum with loss of consciousness status unknown, initial encounter |
| Brain Traumatic Injury | ICD 10 | S06.37AD | Contusion, laceration, and hemorrhage of cerebellum with loss of consciousness status unknown, subsequent encounter |
| Brain Traumatic Injury | ICD 10 | S06.37AS | Contusion, laceration, and hemorrhage of cerebellum with loss of consciousness status unknown, sequela |
| Brain Traumatic Injury | ICD 10 | S06.379 | Contusion, laceration, and hemorrhage of cerebellum with loss of consciousness of unspecified duration |

| | | | |
|---|---|---|---|
| Brain Traumatic Injury | ICD 10 | S06.379A | Contusion, laceration, and hemorrhage of cerebellum with loss of consciousness of unspecified duration, initial encounter |
| Brain Traumatic Injury | ICD 10 | S06.379D | Contusion, laceration, and hemorrhage of cerebellum with loss of consciousness of unspecified duration, subsequent encounter |
| Brain Traumatic Injury | ICD 10 | S06.379S | Contusion, laceration, and hemorrhage of cerebellum with loss of consciousness of unspecified duration, sequela |
| Brain Traumatic Injury | ICD 10 | S06.38 | Contusion, laceration, and hemorrhage of brainstem |
| Brain Traumatic Injury | ICD 10 | S06.380 | Contusion, laceration, and hemorrhage of brainstem without loss of consciousness |
| Brain Traumatic Injury | ICD 10 | S06.380A | Contusion, laceration, and hemorrhage of brainstem without loss of consciousness, initial encounter |
| Brain Traumatic Injury | ICD 10 | S06.380D | Contusion, laceration, and hemorrhage of brainstem without loss of consciousness, subsequent encounter |
| Brain Traumatic Injury | ICD 10 | S06.380S | Contusion, laceration, and hemorrhage of brainstem without loss of consciousness, sequela |
| Brain Traumatic Injury | ICD 10 | S06.381 | Contusion, laceration, and hemorrhage of brainstem with loss of consciousness of 30 minutes or less |
| Brain Traumatic Injury | ICD 10 | S06.381A | Contusion, laceration, and hemorrhage of brainstem with loss of consciousness of 30 minutes or less, initial encounter |
| Brain Traumatic Injury | ICD 10 | S06.381D | Contusion, laceration, and hemorrhage of brainstem with loss of consciousness of 30 minutes or less, subsequent encounter |
| Brain Traumatic Injury | ICD 10 | S06.381S | Contusion, laceration, and hemorrhage of brainstem with loss of consciousness of 30 minutes or less, sequela |
| Brain Traumatic Injury | ICD 10 | S06.382 | Contusion, laceration, and hemorrhage of brainstem with loss of consciousness of 31 minutes to 59 minutes |
| Brain Traumatic Injury | ICD 10 | S06.382A | Contusion, laceration, and hemorrhage of brainstem with loss of consciousness of 31 minutes to 59 minutes, initial encounter |

| | | | |
|---|---|---|---|
| Brain Traumatic Injury | ICD 10 | S06.382D | Contusion, laceration, and hemorrhage of brainstem with loss of consciousness of 31 minutes to 59 minutes, subsequent encounter |
| Brain Traumatic Injury | ICD 10 | S06.382S | Contusion, laceration, and hemorrhage of brainstem with loss of consciousness of 31 minutes to 59 minutes, sequela |
| Brain Traumatic Injury | ICD 10 | S06.383 | Contusion, laceration, and hemorrhage of brainstem with loss of consciousness of 1 hour to 5 hours 59 minutes |
| Brain Traumatic Injury | ICD 10 | S06.383A | Contusion, laceration, and hemorrhage of brainstem with loss of consciousness of 1 hour to 5 hours 59 minutes, initial encounter |
| Brain Traumatic Injury | ICD 10 | S06.383D | Contusion, laceration, and hemorrhage of brainstem with loss of consciousness of 1 hour to 5 hours 59 minutes, subsequent encounter |
| Brain Traumatic Injury | ICD 10 | S06.383S | Contusion, laceration, and hemorrhage of brainstem with loss of consciousness of 1 hour to 5 hours 59 minutes, sequela |
| Brain Traumatic Injury | ICD 10 | S06.384 | Contusion, laceration, and hemorrhage of brainstem with loss of consciousness of 6 hours to 24 hours |
| Brain Traumatic Injury | ICD 10 | S06.384A | Contusion, laceration, and hemorrhage of brainstem with loss of consciousness of 6 hours to 24 hours, initial encounter |
| Brain Traumatic Injury | ICD 10 | S06.384D | Contusion, laceration, and hemorrhage of brainstem with loss of consciousness of 6 hours to 24 hours, subsequent encounter |
| Brain Traumatic Injury | ICD 10 | S06.384S | Contusion, laceration, and hemorrhage of brainstem with loss of consciousness of 6 hours to 24 hours, sequela |
| Brain Traumatic Injury | ICD 10 | S06.385 | Contusion, laceration, and hemorrhage of brainstem with loss of consciousness greater than 24 hours with return to pre-existing conscious level |
| Brain Traumatic Injury | ICD 10 | S06.385A | Contusion, laceration, and hemorrhage of brainstem with loss of consciousness greater than 24 hours with return to pre-existing conscious level, initial encounter |
| Brain Traumatic Injury | ICD 10 | S06.385D | Contusion, laceration, and hemorrhage of brainstem with loss of consciousness greater than 24 hours with return to pre-existing conscious level, subsequent encounter |

| | | | |
|---|---|---|---|
| Brain Traumatic Injury | ICD 10 | S06.385S | Contusion, laceration, and hemorrhage of brainstem with loss of consciousness greater than 24 hours with return to pre-existing conscious level, sequela |
| Brain Traumatic Injury | ICD 10 | S06.386 | Contusion, laceration, and hemorrhage of brainstem with loss of consciousness greater than 24 hours without return to pre-existing conscious level with patient surviving |
| Brain Traumatic Injury | ICD 10 | S06.386A | Contusion, laceration, and hemorrhage of brainstem with loss of consciousness greater than 24 hours without return to pre-existing conscious level with patient surviving, initial encounter |
| Brain Traumatic Injury | ICD 10 | S06.386D | Contusion, laceration, and hemorrhage of brainstem with loss of consciousness greater than 24 hours without return to pre-existing conscious level with patient surviving, subsequent encounter |
| Brain Traumatic Injury | ICD 10 | S06.386S | Contusion, laceration, and hemorrhage of brainstem with loss of consciousness greater than 24 hours without return to pre-existing conscious level with patient surviving, sequela |
| Brain Traumatic Injury | ICD 10 | S06.387 | Contusion, laceration, and hemorrhage of brainstem with loss of consciousness of any duration with death due to brain injury prior to regaining consciousness |
| Brain Traumatic Injury | ICD 10 | S06.387A | Contusion, laceration, and hemorrhage of brainstem with loss of consciousness of any duration with death due to brain injury prior to regaining consciousness, initial encounter |
| Brain Traumatic Injury | ICD 10 | S06.388 | Contusion, laceration, and hemorrhage of brainstem with loss of consciousness of any duration with death due to other cause prior to regaining consciousness |
| Brain Traumatic Injury | ICD 10 | S06.388A | Contusion, laceration, and hemorrhage of brainstem with loss of consciousness of any duration with death due to other cause prior to regaining consciousness, initial encounter |
| Brain Traumatic Injury | ICD 10 | S06.38A | Contusion, laceration, and hemorrhage of brainstem with loss of consciousness status unknown |
| Brain Traumatic Injury | ICD 10 | S06.38AA | Contusion, laceration, and hemorrhage of brainstem with loss of consciousness status unknown, initial encounter |
| Brain Traumatic Injury | ICD 10 | S06.38AD | Contusion, laceration, and hemorrhage of brainstem with loss of consciousness status unknown, subsequent encounter |

| | | | |
|---|---|---|---|
| Brain Traumatic Injury | ICD 10 | S06.38AS | Contusion, laceration, and hemorrhage of brainstem with loss of consciousness status unknown, sequela |
| Brain Traumatic Injury | ICD 10 | S06.389 | Contusion, laceration, and hemorrhage of brainstem with loss of consciousness of unspecified duration |
| Brain Traumatic Injury | ICD 10 | S06.389A | Contusion, laceration, and hemorrhage of brainstem with loss of consciousness of unspecified duration, initial encounter |
| Brain Traumatic Injury | ICD 10 | S06.389D | Contusion, laceration, and hemorrhage of brainstem with loss of consciousness of unspecified duration, subsequent encounter |
| Brain Traumatic Injury | ICD 10 | S06.389S | Contusion, laceration, and hemorrhage of brainstem with loss of consciousness of unspecified duration, sequela |
| Brain Traumatic Injury | ICD 9 | V15.52 | History of traumatic brain injury |
| Brain Traumatic Injury | ICD 9 | V80.01 | Traumatic brain injury |
| Amyotrophic lateral sclerosis | ICD 10 | G12.21 | Amyotrophic lateral sclerosis |
| Amyotrophic lateral sclerosis | ICD 9 | 335.20 | Amyotrophic lateral sclerosis |
| Cerebral infarction | ICD 10 | I63 | Cerebral infarction |
| Cerebral infarction | ICD 10 | I63.0 | Cerebral infarction due to thrombosis of precerebral arteries |
| Cerebral infarction | ICD 10 | I63.00 | Cerebral infarction due to thrombosis of unspecified precerebral artery |
| Cerebral infarction | ICD 10 | I63.01 | Cerebral infarction due to thrombosis of vertebral artery |
| Cerebral infarction | ICD 10 | I63.011 | Cerebral infarction due to thrombosis of right vertebral artery |
| Cerebral infarction | ICD 10 | I63.012 | Cerebral infarction due to thrombosis of left vertebral artery |

| | | | |
|---|---|---|---|
| Cerebral infarction | ICD 10 | I63.013 | Cerebral infarction due to thrombosis of bilateral vertebral arteries |
| Cerebral infarction | ICD 10 | I63.019 | Cerebral infarction due to thrombosis of unspecified vertebral artery |
| Cerebral infarction | ICD 10 | I63.02 | Cerebral infarction due to thrombosis of basilar artery |
| Cerebral infarction | ICD 10 | I63.03 | Cerebral infarction due to thrombosis of carotid artery |
| Cerebral infarction | ICD 10 | I63.031 | Cerebral infarction due to thrombosis of right carotid artery |
| Cerebral infarction | ICD 10 | I63.032 | Cerebral infarction due to thrombosis of left carotid artery |
| Cerebral infarction | ICD 10 | I63.033 | Cerebral infarction due to thrombosis of bilateral carotid arteries |
| Cerebral infarction | ICD 10 | I63.039 | Cerebral infarction due to thrombosis of unspecified carotid artery |
| Cerebral infarction | ICD 10 | I63.09 | Cerebral infarction due to thrombosis of other precerebral artery |
| Cerebral infarction | ICD 10 | I63.1 | Cerebral infarction due to embolism of precerebral arteries |
| Cerebral infarction | ICD 10 | I63.10 | Cerebral infarction due to embolism of unspecified precerebral artery |
| Cerebral infarction | ICD 10 | I63.11 | Cerebral infarction due to embolism of vertebral artery |
| Cerebral infarction | ICD 10 | I63.111 | Cerebral infarction due to embolism of right vertebral artery |
| Cerebral infarction | ICD 10 | I63.112 | Cerebral infarction due to embolism of left vertebral artery |
| Cerebral infarction | ICD 10 | I63.113 | Cerebral infarction due to embolism of bilateral vertebral arteries |
| Cerebral infarction | ICD 10 | I63.119 | Cerebral infarction due to embolism of unspecified vertebral artery |

| | | | |
|---|---|---|---|
| Cerebral infarction | ICD 10 | I63.12 | Cerebral infarction due to embolism of basilar artery |
| Cerebral infarction | ICD 10 | I63.13 | Cerebral infarction due to embolism of carotid artery |
| Cerebral infarction | ICD 10 | I63.131 | Cerebral infarction due to embolism of right carotid artery |
| Cerebral infarction | ICD 10 | I63.132 | Cerebral infarction due to embolism of left carotid artery |
| Cerebral infarction | ICD 10 | I63.133 | Cerebral infarction due to embolism of bilateral carotid arteries |
| Cerebral infarction | ICD 10 | I63.139 | Cerebral infarction due to embolism of unspecified carotid artery |
| Cerebral infarction | ICD 10 | I63.19 | Cerebral infarction due to embolism of other precerebral artery |
| Cerebral infarction | ICD 10 | I63.2 | Cerebral infarction due to unspecified occlusion or stenosis of precerebral arteries |
| Cerebral infarction | ICD 10 | I63.20 | Cerebral infarction due to unspecified occlusion or stenosis of unspecified precerebral arteries |
| Cerebral infarction | ICD 10 | I63.21 | Cerebral infarction due to unspecified occlusion or stenosis of vertebral arteries |
| Cerebral infarction | ICD 10 | I63.211 | Cerebral infarction due to unspecified occlusion or stenosis of right vertebral artery |
| Cerebral infarction | ICD 10 | I63.212 | Cerebral infarction due to unspecified occlusion or stenosis of left vertebral artery |
| Cerebral infarction | ICD 10 | I63.213 | Cerebral infarction due to unspecified occlusion or stenosis of bilateral vertebral arteries |
| Cerebral infarction | ICD 10 | I63.219 | Cerebral infarction due to unspecified occlusion or stenosis of unspecified vertebral artery |
| Cerebral infarction | ICD 10 | I63.22 | Cerebral infarction due to unspecified occlusion or stenosis of basilar artery |
| Cerebral infarction | ICD 10 | I63.23 | Cerebral infarction due to unspecified occlusion or stenosis of carotid arteries |

| | | | |
|---|---|---|---|
| Cerebral infarction | ICD 10 | I63.231 | Cerebral infarction due to unspecified occlusion or stenosis of right carotid arteries |
| Cerebral infarction | ICD 10 | I63.232 | Cerebral infarction due to unspecified occlusion or stenosis of left carotid arteries |
| Cerebral infarction | ICD 10 | I63.233 | Cerebral infarction due to unspecified occlusion or stenosis of bilateral carotid arteries |
| Cerebral infarction | ICD 10 | I63.239 | Cerebral infarction due to unspecified occlusion or stenosis of unspecified carotid artery |
| Cerebral infarction | ICD 10 | I63.29 | Cerebral infarction due to unspecified occlusion or stenosis of other precerebral arteries |
| Cerebral infarction | ICD 10 | I63.3 | Cerebral infarction due to thrombosis of cerebral arteries |
| Cerebral infarction | ICD 10 | I63.30 | Cerebral infarction due to thrombosis of unspecified cerebral artery |
| Cerebral infarction | ICD 10 | I63.31 | Cerebral infarction due to thrombosis of middle cerebral artery |
| Cerebral infarction | ICD 10 | I63.311 | Cerebral infarction due to thrombosis of right middle cerebral artery |
| Cerebral infarction | ICD 10 | I63.312 | Cerebral infarction due to thrombosis of left middle cerebral artery |
| Cerebral infarction | ICD 10 | I63.313 | Cerebral infarction due to thrombosis of bilateral middle cerebral arteries |
| Cerebral infarction | ICD 10 | I63.319 | Cerebral infarction due to thrombosis of unspecified middle cerebral artery |
| Cerebral infarction | ICD 10 | I63.32 | Cerebral infarction due to thrombosis of anterior cerebral artery |
| Cerebral infarction | ICD 10 | I63.321 | Cerebral infarction due to thrombosis of right anterior cerebral artery |
| Cerebral infarction | ICD 10 | I63.322 | Cerebral infarction due to thrombosis of left anterior cerebral artery |
| Cerebral infarction | ICD 10 | I63.323 | Cerebral infarction due to thrombosis of bilateral anterior cerebral arteries |

| | | | |
|---|---|---|---|
| Cerebral infarction | ICD 10 | I63.329 | Cerebral infarction due to thrombosis of unspecified anterior cerebral artery |
| Cerebral infarction | ICD 10 | I63.33 | Cerebral infarction due to thrombosis of posterior cerebral artery |
| Cerebral infarction | ICD 10 | I63.331 | Cerebral infarction due to thrombosis of right posterior cerebral artery |
| Cerebral infarction | ICD 10 | I63.332 | Cerebral infarction due to thrombosis of left posterior cerebral artery |
| Cerebral infarction | ICD 10 | I63.333 | Cerebral infarction due to thrombosis of bilateral posterior cerebral arteries |
| Cerebral infarction | ICD 10 | I63.339 | Cerebral infarction due to thrombosis of unspecified posterior cerebral artery |
| Cerebral infarction | ICD 10 | I63.34 | Cerebral infarction due to thrombosis of cerebellar artery |
| Cerebral infarction | ICD 10 | I63.341 | Cerebral infarction due to thrombosis of right cerebellar artery |
| Cerebral infarction | ICD 10 | I63.342 | Cerebral infarction due to thrombosis of left cerebellar artery |
| Cerebral infarction | ICD 10 | I63.343 | Cerebral infarction due to thrombosis of bilateral cerebellar arteries |
| Cerebral infarction | ICD 10 | I63.349 | Cerebral infarction due to thrombosis of unspecified cerebellar artery |
| Cerebral infarction | ICD 10 | I63.39 | Cerebral infarction due to thrombosis of other cerebral artery |
| Cerebral infarction | ICD 10 | I63.4 | Cerebral infarction due to embolism of cerebral arteries |
| Cerebral infarction | ICD 10 | I63.40 | Cerebral infarction due to embolism of unspecified cerebral artery |
| Cerebral infarction | ICD 10 | I63.41 | Cerebral infarction due to embolism of middle cerebral artery |
| Cerebral infarction | ICD 10 | I63.411 | Cerebral infarction due to embolism of right middle cerebral artery |

| | | | |
|---|---|---|---|
| Cerebral infarction | ICD 10 | I63.412 | Cerebral infarction due to embolism of left middle cerebral artery |
| Cerebral infarction | ICD 10 | I63.413 | Cerebral infarction due to embolism of bilateral middle cerebral arteries |
| Cerebral infarction | ICD 10 | I63.419 | Cerebral infarction due to embolism of unspecified middle cerebral artery |
| Cerebral infarction | ICD 10 | I63.42 | Cerebral infarction due to embolism of anterior cerebral artery |
| Cerebral infarction | ICD 10 | I63.421 | Cerebral infarction due to embolism of right anterior cerebral artery |
| Cerebral infarction | ICD 10 | I63.422 | Cerebral infarction due to embolism of left anterior cerebral artery |
| Cerebral infarction | ICD 10 | I63.423 | Cerebral infarction due to embolism of bilateral anterior cerebral arteries |
| Cerebral infarction | ICD 10 | I63.429 | Cerebral infarction due to embolism of unspecified anterior cerebral artery |
| Cerebral infarction | ICD 10 | I63.43 | Cerebral infarction due to embolism of posterior cerebral artery |
| Cerebral infarction | ICD 10 | I63.431 | Cerebral infarction due to embolism of right posterior cerebral artery |
| Cerebral infarction | ICD 10 | I63.432 | Cerebral infarction due to embolism of left posterior cerebral artery |
| Cerebral infarction | ICD 10 | I63.433 | Cerebral infarction due to embolism of bilateral posterior cerebral arteries |
| Cerebral infarction | ICD 10 | I63.439 | Cerebral infarction due to embolism of unspecified posterior cerebral artery |
| Cerebral infarction | ICD 10 | I63.44 | Cerebral infarction due to embolism of cerebellar artery |
| Cerebral infarction | ICD 10 | I63.441 | Cerebral infarction due to embolism of right cerebellar artery |
| Cerebral infarction | ICD 10 | I63.442 | Cerebral infarction due to embolism of left cerebellar artery |

| | | | |
|---|---|---|---|
| Cerebral infarction | ICD 10 | I63.443 | Cerebral infarction due to embolism of bilateral cerebellar arteries |
| Cerebral infarction | ICD 10 | I63.449 | Cerebral infarction due to embolism of unspecified cerebellar artery |
| Cerebral infarction | ICD 10 | I63.49 | Cerebral infarction due to embolism of other cerebral artery |
| Cerebral infarction | ICD 10 | I63.5 | Cerebral infarction due to unspecified occlusion or stenosis of cerebral arteries |
| Cerebral infarction | ICD 10 | I63.50 | Cerebral infarction due to unspecified occlusion or stenosis of unspecified cerebral artery |
| Cerebral infarction | ICD 10 | I63.51 | Cerebral infarction due to unspecified occlusion or stenosis of middle cerebral artery |
| Cerebral infarction | ICD 10 | I63.511 | Cerebral infarction due to unspecified occlusion or stenosis of right middle cerebral artery |
| Cerebral infarction | ICD 10 | I63.512 | Cerebral infarction due to unspecified occlusion or stenosis of left middle cerebral artery |
| Cerebral infarction | ICD 10 | I63.513 | Cerebral infarction due to unspecified occlusion or stenosis of bilateral middle cerebral arteries |
| Cerebral infarction | ICD 10 | I63.519 | Cerebral infarction due to unspecified occlusion or stenosis of unspecified middle cerebral artery |
| Cerebral infarction | ICD 10 | I63.52 | Cerebral infarction due to unspecified occlusion or stenosis of anterior cerebral artery |
| Cerebral infarction | ICD 10 | I63.521 | Cerebral infarction due to unspecified occlusion or stenosis of right anterior cerebral artery |
| Cerebral infarction | ICD 10 | I63.522 | Cerebral infarction due to unspecified occlusion or stenosis of left anterior cerebral artery |
| Cerebral infarction | ICD 10 | I63.523 | Cerebral infarction due to unspecified occlusion or stenosis of bilateral anterior cerebral arteries |
| Cerebral infarction | ICD 10 | I63.529 | Cerebral infarction due to unspecified occlusion or stenosis of unspecified anterior cerebral artery |
| Cerebral infarction | ICD 10 | I63.53 | Cerebral infarction due to unspecified occlusion or stenosis of posterior cerebral artery |

| | | | |
|---|---|---|---|
| Cerebral infarction | ICD 10 | I63.531 | Cerebral infarction due to unspecified occlusion or stenosis of right posterior cerebral artery |
| Cerebral infarction | ICD 10 | I63.532 | Cerebral infarction due to unspecified occlusion or stenosis of left posterior cerebral artery |
| Cerebral infarction | ICD 10 | I63.533 | Cerebral infarction due to unspecified occlusion or stenosis of bilateral posterior cerebral arteries |
| Cerebral infarction | ICD 10 | I63.539 | Cerebral infarction due to unspecified occlusion or stenosis of unspecified posterior cerebral artery |
| Cerebral infarction | ICD 10 | I63.54 | Cerebral infarction due to unspecified occlusion or stenosis of cerebellar artery |
| Cerebral infarction | ICD 10 | I63.541 | Cerebral infarction due to unspecified occlusion or stenosis of right cerebellar artery |
| Cerebral infarction | ICD 10 | I63.542 | Cerebral infarction due to unspecified occlusion or stenosis of left cerebellar artery |
| Cerebral infarction | ICD 10 | I63.543 | Cerebral infarction due to unspecified occlusion or stenosis of bilateral cerebellar arteries |
| Cerebral infarction | ICD 10 | I63.549 | Cerebral infarction due to unspecified occlusion or stenosis of unspecified cerebellar artery |
| Cerebral infarction | ICD 10 | I63.59 | Cerebral infarction due to unspecified occlusion or stenosis of other cerebral artery |
| Cerebral infarction | ICD 10 | I63.6 | Cerebral infarction due to cerebral venous thrombosis, nonpyogenic |
| Cerebral infarction | ICD 10 | I63.8 | Other cerebral infarction |
| Cerebral infarction | ICD 10 | I63.81 | Other cerebral infarction due to occlusion or stenosis of small artery |
| Cerebral infarction | ICD 10 | I63.89 | Other cerebral infarction |
| Cerebral infarction | ICD 10 | I63.9 | Cerebral infarction, unspecified |
| Cerebral infarction | ICD 9 | 434 | Occlusion of cerebral arteries |

| | | | |
|---|---|---|---|
| Cerebral infarction | ICD 9 | 434.0 | Cerebral thrombosis |
| Cerebral infarction | ICD 9 | 434.00 | Cerebral thrombosis without mention of cerebral infarction |
| Cerebral infarction | ICD 9 | 434.01 | Cerebral thrombosis with cerebral infarction |
| Cerebral infarction | ICD 9 | 434.1 | Cerebral embolism |
| Cerebral infarction | ICD 9 | 434.10 | Cerebral embolism without mention of cerebral infarction |
| Cerebral infarction | ICD 9 | 434.11 | Cerebral embolism with cerebral infarction |
| Cerebral infarction | ICD 9 | 434.9 | Cerebral artery occlusion unspecified |
| Cerebral infarction | ICD 9 | 434.90 | Cerebral artery occlusion, unspecified without mention of cerebral infarction |
| Cerebral infarction | ICD 9 | 434.91 | Cerebral artery occlusion, unspecified with cerebral infarction |
